# Psychometric and biomedical outcomes of glycated haemoglobin target-setting in adults with type 1 and type 2 diabetes: A mixed-methods parallel-group randomised feasibility study

**DOI:** 10.1101/2025.01.11.24319632

**Authors:** Samuel J Westall, Simon Watmough, Ram Prakash Narayanan, Greg Irving, Kevin Hardy

## Abstract

**Background:** Glycated haemoglobin (HbA_1c_) targets are commonly used to guide patient management in diabetes to reduce future risk of diabetes complications, but little is known of the psychological impact of HbA_1c_ target-setting. We explored the feasibility of undertaking a conclusive study evaluating the impact of setting explicit HbA_1c_ targets in adults with diabetes.

**Methods:** A randomised, mixed-methods study design was used to quantitatively and qualitatively evaluate the psychometric and biomedical impact of intensified or relaxed glycaemic targets in adults with diabetes. Alongside baseline measurement of HbA_1c_, blood pressure and body mass index, patients completed baseline validated psychometric questionnaires (EuroQoL-5D-5L, Problem Areas In Diabetes, Summary of Diabetes Self-Care Activities, Well-Being Quetionnaire-12, Diabetes Empowerment Scale-Long Form) and were randomised 1:1. Participants in group A received explicit HbA_1c_ target intervention targets 5 mmol/mol above current HbA_1c_. Participants in group B received explicit HbA_1c_ targets 5 mmol/mol below current HbA_1c_. Rates of eligibility, recruitment, retention and questionnaire response were recorded. Outcomes were re-measured 3-months post-intervention. Patients and healthcare professionals attended semi-structured interviews for qualitative evaluation.

**Results:** Fifty participants were recruited. Withdrawal rate was 34% (n=17). Endpoint evaluation revealed no significant between-group differences in patient-reported outcome measures or HbA_1c_ levels. Overall, levels of distress (-4.4, p=.009), self-efficacy (.25 [.09–.41], p=.004) and subsequent HbA_1c_ readings (-2.8 [-5.0–-.7], p=.012) improved, with non-significant changes seen in health-related quality of life, wellbeing, and self-care. Patients and healthcare professional interviews demonstrated study acceptability alongside specific motivators (e.g., target achievability, hypoglycaemia avoidance) and demotivators (e.g., lack of understanding, lack of target achievability) for striving to reach glycaemic targets. Combined qualitative data from patient and healthcare professional interviews and quantitative study aspects triangulated, enhancing data trustworthiness, and informing future hypotheses and methodologies.

**Discussion:** This mixed-methods study demonstrates feasibility and provides a novel insight into the psychological implications of HbA_1c_ target-setting.

**Trial registration:** The study is registered with the ISRCTN (registration number: <u>12461724</u>; date registered: 11^th^ June 2021).

## Introduction

### Background

Glycated haemoglobin values are commonly used to guide the management of people with diabetes. Far from being a standardised approach, glycated haemoglobin targets are often individualised to the patient in response to the presence of established cardiovascular disease [1–3], advanced age [4,5], long diabetes duration [6], frailty [7,8], excessive comorbidity [9], history of severe hypoglycaemia [10] and comorbid mental health illness [11,12].

For diabetes healthcare professionals, the importance of a personalised approach to management and goal-setting in people with diabetes has never been clearer, evidenced by the release of the joint American Diabetes Association (ADA)/European Association for the Study of Diabetes (EASD) consensus statement [13,14]. Over time, glycated haemoglobin is a predictor of risk of developing diabetes complications [15]. Chronically high glycated haemoglobin readings have been shown to negatively impact upon the physical health of people with diabetes: diabetes is one of the leading causes of adult blindness in the UK [16], increases the risk of cancer [17,18] and dementia [19,20], more than doubles the risk of heart attack [21], trebles the risk of stroke [21] and is the commonest cause of end-stage renal failure [21] and non-traumatic lower limb amputation in Europe [21].

Several factors have a bearing on the achievement of glycated haemoglobin targets in people with diabetes [22]. Diabetes is recognised to have a significant impact on psychological outcomes and mental health. Indeed, mental health illnesses are seen with increased prevalence in those with diabetes [23]. Prior research has shown that a significant proportion of people with diabetes have depression at a level that impairs activities of daily living, quality of life, adherence to medical treatment, glycaemic control, and increases healthcare utilisation, healthcare cost and the risk of diabetes complications [11,12,24,25]. From a patient’s perspective, lower levels of motivation and knowledge and higher levels of mental health comorbidity can create barriers to achieving optimal glycated haemoglobin levels [26]. From a healthcare perspective, contradictory guidelines and complex management options can cause confusion for clinicians in personalising glycated haemoglobin targets in patients [22].

Although awareness of the importance of achieving glycated haemoglobin targets amongst patients with diabetes is improving with patient education [27], many patients remain unaware of the preventable excess risk posed by sub-optimal glycated haemoglobin levels. Despite many new pharmacological interventions that have become available in the past decade, achievement of glycated haemoglobin treatment targets in England and Wales has remained stubbornly below 30 percent for Type 1 Diabetes and below 70 percent for Type 2 Diabetes [28]. Evidence for lack of progress in the achievement of optimal glycated haemoglobin levels is reflected in studies globally [26]. People with diabetes have historically struggled to access evidenced-based diabetes education (though this is improving) and negative patient perceptions on the intensification of diabetes treatment remain prevalent [26,29,30].

Awareness amongst healthcare professionals of the importance using treatment targets to drive improvement in micro- and macro-vascular outcomes is good. Compelling evidence on the use of personalised glycated haemoglobin treatment targets in response to patient factors is gaining traction amongst clinicians in primary and secondary care in the UK. Current literature points to the clear reduction in the risk of diabetes complications by having individualised treatment targets [3,31–34]. A lack of evidence persists on the impact clinician-initiated glycated haemoglobin target-setting has on psychological and self-management abilities of people with diabetes. It would therefore be valuable to determine the acceptability and preliminary impact of glycated haemoglobin target-setting on the psychological well-being of people with diabetes.

### Aims and objectives

#### Aims

The aim of this study was to evaluate the preliminary impact of relaxed or intensified glycated haemoglobin targets on patient reported outcomes (PROs) and subsequent glycated haemoglobin (HbA_1c_) levels with respect to:

i. Identifying methodological, recruitment and retention issues in preparation for a large-scale study.
ii. Providing preliminary indications of the clinical utility of specific glycated haemoglobin targets in people with diabetes.

#### Objectives

The objectives of this study are to evaluate:

1. The feasibility of study through observation of recruitment, retention and data collection adequacy.
2. The preliminary impact of the intervention on health-related quality of life, diabetes-distress, self-care, well-being, diabetes-related psychosocial self-efficacy and subsequent glycated haemoglobin levels, blood pressure (BP) and Body Mass Index (BMI) at 3-month follow-up.
3. The acceptability of the intervention and study processes.
4. The experiences, views and opinions of people with diabetes and their healthcare professionals on the acceptability of the study, the intervention and their wellbeing.

## Materials and methods

The research protocol was prospectively registered with the ISRCTN and available to view online (ISRCTN 12461724). A detailed description of the methods has been published and is available to view [35].

Briefly, the study was completed between May 2021 and May 2022. Setup and delivery was sponsored by St Helens and Knowsley Teaching Hospitals NHS Trust (ref: STHK-2021-003). This non-commercial single-site study was undertaken as part of the corresponding author’s PhD [36] in a salaried clinical research fellow post on-site at the NHS Trust. The study received ethical approval from the UK Health Research Authority (Cornwall and Plymouth Research Ethics Committee, REC reference 21/SW/0043, IRAS ID: 291254, 30^th^ April 2021, see supporting information S2).

### Participants

The participants were adults with type 1 or type 2 diabetes aged 18 and over with a glycated haemoglobin reading between 64 and 125 mmol/mol. Full exclusion criteria are available to view in the published protocol [35].

### Recruitment

We planned to recruit a minimum of 50 patients attending secondary care diabetes clinic appointments over a 4-month period. Sample size was determined according to the published protocol [35]. Recruitment was undertaken by the researcher working in liaison with the diabetes team to identify individuals who met the inclusion and exclusion criteria. An invitation letter with accompanying participant information sheet was posted to eligible individuals prior to their clinic visit. All patient-facing study-related documents were reviewed for face and content validity with a service user group, with adaptations made where clarification was required. Written informed consent was obtained from all participants, with the original copy filed in the study site files and copies of signed documentation provided to participants.

A stratified purposive sample of participants involved in the study were selected for semi-structured interviews with the researcher. Additionally, a convenience sample of diabetes healthcare professionals were interviewed to obtain an additional perspective on the use of glycated haemoglobin targets in people with diabetes.

### Intervention

Participants were randomly allocated (stratified – Type 1 Diabetes/Type 2 Diabetes, random permuted block 1:1, as per protocol [35]) to receive relaxed or intensified glycated haemoglobin targets as described using the TIDieR checklist in Table 1 [37], extracted from the published protocol [35].

**Table 1.**
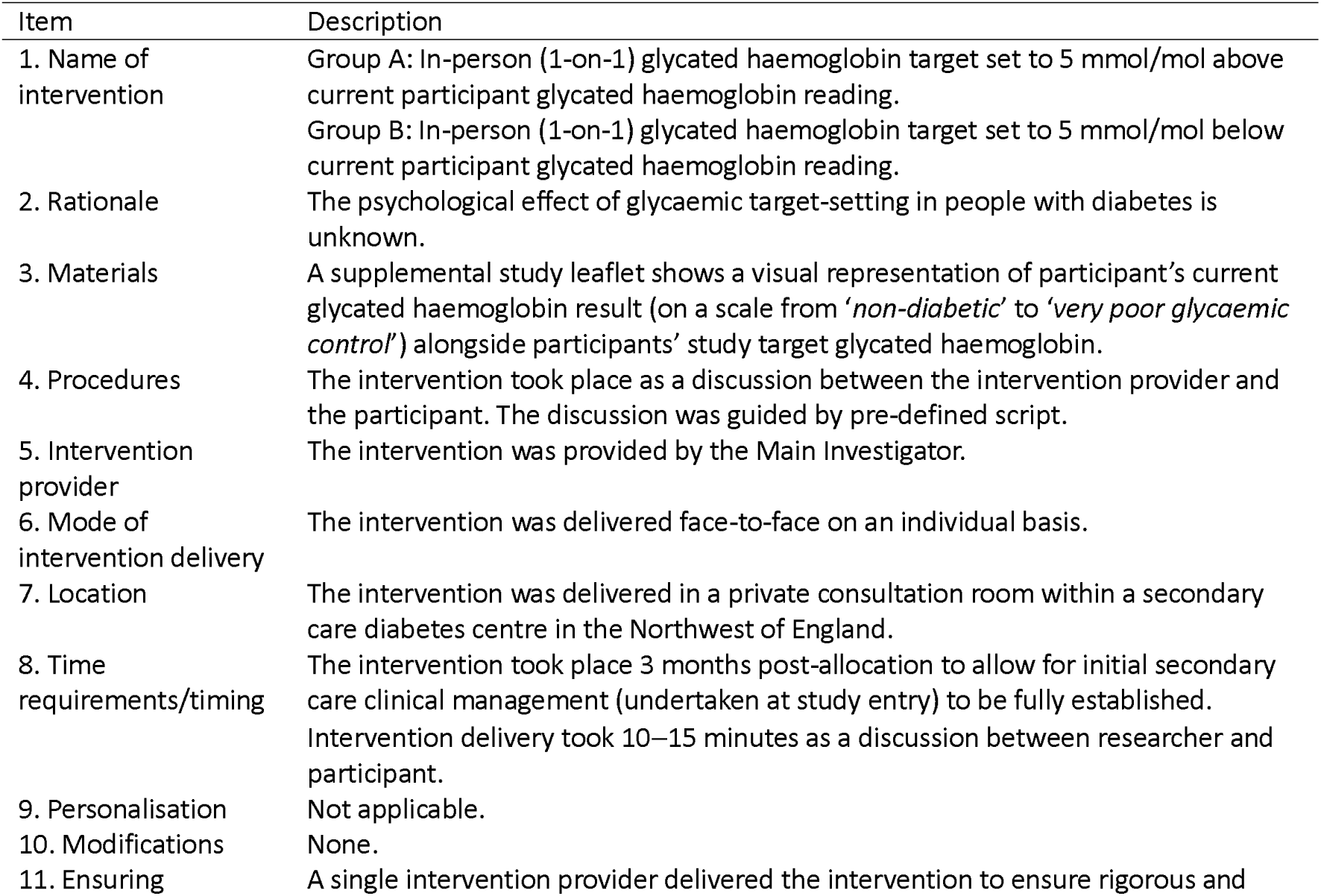
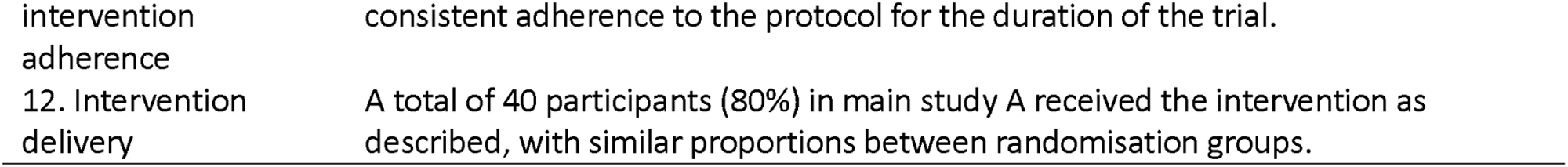
Study intervention described using the TIDieR checklist. [37]

A schedule of procedures and events is presented in Table 2, in accordance with the published protocol and the 2013 SPIRIT statement [35,38].

**Table 2.** Schedule of procedures and assessments.

| Study period | Pre-baseline | Baseline | Intervention | Endpoint | Interviews |
| --- | --- | --- | --- | --- | --- |
| Time point | - | 0 months | 3 months | 6 months | 0–12 months |
| **Screening** |  |  |  |  |  |
| Screen electronic records | X |  |  |  |  |
| Postal invitation | X |  |  |  |  |
| **Enrolment** |  |  |  |  |  |
| Check eligibility |  | X |  |  |  |
| Informed consent |  | X |  |  |  |
| Allocation |  | X |  |  |  |
| 3-month run-in |  | X-----X |  |  |  |
| **Intervention** |  |  |  |  |  |
| Group A or B intervention |  |  | X |  |  |
| **Assessments** |  |  |  |  |  |
| POC glycated haemoglobin |  | X | X | X |  |
| BP, BMI |  | X | X | X |  |
| EQ-5D-5L, PAID, SDSCA, W-BQ12, DES-LF |  |  | X | X |  |
| Acceptability survey |  |  |  | X |  |
| **Additional elements** |  |  |  |  |  |
| Interview with participants<sup>†</sup> |  |  |  |  | X |
| Interview with HCPs<sup>‡</sup> |  |  |  |  | X |

POC: point-of-care; HbA<sub>1c</sub>: glycated haemoglobin; BP: blood pressure; BMI: body mass index; EQ-5D-5L:
EuroQoL-5D-5L; PAID: Problem Areas in Diabetes; SDSCA: Summary of Diabetes Self-care Activities; W-BQ12:
Well-being Questionnaire-12; DES-LF: Diabetes Empowerment Scale-Long Form; HCP: Healthcare
Professional; mmol/mol: millimoles per mole, glycated haemoglobin SI unit of measurement
<sup>†</sup> Sub-study B. Semi-structured interviews in those enrolled in sub-study A.
<sup>‡</sup> Sub-study C. Semi-structured interviews with healthcare professionals.
<sup>§</sup> Sub-study D. In those declining to take part in sub-study A, survey and semi-structured interview.

### Outcomes

#### Quantitative feasibility outcome data

Alongside rates of participant eligibility, recruitment, retention and questionnaire response, acceptability data were captured using an end-of-study Likert-type survey with free-text response options.

#### Other quantitative data

Baseline and endpoint biomedical outcomes (HbA_1c_, BMI, BP) and PROs of health-related quality of life (Euro-QoL-5D-5L), diabetes-related distress (Problem Areas in Diabetes; PAID), self-care (Summary of Diabetes Self-Care Activities; SDSCA), well-being (Well-Being Questionnaire 12; W-BQ12), and self-efficacy (Diabetes Empowerment Scale-Long Form; DES-LF) were collected.

#### Qualitative data

Two interview sub-studies were run alongside the quantitative study aspects. These sub-studies involved semi-structured interviews with feasibility study participants on study acceptability and their wellbeing, and interviews with healthcare professionals on the use of glycated haemoglobin targets in people with diabetes. Separate invitations letters and information sheets containing interview schedules were sent in advance to participants.

#### Progression criteria

We pre-specified criteria to be met in recommending progression to a conclusive study. These criteria were: recruitment of 50 participants within the 4-month recruitment period, retention of 80% or more at endpoint and questionnaire response rate of 75% or more. Additionally, results of a Likert-type acceptability survey and qualitative data free-text and interview transcript data were considered when determining progression, including the acceptability of study processes.

### Data analysis

Data analysis has been previously described in the published protocol [35]. Briefly, the anonymised quantitative data were analysed in IBM statistical package for social sciences (SPSS) version 25 [39]. Descriptive statistics were used to present participant characteristics and primary feasibility outcomes. Additionally, descriptive statistics were used to summarise pre- and post-intervention glycaemic control (HbA_1c_) and patient-reported psychometric outcomes of health-related quality of life, self-care, diabetes-related distress, wellbeing, and psychosocial self-efficacy for completers.

All quantitative data from questionnaires were checked for missing values and managed according to the accompanying questionnaire user guides. Participant glycated haemoglobin levels (HbA_1c_) were monitored at recruitment and pre- and post-intervention using a point-of-care HbA_1c_ analyser maintained and calibrated as per NHS laboratory standards. Participant characteristics and feasibility outcomes are presented with descriptive statistics, with rates presented as percentages. Continuous variables from PROs and glycated haemoglobin values are presented as means (±standard deviation, SD and 95% confidence interval, CI) or medians (±interquartile range, IQR) depending on data skewness. Data skewness was determined using the Shapiro-Wilk test in SPSS alongside visual inspection of normal Q-Q plots and histograms. Where outliers were noted to skew data, they were removed prior to analysis.

Whilst recognising that feasibility studies are underpowered to consider the use of inferential statistics in determining statistically significant changes in pre- and post-intervention scores in psychometric outcomes, inferential statistics were used to understand any emerging trends in the data. The difference between pre- and post-intervention scores (delta) is denoted by the Greek symbol, Δ. Where pre- and post-intervention psychometric outcome scores were continuous and normally distributed, the paired samples t-test was used. Where distributions were skewed, the Wilcoxon matched-pair signed-rank test was used. Differences between randomisation groups were determined using the independent-sample t-test or the Mann-Whitney U test where data were normally distributed or skewed, respectively.

Qualitative data from semi-structured interviews with patients and healthcare professionals were analysed using the Framework Method of thematic analysis [40] in NVivo qualitative data analysis support software [41]. Anonymised quotes from transcripts were coded for each pre-determined theme of interest with additional inductive themes added as they emerged.

With emergent themes potentially arising in later transcripts, earlier transcripts were re-coded to encompass the additional themes. Coding strategy for transcript analysis was determined by discussion amongst the supervisory team followed by consensus on a final coding strategy. Any disagreements were resolved with input from a senior reviewer (SW). This approach allowed contrast of themes from the patient and healthcare professional perspectives.

Interviewing participants taking part in the quantitative study aspects was undertaken to enable a deeper understanding of the themes of glycated haemoglobin target individualisation, mental health in diabetes and study acceptability and their relationship with health-related quality of life, self-care, diabetes-related distress, and wellbeing.

Source triangulation, as described by Lincoln and Guba [42], was enabled by contrasting qualitative data output from healthcare professionals with data from people with diabetes. This improved qualitative data credibility and consistency, enhancing the trustworthiness of findings. To check the consistency of the interview findings, method triangulation was used to compare the qualitative findings with quantitative results of health-related quality of life, self-care, diabetes-related distress, wellbeing, and diabetes-related psychosocial self-efficacy.

## Results

This study is reported using the CONSORT 2010 reporting guidelines for randomised control trials [43] (see supporting information, S1 CONSORT Checklist). The CONSORT flow diagram is presented in Figure 1. There was no deviation from the approved study protocol (see supporting information S3 Approved protocol). Feasibility data, PRO data, HbA_1c_ levels, BMI, BP, survey results and semi-structure interview transcripts comprise the results of this study. Participant characteristics are presented in Table 3. Participants were predominantly of white British ethnicity.

**Fig 1.**
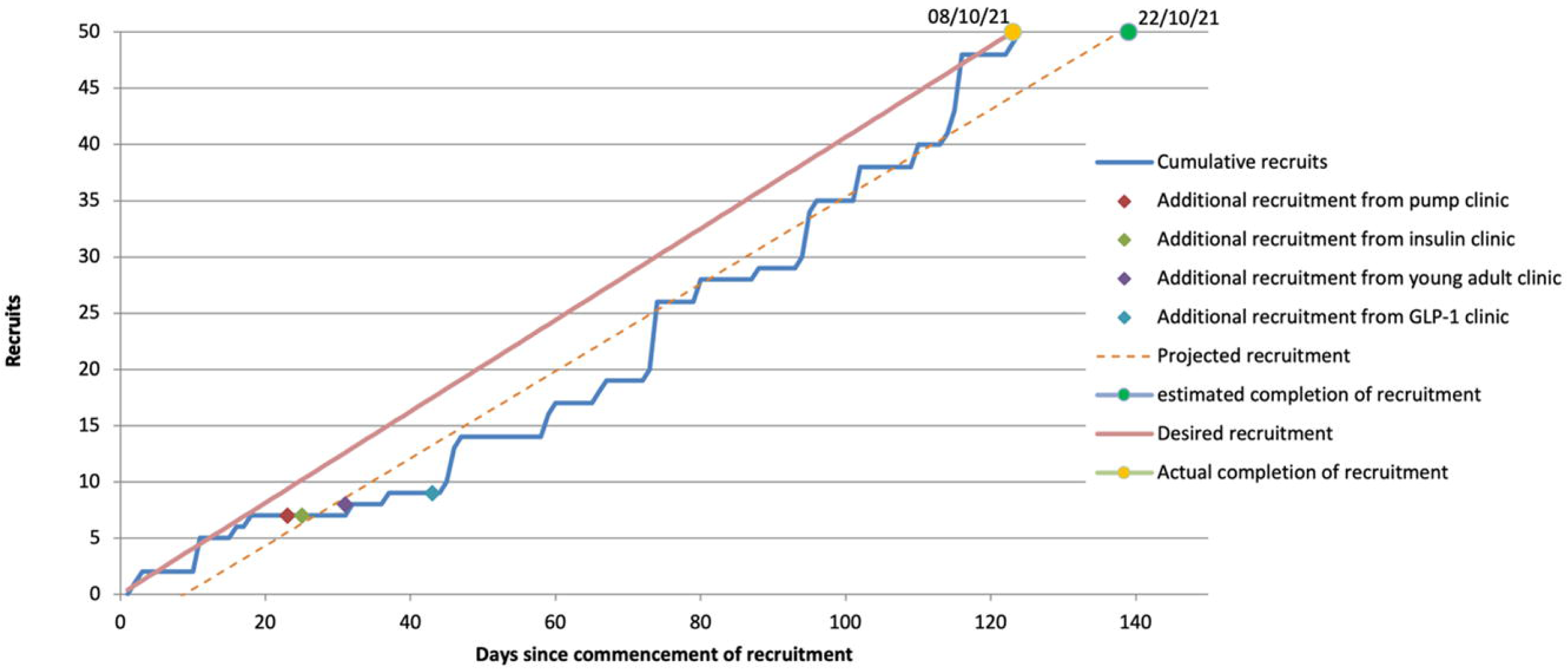
CONSORT flow diagram showing reasons for exclusion, dissent and withdrawal.

**Table 3.** Participant characteristics.

|  | Total sample | Group A | Group B |  | Completers | Non-completers |
| --- | --- | --- | --- | --- | --- | --- |
| Characteristic | n=50 | n=27 | n=23 |  | n=33 | n=17 |
| Group A (%) | 54% | - | - |  | 58% | 47% |
| Completers (%) | 66% | 70% | 61% |  | - | - |
| Age, median (IQR) | 48 (23) | 50 (14) | 46 (36) |  | 51 (17) | 42 (26) |
| Gender (%female) | 40% | 30% | 52% |  | 45% | 29% |
| T1DM (%) | 40% | 41% | 39% |  | 33% | 53% |
| Recruitment BMI, mean (SD) | 31.3 (6.7) | 31.1 (6.8) | 31.6 (6.8) |  | 30.7 (6.7) | 32.5 (6.7) |
| Diabetes duration, mean (SD) | 10.2 (6.5) | 9.9 (6.5) | 10.4 (6.6) |  | 9.8 (6.0) | 10.9 (7.5) |
| IMD decile, median (IQR) | 3 (5) | 3 (5) | 2 (5) |  | 3 (5) | 2 (5) |
| HbA<sub>1c</sub> (recruitment), mean (SD) | 84.4 (14.5) | 83.1 (14.5) | 85.9 (14.6) |  | 83.6 (12.8) | 85.9 (17.5) |

IMD = Index of Multiple Deprivation; IQR = interquartile range; SD = standard deviation

Changes in medication during the run-in period are demonstrated in Figure 2. Predominant changes were cessation of DPP4-i class medications and commencement of insulins and GLP-1 RA class medications. Adjustments to participant diabetes medications between baseline and endpoint were minimal. Figure 2 demonstrates percentage medication usage by medication class at recruitment (0 months), baseline (3 months) and endpoint (6 months).

**Fig 2.**
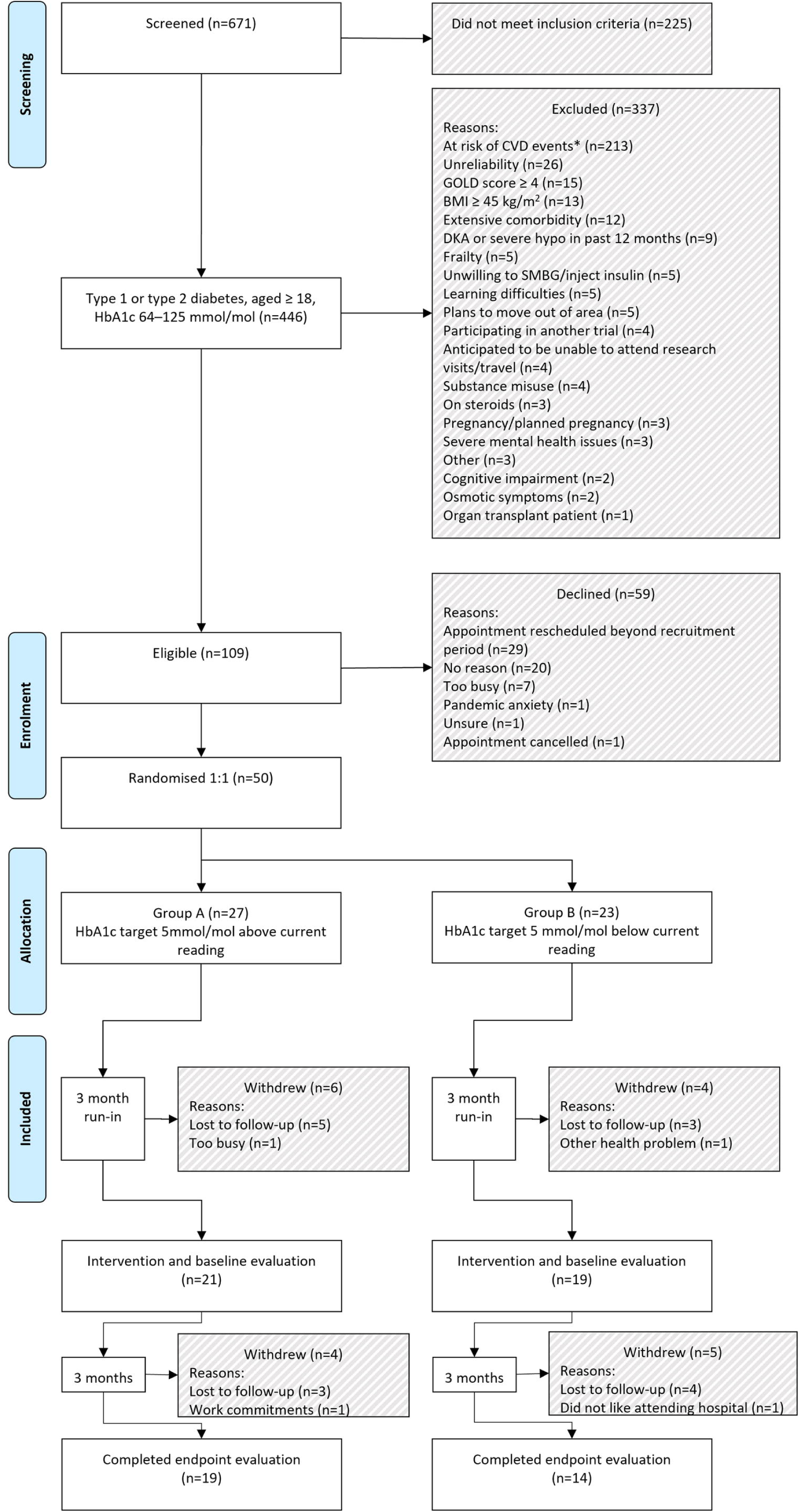
Bar chart showing medication usage at recruitment, baseline and endpoint.

### Feasibility

We planned to recruit 50 participants into the quantitative aspects of the study. Electronic records of 671 people with diabetes due to attend secondary care diabetes clinics were screened for entry. Of those screened, 446 (66.5%) met inclusion criteria. A breakdown of those not meeting inclusion criteria is shown in Table 4.

**Table 4.** Breakdown of inclusion metrics by criterion.

| Criterion | Met<br>n (%) | Not met<br>n (%) |
| --- | --- | --- |
| POC HbA<sub>1c</sub> 64–125 mmol/mol | 464 (69.2) | 207 (30.8) |
| Age 18+ | 671 (100) | 0 (0) |
| Type 1 or type 2 diabetes | 642 (95.7) | 29 (4.3) |
| Type 1 | 265 (39.5) |  |
| Type 2 | 377 (56.2) |  |
| Type 3c |  | 20 (3.0) |
| Gestational |  | 1 (0.1) |
| MODY |  | 1 (0.1) |
| Undetermined |  | 7 (1.0) |

HbA<sub>1c</sub> = glycated haemoglobin; MODY = maturity-onset diabetes of the young; POC = point-of-care.

One-hundred and nine (16.2%) of those screened were eligible for study entry. Reasons for exclusion are shown in Figure 1. Patients at high risk of CV events (Table 5) were excluded due to potentially increased risks associated with adjusting glycated haemoglobin targets in this group.

**Table 5.**
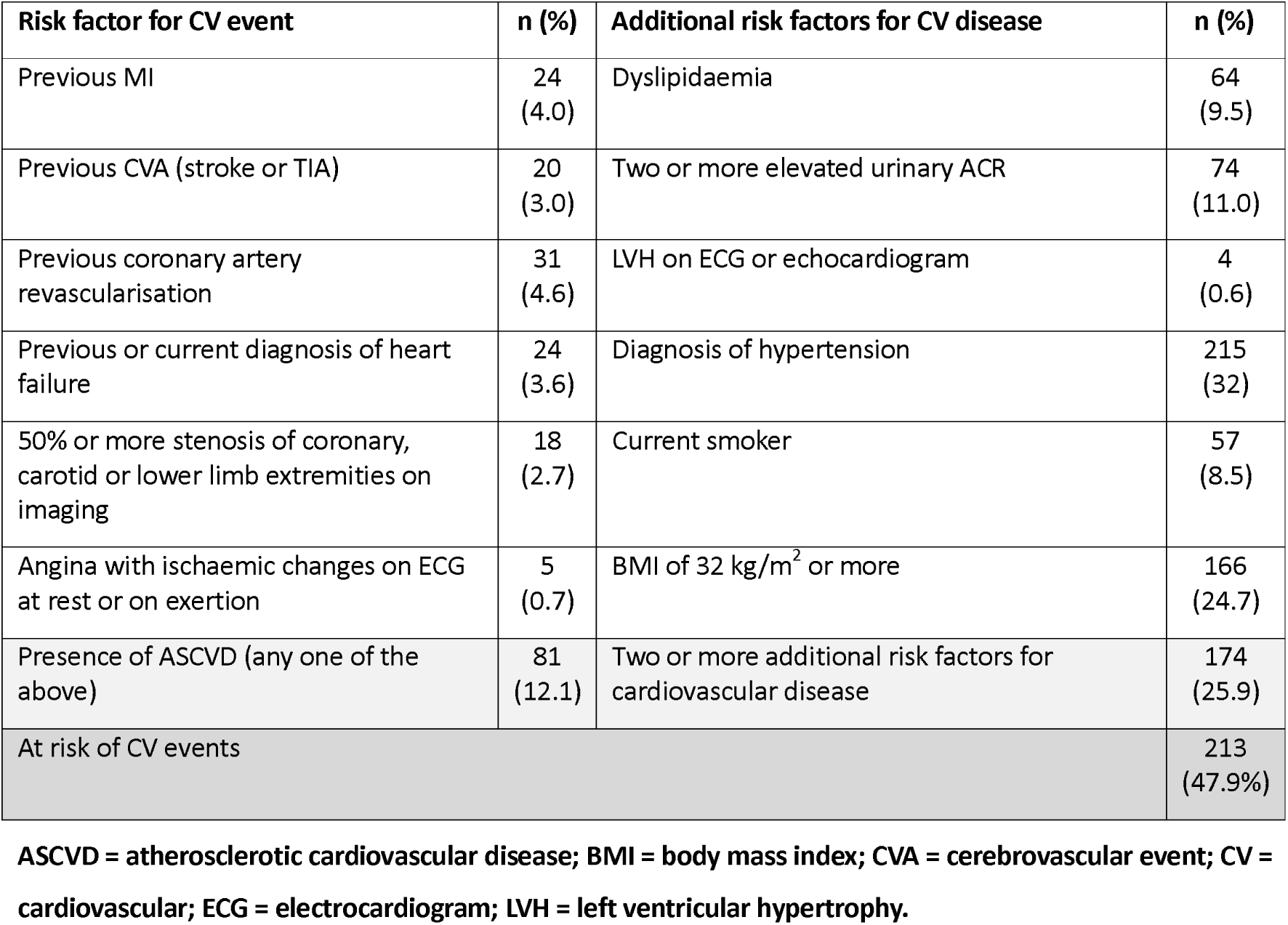
Breakdown of patients meeting inclusion criteria screened for risk of cardiovascular events.

| Risk factor for CV event | n (%) | Additional risk factors for CV disease | n (%) |
| --- | --- | --- | --- |
| Previous MI | 24<br>(4.0) | Dyslipidaemia | 64<br>(9.5) |
| Previous CVA (stroke or TIA) | 20<br>(3.0) | Two or more elevated urinary ACR | 74<br>(11.0) |
| Previous coronary artery revascularisation | 31<br>(4.6) | LVH on ECG or echocardiogram | 4<br>(0.6) |
| Previous or current diagnosis of heart failure | 24<br>(3.6) | Diagnosis of hypertension | 215<br>(32) |
| 50% or more stenosis of coronary, carotid or lower limb extremities on imaging | 18<br>(2.7) | Current smoker | 57<br>(8.5) |
| Angina with ischaemic changes on ECG at rest or on exertion | 5<br>(0.7) | BMI of 32 kg/m<sup>2</sup> or more | 166<br>(24.7) |
| Presence of ASCVD (any one of the above) | 81<br>(12.1) | Two or more additional risk factors for cardiovascular disease | 174<br>(25.9) |
| At risk of CV events |  |  | 213<br>(47.9%) |

ASCVD = atherosclerotic cardiovascular disease; BMI = body mass index; CVA = cerebrovascular event; CV = cardiovascular; ECG = electrocardiogram; LVH = left ventricular hypertrophy.

The required sample was successfully randomised over a 4-month period, with an uptake of 45.9% of those who were eligible to participate. Twenty-seven participants were randomised to Group A, with 23 participants being randomised to Group B. Active monitoring of recruitment rates during the recruitment period noted projected recruitment was below target. Screening of additional participants during the recruitment period allowed for increased recruitment. The graph in Figure 3 demonstrates actual recruitment rates were within the anticipated target. Of those recruited, withdrawal rate was 34% (n=17), with reasons for withdrawal included in the CONSORT flow chart (Figure 1). Thirty-three adults with diabetes completed the study processes, with 19 participants in Group A, and 14 participants in Group B. Non-completers were noted to be significantly younger (Mdn = 42, IQR = 26) versus completers (Mdn = 51, IQR = 17), U = 176.5, p=.033. No associations were detected between non-completion and gender, diabetes-type, diabetes duration, baseline health status, baseline distress, baseline self-care, baseline wellbeing, baseline self-efficacy, intervention group or index of multiple deprivation (IMD) decile.

**Figure 3.**
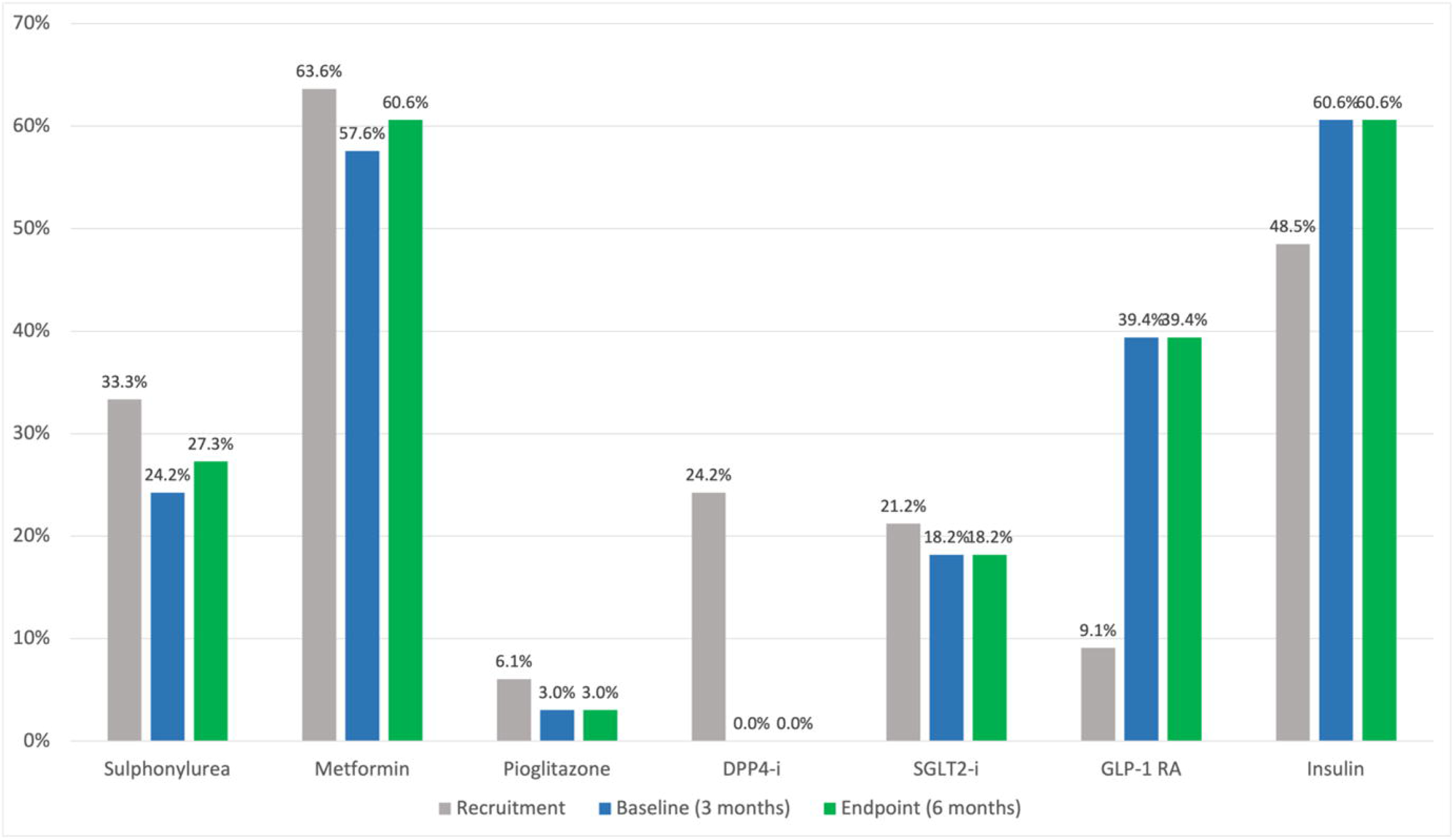
Cumulative recruitment over the 4-month recruitment period.

Response rate for the PROMs was determined on a per-item basis for each questionnaire and each participant. Questionnaire response rate overall was 97.3% in the study completers, or 70.8% if including participants who withdrew. Response rates were similar between groups A (99.7%, or 73.8% when including withdrawers) and B (94.2%, or 67.3% when including withdrawers).

### Effectiveness: glycaemic control

The impact of relaxed (Group A) or intensified (Group B) glycated haemoglobin targets on participants subsequent glycated haemoglobin readings were evaluated.

Where point-of-care glycated haemoglobin was measured in the completers (n=31, outliers removed, 62%), mean (SD) reduction in glycated haemoglobin was 2.8 (95% CI .7−5.0, p=.012) from 69.4 (15.7) at 3 months to 66.6 (16.7) mmol/mol at 6 months. In group A (n=18, outliers removed), mean glycated haemoglobin improved non-significantly by 2.9 (95% CI -.4−6.2, p=.084) from 63.7 (13.6) to 60.8 (13.6) mmol/mol and in group B (n=13, outliers removed) by 2.8 (95% CI -.2−5.7, p=.065) from 77.4 (15.2) to 74.6 (17.7) mmol/mol. Glycated haemoglobin improved to a similar extent in both those with relaxed (group A) or intensified (group B) targets. An independent samples t-test was conducted to compare improvement in glycated haemoglobin in group A and B. There was not a significant difference in improvement in glycated haemoglobin between group B (M = 2.8, SD = 4.9) and group A (M = 2.9, SD = 6.7), t (29)=-.055, p=.957.

### Effectiveness: health-related quality of life

Health status according to the EQ-5D-5L questionnaire outputs two values:

- EQ-5D-5L index, a value between -.594 and 1.000 for the UK with higher values indicating a better health state.
- EQ-VAS, a number derived from a visual analogue scale between 0 (worst health) and 100 (best health).

Minimum clinically important difference (MCID) for EQ-5D-5L index scores is reported in various papers as between 0.030 and 0.074 depending on the condition and country value set used [44–48]. For the UK value set, an MCID of 0.037 is suggested as the smallest detectable change in health-related quality of life according to the EQ-5D-5L that is recognised and valued by the patient [45,46].

At 6 months, 33/50 (66%) of participants completed baseline and endpoint EQ-5D-5L questionnaires. Overall, in the 33 completers, median (IQR) EQ-5D-5L index score reduced from .819 (.285) at 3 months to .795 (.355) at 6 months. In group A completers (n=19), median EQ-5D-5L index score did not change at 3 months, .837 (.248), and 6 months, .837 (.250). In group B completers (n=14), median EQ-5D-5L index score reduced from .759 (.508) to .754 (.337) with the Wilcoxon Signed-Ranks Test indicating that there was no significant difference in baseline and endpoint scores, Z = 19, p=.678. Because the data were skewed, a Wilcoxon Signed-Ranks Test was run and the output indicated that overall, endpoint EQ-5D-5L index scores were not significantly lower than baseline, Z = 155.5, p=.875. The Mann-Whitney U test indicated that baseline EQ-5D-5L index scores for group A (Mdn=.837) were not significantly different from those in group B (Mdn=.759); U = 93.0, p=.152 and endpoint EQ-5D-5L index scores for group A (Mdn=.837) were not significantly different from those in group B (Mdn=.754); U = 92.5, p=.142. Because ΔEQ-5D-5L index scores were not skewed, the independent samples t-test was used to compare delta values between groups. The test indicated that delta values for group A (M=-.030, SD=.123) were not significantly different from those in group B (M=.016, SD=.160); t(29)=-.900, p=.375.

At 6 months, overall median EQ-VAS score improved from 75 (20) to 80 (21) with the Wilcoxon Signed-Ranks Test indicating that endpoint scores were not significantly higher than baseline, Z = 184.5, p=.818. In group A completers, median EQ-VAS score did not change at 3 months, 80 (20), and 6 months, 80 (15). In group B completers, median EQ-VAS score improved from 68 (20) at 3 months to 73 (37) at 6 months with the Wilcoxon Signed-Ranks Test indicating that endpoint scores were not significantly higher than baseline, Z = 39, p=.590. The Mann-Whitney U test indicated that baseline EQ-VAS scores for group A (Mdn=80) were not significantly different from those in group B (Mdn=68); U = 80.5, p=.055, and endpoint EQ-VAS scores for group A (Mdn=80) were not significantly different from those in group B (Mdn=73); U = 80.0, p=.055. Because ΔEQ-VAS scores (n=31, outliers removed) were not skewed, the independent samples t-test was used to compare delta values between groups. The test indicated that delta values for group A (M=-1.65, SD=7.8) were not significantly different from those in group B (M=-1.93, SD=12.9); t(29)=.075, p=.941.

The independent samples t-test was used to compare EQ-5D-5L index delta values between those with type 1 diabetes and those with type 2. The test indicated that delta values for people with type 1 diabetes (M = -.073, SD = .139) were significantly different from those with type 2 (M = .041, SD = .145), mean difference .114 (95% CI .006–.222, p=.038).

Further analysis demonstrated no other significant between-group difference when considering gender and whether participants achieved the HbA_1c_ target set.

### Effectiveness: diabetes-related distress

Where diabetes distress was measured in completers (n=33, 66%), median (IQR) PAID score reduced from 22.5 (26.3) at 3 months to 18.1 (29.4) at 6 months. Because the data were skewed, a Wilcoxon Signed-Ranks Test was run and the output indicated that endpoint PAID scores were significantly lower than baseline, Z = 381.5, p=.009. In group A completers (n=19), median PAID score reduced from 17.5 (26.3) at 3 months to 10.0 (23.8) at 6 months. The Wilcoxon Signed-Ranks Test indicated that endpoint PAID scores for group A were significantly lower than baseline, Z = 137, p=.025. In group B completers (n=14), median PAID score reduced from 28.8 (26.3) at 3 months to 24.4 (39.7) at 6 months. The Wilcoxon Signed-Ranks Test indicated that endpoint PAID scores for group B were not significantly lower than baseline, Z = 66, p=.152. The Mann-Whitney U test indicated that baseline PAID scores for group A (Mdn=17.5) were not significantly different from those in group B (Mdn=28.8); U = 90.0, p=.123 and endpoint PAID scores for group A (Mdn=10.0) were not significantly different from those in group B (Mdn=24.4); U = 102.0, p=.271.

Change in PAID score distributions were not skewed. The independent samples t-test was performed to compare change in PAID score between groups A and B. There was no significant difference in delta values for group A (M=-5.5, SD=9.6) when compared to group B (M=-4.2, SD=12.2); t(31)=-.334, p=.741.

### Effectiveness: wellbeing (general)

Overall median (IQR) general wellbeing increased by 3.0 from 20.0 (15.0) at 3 months to 23.0 (11.0) at 6 months with the Wilcoxon Signed-Ranks Test indicating that there was no significant difference in baseline and endpoint scores, Z = -.943, p=.346.

In group A completers (n=19), median (IQR) general wellbeing increased by 2.0 from 21.0 (14) to 23.0 (10) with the Wilcoxon Signed-Ranks Test indicating that there was no significant difference in baseline and endpoint scores, Z = -.228, p=.820. In group B completers, (n=14), median (IQR) general wellbeing increased by 5.5 from 16.5 (15) to 22.0 (16) with the Wilcoxon Signed-Ranks Test indicating that there was no significant difference in baseline and endpoint scores, Z = -1.102, p=.270. The Mann-Whitney U test indicated that baseline general wellbeing scores for group A (Mdn=21.0, IQR=14) were not significantly different from those in group B (Mdn=16.5, IQR=15); U = 90.5, p=.121, and endpoint general wellbeing scores for group A (Mdn=23.0, IQR=10) were not significantly different from those in group B (Mdn=22.0, IQR=16); U = 122.0, p=.688.

Additionally, after removal of outliers, the Mann-Whitney U test indicated that change in general wellbeing scores for group A (Mdn=0.0) were not significantly different from those in group B (Mdn=-1.0); U = 97.0, p=.746.

There was no statistically significant change in general wellbeing delta values when analysing by gender.

Multiple linear regression analysis was performed to evaluate for confounders with change in the W-BQ12 general wellbeing score (Δgeneral wellbeing) as the dependent variable and potential confounders as independent variables modelled. It was found that none of the confounding variables significantly predicted change in W-BQ12 general wellbeing score (F[5,27] = 1.322, p = ns).

### Effectiveness: wellbeing (positive)

Overall mean (SD) PWB score increased by .2 (95% CI -.8–1.1, p=.750) from 6.5 (2.6) at 3 months to 6.7 (2.7) at 6 months. In group A completers (n=19), mean PWB reduced by .4 (95% CI -.9–1.6, p=.546) from 7.0 (2.5) to 6.6 (2.8). In group B completers (n=14), mean PWB increased by .8 (95% CI **-**.7−2.5, p=.268) from 5.9 (2.6) to 6.7 (2.8).

The independent samples t-test was performed to compare change in PWB score between groups A and B. There was no significant difference in delta values for group A (M=-.4, SD=2.6) when compared to group B (M=.9, SD=2.8); t(31)=-1.300, p=.203.

When comparing outcomes by gender, female (n=15) PWB improved to a significantly greater extent when compared to males (n=18), with a mean difference of 2.0 (95% CI .2– 3.9, p=.028).

Multiple linear regression analysis was performed to evaluate for confounders with change in the W-BQ12 positive wellbeing score (Δpositive wellbeing) as the dependent variable and potential confounders as independent variables modelled. It was found that none of the confounding variables significantly predicted change in W-BQ12 positive wellbeing score (F[5,27] = 2.25, p = ns).

### Effectiveness: wellbeing (negative)

Where NWB was measured in the completers (n=33, 66%), overall median (IQR) score reduced by 1.0 from 3.0 (6) at 3 months to 2.0 (4) at 6 months with the Wilcoxon Signed-Ranks Test indicating that there was no significant difference in baseline and endpoint scores, Z = -1.712, p=.087.

In group A completers (n=19), median (IQR) NWB increased by 1.0 from 1.0 (5) to 2.0 (4) with the Wilcoxon Signed-Ranks Test indicating that there was no significant difference in baseline and endpoint scores, Z = -1.758, p=.079. In group B completers, (n=14), median (IQR) NWB reduced by 0.5 from 3.5 (6) to 3.0 (5) with the Wilcoxon Signed-Ranks Test indicating that there was no significant difference in baseline and endpoint scores, Z = -.517, p=.607. The Mann-Whitney U test indicated that baseline NWB scores for group A (Mdn=1.0, IQR=5) were not significantly different from those in group B (Mdn=3.5, IQR=6); U = 101.5, p=.243, and endpoint NWB scores for group A (Mdn=2.0, IQR=4) were not significantly different from those in group B (Mdn=3.0, IQR=5); U = 91.0, p=.117.

Additionally, the Mann-Whitney U test indicated that change in NWB scores for group A (Mdn=0.0) were not significantly different from those in group B (Mdn=0.0); U = 117.5, p=.559.

There was no statistically significant change in NWB delta values when analysing by gender.

Multiple linear regression analysis was performed to evaluate for confounders with change in the W-BQ12 positive wellbeing score (Δpositive wellbeing) as the dependent variable and potential confounders as independent variables modelled. It was found that none of the confounding variables significantly predicted change in W-BQ12 positive wellbeing score (F[5,27] = 2.25, p = ns).

### Effectiveness: wellbeing (energy)

Overall mean (SD) energy score increased by .6 (95% CI -.3–1.5, p=.207) from 5.1 (3.0) at 3 months to 5.7 (3.0) at 6 months. In group A completers (n=19), mean energy score reduced by .1 (95% CI -.8–1.0, p=.771) from 5.8 (2.8) to 5.7 (2.5). In group B completers (n=14), mean energy score increased by 1.5 (95% CI -.3–3.3, p=.092) from 4.1 (3.1) to 5.6 (3.6).

The independent samples t-test was performed to compare change in energy score between groups A and B. With outliers removed, there was no significant difference in delta values for group A (M=-.1, SD=1.8) when compared to group B (M=.6, SD=2.2); t(29)=-.974, p=.338. There was no statistically significant change in energy delta values when analysing by gender.

Multiple linear regression analysis was performed to evaluate for confounders with change in W-BQ12 energy scores (Δenergy) as the dependent variable and potential confounders as independent variables modelled. It was found that none of the confounding variables significantly predicted change in W-BQ12 energy score (F[5,27] = .36, p = ns).

### Effectiveness: diabetes-related psychosocial self-efficacy (managing psychosocial aspects of diabetes)

Where diabetes-related psychosocial self-efficacy was measured in completers (n=33), mean (SD) improvement in managing psychosocial aspects of diabetes was .27 (95% CI .02−.51, p=.032), from 3.71 (.74) at 3 months to 3.97 (.48) at 6 months. In group A completers (n=19), mean (SD) improvement was .10 (95% CI -.15−.35, p=.420), from 3.89 (.57) to 3.99 (.52). In group B completers (n=14), mean (SD) improvement was .49 (95% CI .02−.96, p=.041), from 3.45 (.88) to 3.94 (.42).

The independent samples t-test was performed to compare change in managing psychosocial aspects of diabetes score between groups A and B. After removing outliers, there was no significant difference in delta values for group A (M=.10, SD=.52) and group B (M=.26, SD=.61); t(29)=-.778, p=.443. These results suggest that the study intervention had no impact on managing psychosocial aspects of diabetes scores.

Multiple linear regression analysis was performed to evaluate for confounders with change in DES-LF managing psychosocial aspects of diabetes scores (ΔDES-LF managing psychosocial aspects of diabetes) as the dependent variable and potential confounders as independent variables modelled. It was found that none of the confounding variables significantly predicted change in DES-LF managing psychosocial aspects of diabetes score (F[5,27] = .75, p = ns).

### Effectiveness: diabetes-related psychosocial self-efficacy (assessing dissatisfaction and readiness for change)

Overall scores for assessing dissatisfaction and readiness to change (n=33, 66%) did not change, with median (IQR) scores at 3 months, 3.89 (.61), and 6 months, 3.89 (.78). In group A completers (n=19), median (IQR) scores improved by .11 from 3.78 (.67) to 3.89 (.89) with the Wilcoxon Signed-Ranks Test indicating there was no significant difference in baseline and endpoint scores, Z = -1.647, p=.100. In group B completers (n=14), median (IQR) scores improved by .11 from 3.89 (.53) to 4.00 (.72) with the Wilcoxon Signed-Ranks Test indicating that there was a significant difference in baseline and endpoint scores, Z = - 2.210, p=.027. The Mann-Whitney U test indicated that baseline scores for group A (Mdn=3.78, IQR=.67) were not significantly different from those in group B (Mdn=3.89, IQR=.53); U = 132.5, p=.985, and endpoint scores for group A (Mdn=3.89, IQR=.89) were not significantly different from those in group B (Mdn=4.00, IQR=.72); U = 114.5, p=.497. After removal of outliers, change in assessing dissatisfaction and readiness to change score distributions were not skewed. The independent samples t-test was performed to compare delta values between groups A and B. There was no significant difference in scores for group A (M=.25, SD=.39) and group B (M=.26, SD=.40); t(29)=-.127, p=.900. These results suggest that the study intervention had no impact on assessing dissatisfaction and readiness to change scores.

Multiple linear regression analysis was performed to evaluate for confounders with change in DES-LF assessing dissatisfaction and readiness for change scores (ΔDES-LF assessing dissatisfaction and readiness for change) as the dependent variable and potential confounders as independent variables modelled. It was found that none of the confounding variables significantly predicted change in DES-LF assessing dissatisfaction and readiness for change score (F[5,27] = .81, p = ns).

### Effectiveness: diabetes-related psychosocial self-efficacy (setting and achieving diabetes goals)

After removal of outliers, where setting and achieving diabetes goals was measured in completers (n=31), mean (SD) score improved by .16 (95% CI -.09−.41, p=.214) from 3.90 (.56) to 4.06 (.51). In group A completers (n=18), mean (SD) improvement was .10 (95% CI - .21−.41, p=.511) from 4.06 (.56) to 4.16 (.46). In group B completers (n=13), mean (SD) improvement was .23 (95% CI -.23−.69, p=.296) from 3.68 (.49) to 3.92 (.55).

The independent samples t-test was performed to compare change in setting and achieving diabetes goals score between groups A and B. There was no significant difference in delta values for group A (M=.10, SD=.63) and group B (M=.23, SD=.76); t(29)=-.526, p=.603. These results suggest that the study intervention had no impact on setting and achieving diabetes goals scores.

Multiple linear regression analysis was performed to evaluate for confounders with change in DES-LF setting and achieving diabetes goals scores (ΔDES-LF setting and achieving diabetes goals) as the dependent variable and potential confounders as independent variables modelled. It was found that none of the confounding variables significantly predicted change in DES-LF setting and achieving diabetes goals score (F[5,27] = .80, p = ns).

### Effectiveness: diabetes-related psychosocial self-efficacy (overall self-efficacy)

After removal of outliers, where overall self-efficacy was measured in completers (n=31), mean (SD) score improved by .25 (95% CI .09−.41, p=.004) from 3.78 (.46) to 4.03 (.44). In group A completers (n=18), mean (SD) improvement was .22 (95% CI .04−.40, p=.018) from 3.84 (.46) to 4.06 (.46). In group B completers (n=13), mean (SD) improvement was .29 (95% CI -.05−.63, p=.086) from 3.70 (4.7) to 3.99 (.44).

The independent samples t-test was performed to compare change in overall score between groups A and B. There was no significant difference in delta values for group A (M=.22, SD=.36) and group B (M=.29, SD=.56); t(29)=-.419, p=.678.

Multiple linear regression analysis was performed to evaluate for confounders with change in overall self-efficacy scores (ΔDES-LF overall self-efficacy) as the dependent variable and potential confounders as independent variables modelled. It was found that none of the confounding variables significantly predicted change in overall self-efficacy score (F[5,27] = .77, p = ns).

### Effectiveness: body mass index and blood pressure

Where BMI was measured in completers, (n=31, 62%), mean (SD) reduction in BMI was .4 (95% CI -.16-1.0, p=.151) from 31.0 (6.4) to 30.6 (6.3) kg/m^2^. In group A (n=18), mean BMI reduced by .3 (95% CI -.3-.7, p=.387) from 30.5 (6.0) to 30.2 (5.9). In group B (n=11), mean BMI reduced by .8 (95% CI -.7-2.3, p=.267) from 31.9 (7.1) to 31.1 (7.1).

Where blood pressure was measured in completers, (n=20), mean (SD) reduction in SBP was 9.2 (95% CI 3.0-15.3, p=.006) from 138.4 (19.1) to 129.3 (21.0) and mean reduction in DBP was 1.9 (95% CI -2.0-5.7, p=.325) from 79.0 (9.1) to 77.1 (9.0). In group A (n=12), mean reduction in SBP was 8.8 (95% CI .3-17.4, p=.044) from 137.4 (18.1) to 128.6 (23.9) and mean reduction in DBP was 2.5 (95% CI -1.8-6.8, p=.228) from 79.3 (9.1) to76.8 (10.4). In group B (n=8), mean reduction in SBP was 9.6 (95% CI -1.9-21.1, p=.090) from 139.9 (21.8) to 130.3 (17.3) and mean reduction in DBP was .9 (95% CI -7.8-9.6, p=.818) from 78.4 (9.8) to 77.5 (6.8).

A Summary of all quantitative results is presented in Table 6, with between group differences and p-values in Table 7.

**Table 6.**
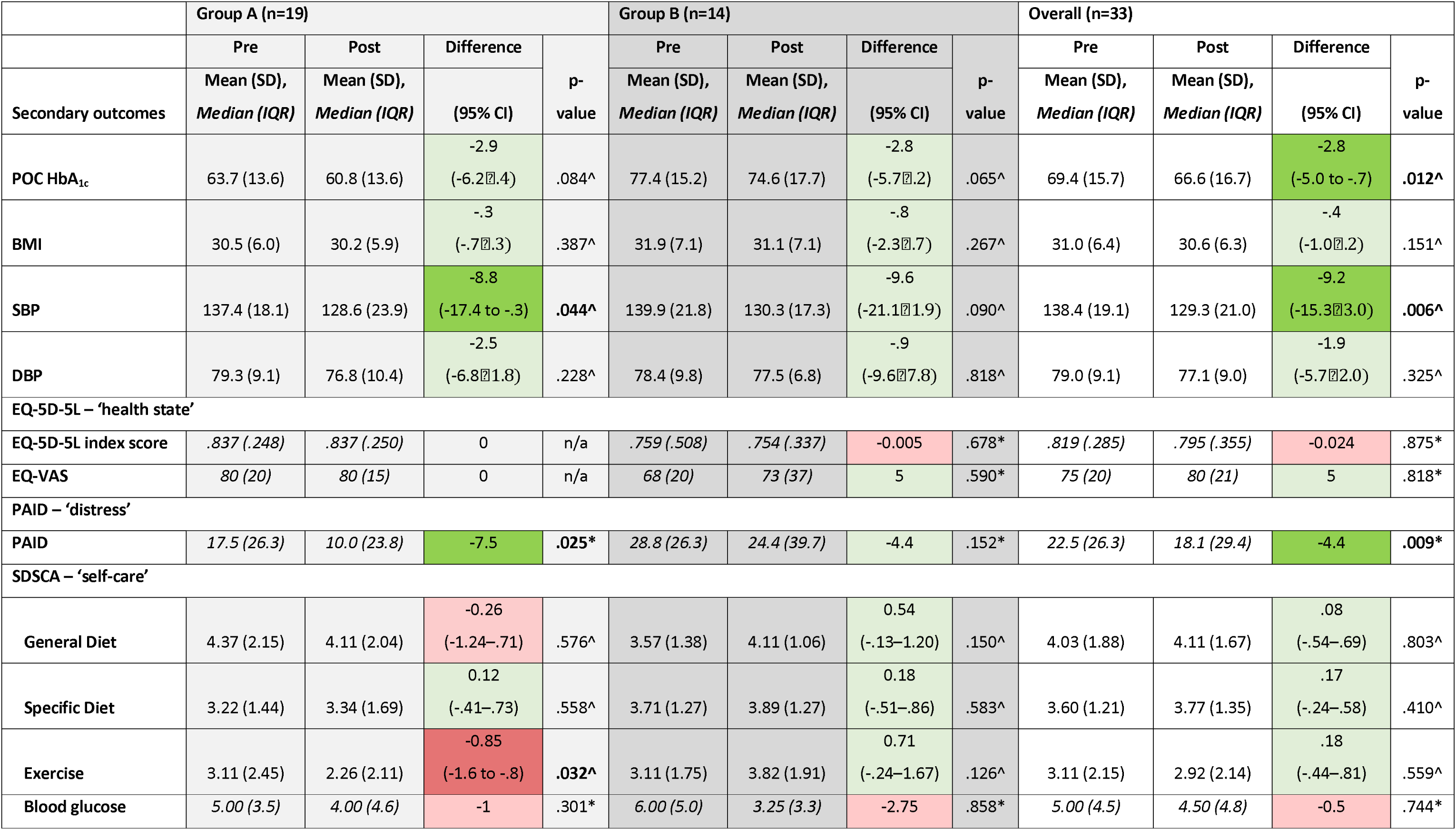
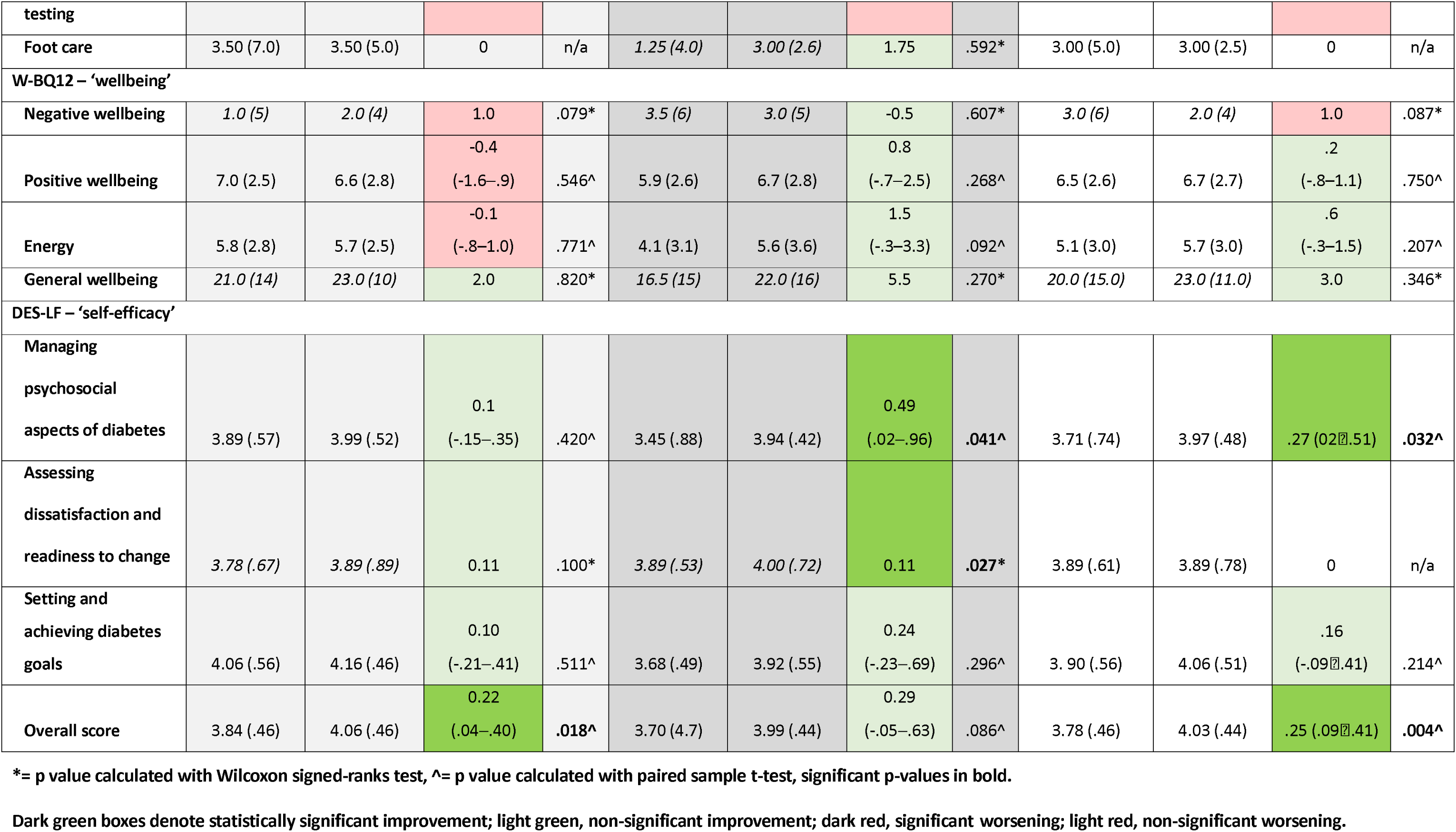
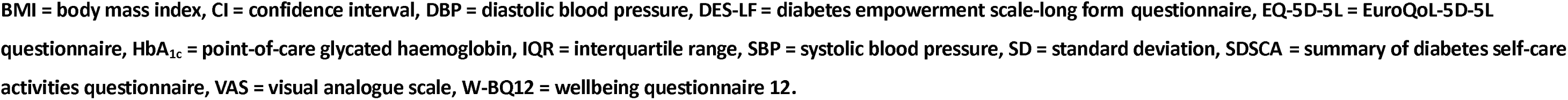
Results table demonstrating pre- and post-intervention scores for secondary outcome measures in the completers.

**Table 7.**
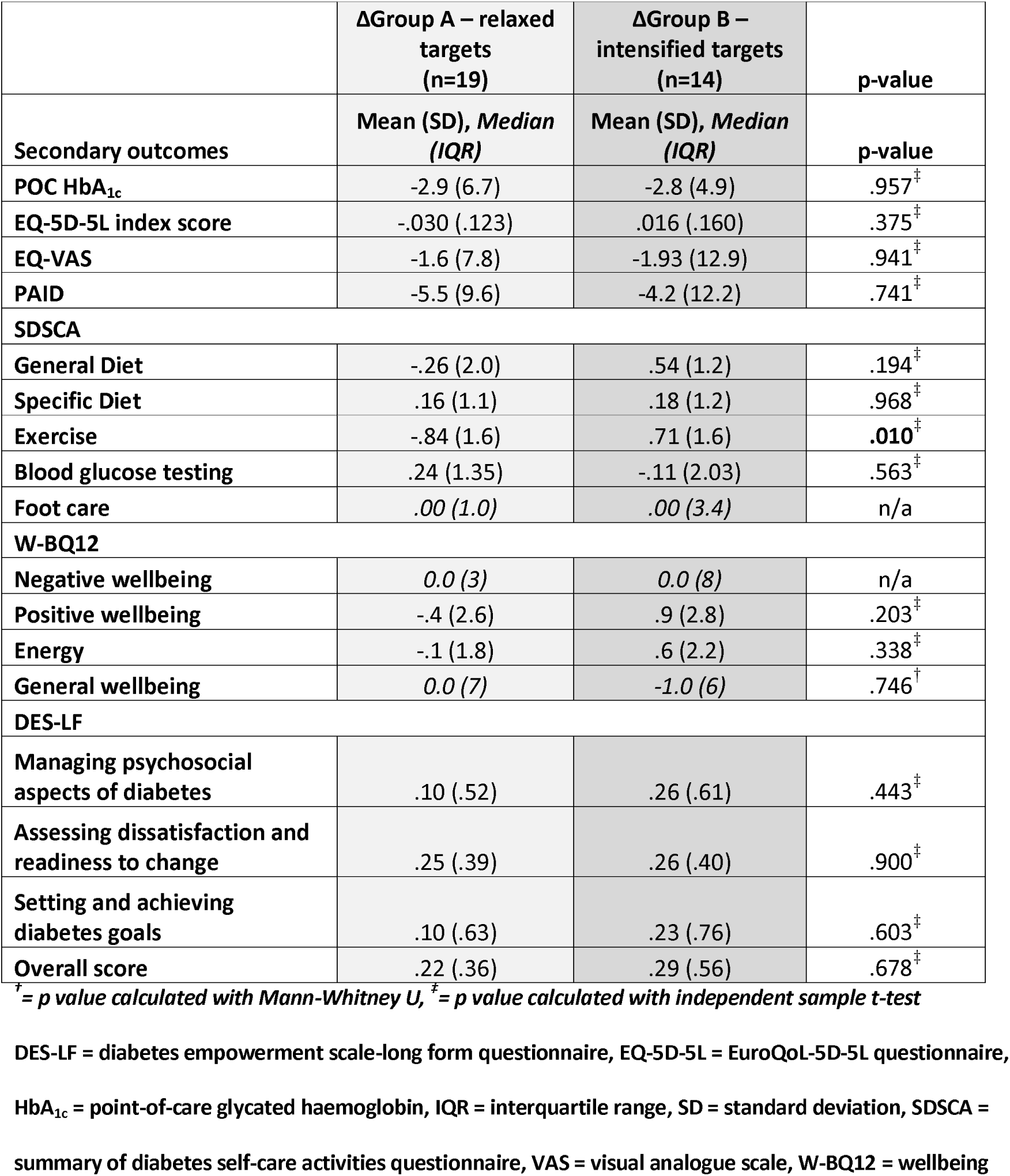
Results table demonstrating delta values for secondary outcome measures by intervention group.

### Confounders

Evaluation for confounding variables (gender, age, diabetes duration, IMD decile, BMI) with multiple linear regression stepwise modelling revealed only one out of twenty-five secondary outcomes listed in Table 7 were potentially affected by these confounders. For the change in SDSCA foot care score, multiple linear regression analysis resulted in a model suggesting BMI was a significant predictor of SDSCA foot care scores with an inverse relationship – as BMI increased, foot care activity decreased (β = -.60, t[32] = -3.82, p < .01).

### Semi-structured interviews: participant experiences

Semi-structured interviews were used to extract detailed narratives from participants on pre-determined topics of interest; perceptions of the use of individualised glycated haemoglobin targets, diabetes and wellbeing, and acceptability of study processes. All 50 participants in the feasibility study were eligible. A stratified, purposive sampling method was applied until the information power of the sample was deemed satisfactory by agreement between members of the research team (SJW, SW and GI). Fourteen participants were selected for interview.

Interview timing was scheduled in advance to ensure participants and the researcher were able to coordinate a distraction-free environment. All interviews took place via telephone and were recorded using a password-protected digital Dictaphone. Written, informed consent was obtained prior to interview commencement. Following introductions and confirming participant identity, an interview topic guide (Supporting information S4) was used to explore themes of interest and to standardise topics covered. As described in greater depth in the protocol, a semi-structured interview technique was used to provide deeper understanding of topics covered using clarification and follow-up questioning. Interviews were between 18 and 58 minutes in duration (Table 8).

**Table 8.** Characteristics of participants completing interview and free-text responses to an end-of-study acceptability survey, including baseline values from psychometric questionnaires.

| Participant number | Randomisation group (feasibility study) | Age (range) | Gender | Diabetes type | Diabetes duration (years) | IMD decile | Interview duration (minutes) | HbA<sub>1c</sub> (mmol/mol) | EQ-5D-5L INDEX | EQ-VAS | SDSCA general diet | SDSCA exercise | SDSCA blood glucose testing | PAID | W-BQ12 NWB | W-BQ12 PWB | W-BQ12 energy | W-BQ12 overall wellbeing | DES-LF psychosocial* | DES-LF dissatisfaction† | DES-LF goals‡ | DES-LF overall self-efficacy |
| --- | --- | --- | --- | --- | --- | --- | --- | --- | --- | --- | --- | --- | --- | --- | --- | --- | --- | --- | --- | --- | --- | --- |
| P1 | A | 70-74 | Male | Type 2 | 18 | 1 | 29 | 52 | 1.000 | 75 | 5.5 | 7 | 5.5 | 18.75 | 0.0 | 9.0 | 9.3 | 30.3 | 3.78 | 3.56 | 4.00 | 3.79 |
| P2 | A | 55-59 | Male | Type 2 | 14 | 1 | 24 | 62 | 0.750 | 70 | 3 | 3.5 | 7 | 60 | 5.0 | 3.0 | 4.0 | 14.0 | 3.44 | 3.44 | 3.50 | 3.46 |
| P3 | A | 45-49 | Male | Type 1 | 16 | 8 | 52 | 76 | 1.000 | 90 | 7 | 7 | 7 | 38.75 | 0.0 | 9.0 | 8.0 | 29.0 | 4.00 | 4.22 | 4.00 | 4.07 |
| P4 | A | 40-44 | Male | Type 1 | 0.12 | 3 | 44 | 43 | 1.000 | 85 | 4.5 | 0.5 | 3.5 | 15 | 0.0 | 5.0 | 7.0 | 24.0 | 4.00 | 3.33 | 3.60 | 3.64 |
| P5 | B | 60-64 | Female | Type 2 | 24 | 1 | 31 | 73 | 0.710 | 70 | 6 | 3 | 7 | 22.5 | 0.0 | 10.0 | 8.0 | 30.0 | 4.11 | 4.11 | 4.40 | 4.21 |
| P6 | A | 40-44 | Male | Type 1 | 14 | 8 | 24 | 65 | 0.795 | 70 | 0 | 4.5 | 7 | 26.25 | 0.0 | 11.0 | 7.0 | 30.0 | 4.44 | 4.11 | 3.70 | 4.07 |
| P7 | B | 45-49 | Female | Type 2 | 4 | 6 | 30 | 87 | 0.249 | 25 | 3.5 | 0 | 7 | 48.75 | 6.0 | 2.0 | 2.0 | 10.0 | 3.00 | 4.44 | 2.50 | 3.29 |
| P8 | B | 60-64 | Male | Type 2 | 10 | 1 | 19 | - | - | - | - | - | - | - | - | - | - | - | - | - | - | - |
| P9 | B | 20-24 | Male | Type 1 | 3.5 | 6 | 42 | 110 | 0.750 | 80 | 2 | 5 | 5 | 41.25 | 4.0 | 6.0 | 5.0 | 19.0 | 3.78 | 3.78 | 4.00 | 3.86 |
| P10 | A | 45-49 | Male | Type 2 | 15 | 1 | 34 | 58 | 0.768 | 75 | 6 | 3.5 | 4 | 31.25 | 4.0 | 5.0 | 3.0 | 16.0 | 3.56 | 4.11 | 3.70 | 3.79 |
| P11 | A | 20-24 | Male | Type 1 | 10 | 1 | 49 | 66 | 1.000 | 80 | 6.5 | 1.5 | 7 | 5 | 1.0 | 9.0 | 6.0 | 26.0 | 4.56 | 4.67 | 4.70 | 4.64 |
| P12 | A | 25-29 | Male | Type 1 | 13 | 3 | 58 | 76 | 0.721 | 70 | 0 | 0.5 | 5 | 28.75 | 6.0 | 5.0 | 4.0 | 15.0 | 3.89 | 4.11 | 3.50 | 3.82 |
| P13 | A | 20-24 | Male | Type 1 | 9 | 3 | 18 | 54 | 0.879 | 90 | 6 | 5.5 | 6.5 | 8.75 | 0.0 | 9.0 | 8.0 | 29.0 | 4.78 | 4.22 | 4.30 | 4.43 |
| P14 | B | 45-49 | Female | Type 2 | 6 | 1 | 42 | 53 | 0.768 | 65 | 2.5 | 0 | 2.5 | 22.5 | 1.0 | 4.0 | 2.0 | 17.0 | 3.11 | 3.89 | 3.60 | 3.54 |

DES-LF = diabetes empowerment scale – long form; EQ-5D-5L = EuroQoL-5D health related quality of life questionnaire; EQ-VAS = EuroQoL Visual Analogue Scale (overall health state); HbA<sub>1c</sub> = glycated haemoglobin; IMD = index of multiple deprivation; mmol/mol = millimoles per mole; NWB = negative wellbeing; PAID = problem areas in diabetes; PWB = positive wellbeing; SDSCA = summary of diabetes self-care activities; W-BQ12 = wellbeing questionnaire (12-item).
\* managing psychosocial aspects of diabetes
† assessing dissatisfaction and readiness for change
‡ setting and achieving diabetes goals
Cell results for PROs in each column are colour coded with conditional formatting rules, with better values being green, intermediate values yellow to orange, and worse values being red.

### Study feasibility

Alongside quantitative feasibility data, participant responses to interviews were used to evaluate study feasibility. Coding of transcripts considered five domains of interest: Acceptability, study-related documentation, questionnaire ease of completion, study-related time commitments and, due to high drop out rates, reasons for not dropping out.

#### Study acceptability

Many participants’ decisions to take part in the research were dependent on the nature of the study and the ability to discuss the study with researchers prior to consent. One participant commented:

> P9: “It’s a lot more helpful the fact that [recruitment is] human to human […] rather than just reading a piece of paper […] and then you can make your mind up if you want to participate or not.”—Type 1, male, group B

Rationale for the use of randomisation in the study was understood by participants. Participants further commented on study randomisation processes:

> P13: “Yeah, I think that’s a good way of choosing the groups because it will help show how A_1c_ targets affects wellbeing.”—Type 1, male, group A

Participants noted decisions to take part were influenced by their perceptions of the research; whether they felt the research was meaningful, and whether the research aims aligned with their values.

> P3: “I’m always a bit dubious about it [research] I needed to feel comfortable about it […] it would be [a] very strange person [not to be wary] […] it just depends if there’s a real value there or if you feel it’s just, you know, a box ticking exercise.”—Type 1, male, group A

#### Study documentation

Whilst many participants noted there was a lot of information to digest with the participant information documentation, some did not read the information prior to the study, instead relying on face-to-face interaction with the research to decide on consenting to take part.

> P6: “I’ll be completely honest; I hadn’t necessarily digested it… or read it [laughs]. I’d not really took it on board […] my initial reaction when I got the [invitation] letter was, “no, I don’t want to take part” is my open and honest opinion. But then when I was in the hospital and the consultant asked me to come and see you […] I think the thing that sold it to me was the conversation that we had.”—Type 1, male, group A

Even where not required, contact availability of the research team was deemed important by participants.

> P14: “I knew people would be on the end of the phone if I need to speak to anybody. It did make a really big difference.”—Type 2, female, group B

#### Study questionnaires

Baseline and endpoint PROs obtained using validated questionnaires informed preliminary outcomes of the impact of the study intervention. Some of the questions evaluated the impact diabetes had on participants’ psychological wellbeing. Despite this, participants did not feel they were inappropriately prying.

> P2: “There were none where I thought “ooo, you can’t ask that!”, didn’t bother me at all.”—Type 2, male, group A

> P14: “The questionnaires, they made you just stop and think. Well, how do I feel about that? Because you just feel it you don’t stop and think. You don’t recognise that you feel it.”—Type 2, female, group B

Some participants felt some of the questions were redundant and not applicable or poorly worded.

> P1: “I did find some of the questions hard to answer, insomuch as I felt some of the questions didn’t really apply [to me].”—Type 2, male, group A

#### Study time commitments

Time was often equated to value by participants. Research time commitments were acceptable if the participant felt there was either personal or general value in the research.

> P3: “[If] I felt there was value within it, I’d book a half day off—don’t worry about that—because I think it’s important.”—Type 1, male, group A

Maintaining flexibility around follow-up dates and times to fit in with participants’ life schedules was important in maximising retention over the 6-month study period.

> P10: “It wasn’t inconvenient for me at all, because obviously, [I was] communicating with you and you’ve managed to work around my work commitments, so it was no impact on me whatsoever.”—Type 2, male, group A

With higher-than-expected dropout seen in the feasibility study, participants were asked what aspects of the research encouraged them to remain in the study and attend study visits.

Having a particular interest in the topics covered in the research enhanced motivation to take part.

> P11: “I was quite motivated to stay on because I think I really care about this sort of wellbeing aspect of diabetes.”—Type 1, male, group A

A transactional relationship between participant and researcher was described by other individuals who completed the feasibility study. A two-way relationship was described, with participants feeling useful by being part of research, but also recognising that the research may confer benefit to them.

> P14: “I felt very useful to be honest […] I know, it sounds corny, but ‘giving something back’, because I was getting all this help from you and from the hospital. I was getting the full MOT.”—Type 2, female, group B

Additionally, some participants perceived an improvement in their mood by taking part in the research, though it was not clear from the results which aspect of the research improved mood.

> P7: “It’s a benefit to me, as well as to you. So, I suppose for me, I get to try and lower my HbA_1c_ level and be given that aim […] I think, feeling something positive about having diabetes, you know that you can do something positive!”—Type 2, female, group B

### Study intervention

Four codes were derived from the analysis in this category: motivators, demotivators, individualisation, and knowledge/opinions/experiences.

#### Motivations for achieving HbA_1c_ targets

Participants described factors motivating them towards achieving their HbA_1c_ target, whether in the study or in general. Positive motivators were generally goal-oriented. Further positive points illustrated a more nuanced picture, demonstrating even small improvements on the way towards an individual’s goal were perceived as motivating. In many participants, being assigned a glycated haemoglobin target, whether they were randomised to relaxed or intensified targets, resulted in improvements in PROs irrespective of whether glycaemic control improved or whether they achieved their study target.

> P5: “I think it’s really a good idea to have a target because I feel that if you went to your follow-up so many months later and they said, “well, you’re near that target now” I think it would give you more incentive to keep going.”—Type 2, female, group B

> P6: “It would be even better if you could see how your A_1c_ changes over time and really take time to celebrate achievements, even if you’re not at the level you should be.”—Type 1, male, group A

Other motivators were directed towards participants’ desire to avoid negative situations associated with their diabetes, such as avoidance of hypoglycaemia or diabetes complications. These goals felt more personal, with specific experiences contributing to participants’ responses. Additionally, negative motivators could sometimes be interpreted to stray away from the perceived wisdom of ‘lower (HbA_1c_) is better’.

> P3: “It just worries me that I need to stay at a certain point [blood sugar, HbA_1c_]. Because if I don’t catch it the right time…[pauses] I’m gonna die basically.”—Type 1, male, group A

#### Demotivators for achieving HbA_1c_ targets

Similarly to motivators, demotivators were often differentiated by participants’ experiences. Frustrations and lack of understanding were barriers to participants pursuing their HbA_1c_ targets.

> P5: “[when my HbA_1c_ didn’t improve] I think it used to make me feel, “why bother?””—Type 2, female, group B

Historical paternalistic healthcare interactions brought negative connotations to glycated haemoglobin levels with patients.

> P6: “I was getting told off every time I [my HbA_1c_] was too high [laughs].”—Type 1, male, group A

Lack of adequate education around diabetes meant participants lacked the appropriate knowledge to incentivise glycated haemoglobin target achievement.

> P10: “A doctor there started giving me a hard time about me not taking my diabetes seriously and I was saying, “Well, I have started taking it seriously” and he said, “well, your HbA_1c_ is really high.” and I didn’t really know what that was.”—Type 2, male, group A

The knowledge requirements to effectively self-manage diabetes are significant. Participants often reported information overload in relation to the requirements of monitoring and managing their diabetes.

> P9: “It did seem like, “oh, look… another thing I’ve got to measure, another thing I’ve got to manage” […] it’s just another fancy labelled medical thing that I’ve got to look at!”—Type 1, male, group B

Other individuals were more singularly focussed on the rift between their current glycaemic control and the target assigned. Having goals which were not achievable was negatively perceived; either causing feelings of obsessive behaviour or apathy and defeat.

> P13: “I think if the goals aren’t achievable, you can become obsessive.”— Type 1, male, group A

> P14: “If it isn’t achievable and I think I’m not going to get that then, you know, I’m not even going to try.”—Type 2, female, group B

In those with type 2 diabetes, where prior improvements or successes were noted, or where diabetes was not perceived to be impacting upon their health, participants were less focussed on achieving their target.

> P7: “You don’t have symptoms, so you just think “oh, well, it’s fine.””— Type 2, female, group B

#### Use of individualised HbA_1c_ targets

Use of individualised glycated haemoglobin targets based on patient characteristics is common practice for diabetes healthcare professionals. Whilst many patients understand what HbA_1c_ is, many remain unaware of their target or that it may be individualised to them.

Participants were noted to aim for glycated haemoglobin levels that catered to their own current social or functional goals rather than biomedical goals.

> P3: “That [HbA_1c_] might sound very high. But it’s not high for me […] allow me to be happy with the risk on the higher side. […] I think transparency is everything.”—Type 1, male, group A

Noting that unrealistic or unachievable glycated haemoglobin targets were demotivating for participants, with the proviso that targets were achievable, participants were in favour of individualised targets. Achievability was not defined in a standardised way by participants, highlighting what was achievable by some may not be by others.

> P6: “I think it’s good to have a target for something to aim for—it’s how manageable that target is [that’s important] and whether you are able to manage it on a day-to-day basis […] it’s what is a manageable target that can be done. […] I think having a target has to be [compatible with] your lifestyle.”—Type 1, male, group A

Participants, especially those with type 1 diabetes, reported good awareness of risk stratification in relation to individualised glycated haemoglobin targets. Despite the study intervention and achieving their study HbA_1c_ target, self-efficacy and the subscale of DES-LF, ‘setting and achieving diabetes goals’ did not always correlate with the views expressed by interviewees.

> P10: “I can only say from my perspective; it works for me that everybody has individual goal. It’d be crazy, I think, to say that you’ve all got the same targets. You’ve got to cater for the individual.”—Type 2, male, group A

#### Participant knowledge and experiences with the use of HbA_1c_ in diabetes

Levels of knowledge on the utility of HbA_1c_ targets to guide diabetes management and stratify risk of complications were varied among participants. Anecdotally, participants with type 1 diabetes appeared to have a greater understanding of HbA_1c_.

> P3: “[It’s] how I’m looking after myself basically […] so that you can monitor any changes […] so that [pauses] …you get to know me—what [blood sugar] I’m happy with.”—Type 1, male, group A

Participants desired more information regarding glycated haemoglobin and targets, noting they felt better when knowing the risks associated with sub-optimal HbA_1c_ values.

> P7: “I think educating myself helped me to understand it better and talking to other people […] when I’m more informed, I feel better.”—Type 2, female, group B

Good awareness of glycated haemoglobin levels in individuals could cause frustrations in those who, despite efforts, were not achieving their targets. Knowledge of glycated haemoglobin and the long-term risks associated with chronic hyperglycaemia alone was not sufficient in enabling individuals to achieve their targets but did improve distress and self-efficacy.

> P5: “I constantly [pauses] …constantly thought about it, but I didn’t quite know—because they were always telling me it’s still high—where I was going wrong, really. Because I did… I tried [to get the HbA_1c_ down] all the time, you know?”—Type 2, female, group B

Despite worsened glycated haemoglobin levels and failing to meet the study targets, some participants were comfortable with a target they felt was appropriate for them, rather than an externally imposed target.

> P1: “I think that I’m at a reasonable [HbA_1c_] level for me. […] you know, I think to get to the stage I’m at with my target, I’ve been realistic.”—Type 2, male, group A

Information overload and lack of good understanding of glycated haemoglobin were mentioned as persistent issues prior to study entry amongst those interviewed.

> P5: “I’m trying to think of everything that they taught me in that appointment. And I worry and think “have I remembered everything?””— Type 2, female, group B

### Diabetes and wellbeing – psychological aspects

Low levels of wellbeing, self-efficacy, and self-care alongside high levels of diabetes-related distress were seen at baseline in the feasibility study participants reported so far. Sub-themes related to psychological aspects and physical aspects are used to explore this topic.

Participant responses in this section help contextualise and triangulate the quantitative data, aiding interpretation of the results and enhancing understanding.

Two codes were used in this sub-theme to explore participants’ mental health experiences with diabetes: positives and negatives. Negative psychological aspects of diabetes coded in transcripts were further divided into the following subcodes: burden, coping mechanisms, denial, and guilt and failure.

#### Positive psychological effects of diabetes

The impact of diabetes on participants was not always negative. Participants, particularly those who had type 1 diabetes from a young age, noted how having diabetes had brought organisation to other aspects of their life. A diligent approach to daily life is often required of people with type 1 diabetes – self-management activities span from monitoring and responding to blood glucose levels, to attending multiple annual hospital appointments, requiring significant mental effort and planning. Some of the skills required in the self-management of diabetes were seen to be transferrable and useful in other aspects in life, as elaborated upon by participant P3 below.

> P3: “I think diabetes has probably made me more process-driven, if you like, because I’ve got to be methodical… with my diet, with my checking my blood, with eating on time, etc.”—Type 1, male, group A

The modality of blood glucose monitoring, specifically in people with type 1 diabetes using intermittently scanned continuous glucose monitors (isCGM), was noted to have positive benefits. Despite improvements in glycated haemoglobin levels in the study, meeting the study target and improvements in self-efficacy, similar improvements were not always seen in health state, wellbeing and distress following the study intervention. Despite this, the benefits of access to diabetes technology were positively commented on frequently by people with type 1 diabetes. Individuals discussed personal benefits and the enhanced visibility and wider knowledge to the public that use of wearable diabetes technology has brought.

> P3: “I enjoy life! I don’t let diabetes rule it […] It doesn’t ruin your life. It’s just that you’ve got to learn to cope with [it]. […] Libre has given me another lease life.”—Type 1, male, group A

Participants acknowledged that acquiring a diagnosis of diabetes improved health awareness and triggered positive health-improving behaviours such as improved levels of exercise and healthy eating.

> P13: “Having diabetes helped me lose the weight and by motivating me to get into the gym and eat healthy and stuff.”—Type 1, male, group A

The importance of any positive aspects of diabetes, including achieving personal diabetes goals or improving one’s health cannot be overstated. As shown in interviews with healthcare professionals discussed later, highlighting the importance of any positive outcomes for people with diabetes is critical in consultations to improve motivation and sense of achievement.

#### Negative psychological aspects of diabetes

The life-long nature of diabetes was noted to take a toll on some individuals, especially where medication side-effects were felt to be troublesome or where improvements in diabetes were not achieved. Trends in quantitative outcomes discussed earlier indicate general improvements in distress, wellbeing and self-efficacy three months post-intervention. Data at an individual level demonstrates a considerable range of positive and negative outcomes between participants.

> P3: “I’ll be quite black and white about it: Diabetes, completely ruined my life […] about 18 months ago, I was probably the worst I’ve ever been: really, really worried about… about diabetes and how it would… I could die from it, basically.”—Type 1, male, group A

In the initial stages of diagnosis, participants noted negative impacts on their mood, often exhibiting characteristics associated with the Kübler-Ross [49] stages of grief, though not necessarily transitioning through these stages in the traditional linear fashion.

> P7: “I suppose I was quite down about it in the beginning. But I’ve kind of learnt to think, well, you know, you’ve got to learn to live with it, it’s not something you can just ignore.”—Type 2, female, group B

> P9: “I think it was an initial shock when I got home. Then I was like, you know, “everything’s gonna be fine.” And then the moment I actually went to bed, I started, you know, just sitting there mulling it over, I then just started to bawl.”— Type 1, male, group B

Aside from side-effects and lack of improvements seen in biomedical metrics such as HbA_1c_, day-to-day impacts of diabetes on general physical wellbeing were also noted by participants to impact mood.

> P14: “It lowers your mood because you feel so ‘ugh’ and so tired. You get fed up with yourself, get fed up with feeling the way you’re feeling. […] It can massively lower your mood.”—Type 2, female, group B

#### The psychological burden of diabetes

The psychological burden of diabetes can mimic signs of anxiety and depression. Diabetes was likened to a weight individuals carry around, reflecting a condition that can require a significant allocation of mental effort to maintain self-management. Others noted the burden of treatments for diabetes.

> P12: “My diabetes is like a backpack—it just sits on my back.”—Type 1, male, group A

Participants noted not only the burden of diabetes on themselves but the perceived burden they placed on others around them.

> P3: “You do feel like a burden to people. And I think that’s where you get your worries from is—is it [my diabetes] affecting other people […] [it] makes you feel down sometimes because like, you stew over these things; you think you are a burden. You are a burden to people.”—Type 1, male, group A

In those with type 1 diabetes, the sudden and unexpected nature of the diagnosis can be daunting for individuals, often taking time to process. For many, the recognition of the lifelong requirements of managing diabetes can take some time to realise.

> P4: “This is something I need to do now for the <u>rest of my life</u>.”—Type 1, male, group A

#### Comping mechanisms in diabetes

Psychosocial factors influenced the coping mechanisms used by participants, often impacting upon individuals’ self-management abilities and subsequent glycaemic control. Whilst recognising some coping mechanisms were unhealthy, motivations behind certain behaviours were governed by life-circumstances.

> P9: “It’s another reason why I always have my blood [sugars] so high. I would rather have it higher and deal with the ketones than it be low and me be at a chance of losing consciousness. […] I live alone, you know, nobody would know anything was up until the day later when I don’t show up for work.”—Type 1, male, group B

Coping mechanisms were also related to avoidance of unpleasant experiences such as blame or insulin injection. Anxiety around insulin injection resulted in skipping insulin injections or avoiding food in the hopes of obviating the need for injection, despite the risks.

> P9: “I remember one time when I was young, I refused to eat anything because I would have to take an injection.”—Type 1, male, group B

#### Denial in diabetes

Denial in diabetes was common amongst participants and often self-recognised in retrospect. Reinforcing differences in the mentality of people with type 1 or type 2 diabetes, those with type 2 cite the lack of symptoms as contributory to low levels of self-management and underestimating disease severity.

> P5: “Yes… It didn’t feel real! If… If you know what I mean? I thought, “Oh I’m fine… I feel well.”… Yeah… [sounds unsure] […] there’s a temptation to think it isn’t really serious; but it is obviously, but you don’t feel it is. Because you’re not feeling ill and because you’re not seeing anybody… medically. You feel it’s not real, you know, which is daft.”—Type 2, female, group B

Normalising high blood sugar levels due to lack of symptoms also occurred in those with type 1.

> P12: “I felt <u>fine</u>. I didn’t feel unwell. I knew I had a high blood sugar. It wasn’t horrible… It just didn’t register.”—Type 1, male, group A

#### Guilt and failure

Diabetes, whether type 1 or type 2 tends to be progressive and get worse by itself over time— whether from progressive beta-cell destruction in type 1 diabetes or progressive insulin resistance and beta-cell loss in type 2. Despite the natural progressive nature of diabetes, many participants felt personally responsible for deterioration or lack of improvement in their metabolic control.

> P9: “It really does… and I am using this word accurately… [long pause] …it does <u>hurt</u>. To sort of see that the effort that you’ve put in it just immediately gets sort of thrown away because of that A_1c_ reading… because I wasn’t trying hard enough. So, I have to try harder still.”—Type 1, male, group B

Participants with type 2 diabetes found solace in knowing that they were not responsible for their diagnosis, with two major contributors—genetics and age—being uncontrollable.

> P5: “It would have been helpful to see other people [with diabetes] to… to be reassured, you know, this isn’t something that you’ve brought on yourself.”—Type 2, female, group B

Historical negative perceptions of being ‘told off’ at diabetes appointments still endure in those with longer diabetes duration.

> P5: “When it gets to the time when I’ve got to go and see somebody—the diabetic nurse, or the team—I used to start panicking. I’m thinking, [whispers] “Oh God!”, because I didn’t test my blood as often as I should… And if it was high, I’d think, “Oh God”, you know…I’m going to face the wrath.”—Type 2, female, group B

Being a long-term condition, many issues can crop up over the lifetime of individuals with a long duration of diabetes. Some issues will be related to their diabetes diagnosis, whilst others will be incidental. Frustration at the chronicity of the condition alongside multiple related and unrelated health concerns that can occur was difficult for some.

> P3: “It sets you on that downward spiral of: “Okay, what’s next? What else is going to be blamed on diabetes? Am I ever going to win the battle?””— Type 1, male, group A

Individuals may have contributed to their own increased risk of diabetes to some extent but often there are many external factors outside of their control which contribute to their risk. Despite this, participants—specifically those with type 2—often felt personally responsible for their diagnosis.

> P14: “Because I have done this to myself. I’ll fully hold the hands up and say it was me. I did it. I ate all that rubbish. I didn’t look after myself.”— Type 2, female, group B

### Diabetes and wellbeing – physical aspects

Three codes were used in this sub-theme to explore the interaction between physical and mental health in diabetes: blood sugars, complications, and side-effects. Covid-19 was added as an additional code during the interviews as the pandemic was noted to impact on people with diabetes in unique ways.

#### Blood sugars

Daily variations in blood glucose readings impacted participants in different ways. Good blood glucose readings were noted to positively impact participants’ sense of physical wellbeing.

> P2: “When my blood [sugars] are right down, I feel absolutely fine. I have more energy and I’m not as tired.”—Type 2, male, group A

Conversely, problematic or erratic blood glucose readings, whether too high or too low were negatively perceived and caused moderate distress, despite high blood glucose-testing self-care scores.

> P5: “I was tormented; my bloods [sugars] were everywhere. They were going high, and I was having hypos every couple of days at night […] I was a bit nervous to go to bed, to be honest, because luckily, I’m aware, I wake up and I’m aware of [the hypo].”—Type 2, female, group B

Monitoring of blood glucose was generally negatively perceived. The relatively recent advent of isCGM has helped participants with anxiety around finger-pricking. Participants’ self-confidence and self-efficacy directly impacted their ability to undertake blood glucose self-monitoring.

> P3: “A lot of my anxiety did come from—in the latter years—is the finger pricking […] Libre has given me another lease life.”—Type 1, male, group A

#### The complications of diabetes

Treating and managing the complications of diabetes requires a significant amount of healthcare resources. The avoidance of diabetes complications by modifying patient risk factors, such as glycated haemoglobin levels, is therefore central to healthcare interventions in diabetes. Participants were acutely aware of the risks of future complications which was often seen as a motivator for improved self-management.

> P1: “I am aware [of the complications of diabetes] so because I’m aware, I have to look after myself now.”—Type 2, male, group A

The initial development of complications may go unnoticed by participants and is often picked up on annual screening. For most, the development of diabetes complications is almost inevitable, even with optimal glycated haemoglobin levels. Complications can often come as a shock for individuals and have an impact on mood.

> P2: “When you read about it, what it can do to you and everything, it’s just like, “oh my god!”, you just don’t realise [what diabetes can do] and you think, “how long have I got?”, type of thing. I was just, like, <u>depressed</u>.”— Type 2, male, group A

#### Treatment side effects

Whether the absolute life-sustaining requirement for insulin injections in those with type 1 or the spectrum of tablets and injections used in type 2 diabetes, the most common side-effect is hypoglycaemia. The physical effects of hypoglycaemia can be dramatic and cause health-damaging avoidance of low blood sugars by maintaining glucose levels at the opposite extreme.

> P9: “The thing that bothers me most is something that scares me in general which is just the vulnerability. The human body can just do wonderful things just by itself but with diabetes, God forbid I go without a Mars bar for like three hours and I suddenly start hypoing and then I’m suddenly put in this very vulnerable state. That’s the thing that scares me about diabetes the most.”—Type 1, male, group B

Alongside physical effects, participants acknowledge the psychological effects can also require significant mental resources.

> P11: “The risk of hypoglycaemia is something I’m thinking about every single shift.”—Type 1, male, group A

For some, despite not experiencing the worst level of hypoglycaemia (severe hypoglycaemia), avoidance of hypoglycaemia was a significant source of anxiety and fear.

> P3: “All of a sudden, when you have a few… I wouldn’t even call them hypos… I’ve never really had an extremely bad one. […] But I do feel like it’s crept up and crept and crept up over the years. […] I put my anxiety down to having hypo and dying from it.”—Type 1, male, group A

#### Covid-19 and diabetes

Added as an additional code during the interview processes, the coronavirus pandemic was brought up by several participants without prompt. During the recruitment and data collection periods, national guidance on mask-wearing, self-isolation, shielding, travel and working from home was frequently changing. With higher mortality rates seen in people with diabetes, participants received shielding letters in efforts to prevent the spreading virus negatively impacting those at increased risk of adverse outcomes. From a healthcare perspective, it made sense. At an individual level, participants often experienced even higher levels of anxiety.

> P3: “Receiving letters and the like from government. Having to go on the gov.uk websites to register the fact that you are at an undue risk of dying, et cetera… it’s a lot to accept […] because the first message that was thrown out is that diabetics were suffering much worse than… than others in this pandemic.”—Type 1, male, group A

For many individuals shielding, Covid-19 lockdowns were an isolating experience, potentially reducing or eliminating the usual support networks present.

> P4: “It is scary because it’s not just being unwell, but through Covid, you can’t have any visitors, you just feel you’re on your own.”—Type 1, male, group A

#### Advice to others

With feedback posted to participants on their qualitative outcomes, additional experiences were sought from qualitative interviews to add greater interest to the feedback provided. Interviewees were asked what piece of advice they would provide to their younger self, or someone else struggling with diabetes. Discussions on this topic with participants were varied but often focused on the avoidance of negative experiences they had with diabetes.

Psychological aspects of diabetes were felt to be as important as physical aspects of self-management.

> P9: “Make sure that you yourself are okay, both physically but more important mentally, because you don’t end up doing the medication and look after yourself if you’re not in the right mindset to do it.”—Type 1, male, group B

Participants stated difficulty in acknowledging the diagnosis as they were not experiencing negative symptoms, but reinforced advice to accept the diagnosis earlier before the damage associated with high blood sugars was established.

> P5: “There’s a temptation to think it isn’t really serious; but it is obviously, but you don’t feel it is… Because you’re not feeling ill and because you’re not seeing anybody…medically. You feel it’s not real, you know, which is daft […] it is real and you’ve got to do it.”—Type 2, female, group B

Other advice from participants was focused on enhancing positivity and acceptance.

> P11: “Don’t let the idea or the thought of, “Oh, I have diabetes!”, overwhelm you as a person and make you feel unworthy or unable to live life as you would like, or to aim for those targets that you’ve been given.”— Type 1, male, group A

### Semi-structured interviews: healthcare professional experiences

In total, eight healthcare professionals were recruited for interview, following the interview topic guide (Supporting information S5) to explore themes of interest and to standardise topics covered. Baseline characteristics are noted in Table 9. Participants were recruited from a range of professional backgrounds directly involved in the care of people with diabetes.

**Table 9.** Participant characteristics.

| Participant number | Role | Band | Interview duration |
| --- | --- | --- | --- |
| H1 | Specialist Diabetes Dietitian | 6 | 35 |
| H2 | Diabetes Specialist Nurse | 7 | 30 |
| H3 | Research Nurse | 6 | 52 |
| H4 | Diabetes Specialist Nurse | 7 | 27 |
| H5 | Consultant Physician | N/A | 24 |
| H6 | Specialist Diabetes Dietitian | 7 | 45 |
| H7 | Diabetes Specialist Nurse | 7 | 17 |
| H8 | Specialist Trainee (doctor) | ST5 | 36 |

Three participants were diabetes specialist nurses (DSNs), two were specialist diabetes dietitians, two were doctors and one a research nurse. To maintain participant confidentiality, participants were assigned a unique identifying code (Table 9).

Healthcare professionals’ engagement with the interview process was good, with participants describing their views and experiences on the perceived positive and negative impacts of glycated haemoglobin targets and levels on people with diabetes in a straightforward and articulate manner. Additionally, participants noted the profound impact diabetes has on the mental and physical wellbeing of their patients. Many of the findings demonstrated a good level of consistency with the patient interviews, indicating healthcare professionals’ perceptions of the views and experiences of people with diabetes concerning their wellbeing and use of glycated haemoglobin in diabetes were accurate.

When the interview data were analysed, two major themes were identified:

1. Diabetes management.
2. Diabetes and wellbeing.

Sub-themes were used to further categorise the data, with deductively and inductively derived codes representing distinct topics of interest within each theme.

### Managing diabetes – treatment targets

Six codes were used in this sub-theme to explore healthcare professionals’ perceptions of the impact glycated haemoglobin targets have on their patients: achievability, advantages and motivators, disadvantages and demotivators, engagement, individualisation, and patient understanding.

#### Achievability

Echoing the experiences of patients reported earlier, healthcare professionals noted the importance of HbA_1c_ targets being achievable in motivating patients to improve their self-management and strive towards their target, mentioning the importance of informing patients of the rationale for the usage of HbA_1c_ monitoring and targets as part of diabetes management.

> H3: “It’s always like something [to aim for] and if they know the reasonings behind why those targets are actually set, then I feel that it’s a good thing. But I can see why people do get a bit demoralised. And like also say, “well, if I’m never going to achieve it, why do I bother?” But it’s hard, isn’t it?”

Use of small aims on the way towards larger achievements was a strategy used by some professionals to maintain motivation alongside only using HbA_1c_ targets as a motivator for improved self-management when their patient is close to target.

> H4: “If I can see they were struggling, I would just maybe focus on the smaller aims and achievements. […] I would aim to be putting the prompts of HbA_1c_ targets for people that aren’t a million miles off their target.”

Intensive glycated haemoglobin targets are notably difficult for patients to achieve with professionals noting safety concerns, such as excess risk of hypoglycaemia, for some patients attempting to reach more intensive targets. Many individuals never reach the standard target range used to monitor diabetes in primary care in the UK.

> H8: “An A_1c_ of less than 48 [mmol/mol] is not always achievable […] It’s just trying to get balance, really. […] I do always kind of emphasise and say that’s not achievable, or it’s not safely achievable for everyone.”

#### Advantages and motivators

Patient experiences noted several motivating factors for improved self-management, including reducing the risk of future complications and accommodating specific life circumstances, such as the need for surgery to go ahead. Professionals acknowledged similar views to their patients, demonstrating a good level of understanding of patient motivators.

> H1: “Some of them are very driven with their A_1c_ targets, but that is usually to get surgery.”

With the findings so far noting that PROs of diabetes-related distress and self-efficacy improve irrespective of the target set—suggesting that the process of being given a target rather than the target itself improved these metrics—professionals viewed specific HbA_1c_ targets as motivators for individuals, with patients of their who had received a target often looking forwards to repeat HbA_1c_ measurements to track progress.

> H2: “I think it probably does motivate [patients] if they’ve got kind of a target to aim towards. […] They do look forward to knowing what the result is, especially if they’ve worked hard.”

Where professionals note improvements, they would integrate HbA_1c_ results into consultations to maintain motivation, especially where patients were noted to be close to achieving their target.

> H4: “You’ve got to focus on the positives of why they’re here. So, it could be on an A_1c_ result. […] It’s trying to focus on the positives of the consultation and then integrating the A_1c_ in that way.”

#### Disadvantages and demotivators

Further demonstrating the deep understanding they had of their patients, professionals were highly aware of circumstances which could be demotivating for people with diabetes.

> H1: “If it’s not come down or it’s stayed the same, or they’re aware that nothing’s really changed, there’s… [pauses] there’s disappointment. […] But I feel that that’s kind of like constant disappointment with them really.”

Whether in the initial stages of diagnosis or over the course of years of intensive self-management, the burden of diabetes was noted to be overwhelming for patients and was a source of frustration.

> H2: “Patients can get very overwhelmed with the diabetes. They often say they get fed up when they see high [HbA_1c_/blood sugar] numbers […] it is really frustrating when you’re doing all <u>you</u> think you can plus the patient’s doing everything they can and it’s still not budging.”

Professionals avoided discussions around HbA_1c_ levels and targets if patients had poor glycaemic control or were far from their target.

> H5: “If someone is very far from the [HbA_1c_] target, to motivate them to come back, I don’t talk about HbA_1c_, most of the time… [pauses] they just get demotivated more than anything else.”

#### Engagement

Life circumstances or denial remain consistent barriers to engagement with self-management and with healthcare services.

> H4: “I guess it gets to a point where some people just aren’t ready. […] elements of denial, maybe in some respects, you know, it’s just frustration. […] [it might be the] work situation, they might be away at uni, might be on holiday, they might just have issues coming into clinic, you know, financially, […] [or are] carers for family members. So, I think there’s probably multiple issues why people don’t always engage.”

The patient-healthcare professional relationship was highlighted as a key element for enhancing patient engagement and trust.

> H5: “The relationship between the DSN, the doctor and the patient, it just makes a huge difference. They have to trust you. And they have to feel confident ask any question that they have.”

#### Glycaemic target individualisation

Professionals highlighted a lack of consistent understanding amongst people with diabetes of the use of glycated haemoglobin targets to balance the risk of future complications against the risk of intensive glycaemic control, agreeing with the findings of interviews with patients. Patients were often noted to be unaware that glycated haemoglobin targets were individualised based on their characteristics and so may understandably lack the motivation to achieve them.

> H1: “People don’t know that it’s [HbA_1c_ targets] tailored towards them, they probably just see that as a number rather than the decisions made around it. […] I feel as though the main point of that conversation is the significance of that target and what that’s actually measuring.”

A recurrent theme that occurred on review of the interviews with participants and in interviews with professionals, was the need to individualise targets not only based on patient characteristics but also considering specific life circumstances.

> H3: “Some people will accept it, you know, and are quite happy. They will say, “yeah, that’s fine, we’ll work towards that target [HbA_1c_], that that’s not a problem.” Whereas other people are like, “Well, no, because I won’t be able to drive” or, “I’ll end up going hypo at this time” and, you know, I feel like they know their own body, which is most probably true in some respects.”

#### Patient understanding

There was a perception among professionals that patients prefer the immediacy of blood glucose readings over the long-term picture provided by glycated haemoglobin. Interviews with participants were less binary, suggesting that where day-to-day self-monitored readings were erratic in someone having issues with self-management, blood glucose measurement could be a significant source of anxiety.

> H1: “[blood sugars] are more relatable to them, and they can visually see it rather than waiting three months.”

### Managing diabetes – diabetes care

Five codes were used in this sub-theme to explore healthcare professionals’ experiences managing people with diabetes: practice, healthcare professional knowledge, management differences between type 1 and type 2 diabetes, patient autonomy and young adults.

#### Current practice

Beyond what is learned as part of a university course or diploma, professionals discussed additional facets of their practice that they had acquired over many years of caring for people with diabetes. The frequency of psychological issues occurring in diabetes consultations was mentioned by several professionals. A lack of adequate training in the management of psychological issues within consultations was noted.

> H1: “With being untrained and you’re kind of almost using psychology methods […] It is about picking up on the right things that they say.”

During patient interviews, the initial stages of diabetes management soon after diagnosis were noted to be overwhelming for people with diabetes. To help give meaning to the concept of glycated haemoglobin, professionals often related HbA_1c_ targets to corresponding day-to-day blood glucose readings to aim for.

> H2: “I would always say to them, you know, “I’m going to adjust your target”, then I usually say to them, “this is what this [HbA_1c_] corresponds to in blood glucose levels.”, so they have a rough idea of what blood sugar levels range, they should [be aiming for].”

#### Healthcare professional knowledge

With an increased prevalence of mental health issues seen when compared with the general population, presence of mental health and psychological issues within diabetes consultations was accepted by professionals. Professionals identified gaps in their knowledge in relation to the management of mental health issues.

> H1: “It’s hard with no training [in mental health].”

> H7: “I think that we as diabetologists probably need a bit more education from them, what to look for, and you know how to get resources, et cetera. At the moment, I think I’m a bit clueless here. If even if a patient expressed a concern over their wellbeing, I would be a bit clueless as to what to ideally do, you know?”

#### Management approaches – type 1 vs type 2 diabetes

Scopes of professional practice were wide, with most looking after combinations of people with type 1 diabetes or type 2 diabetes. Management strategies for people with type 1 diabetes were often dramatically different from those with type 2 diabetes. Professionals reported an increased understanding and awareness of HbA_1c_ in those with type 1, perhaps illustrative of the younger age of diagnosis or greater emphasis on initial intensive education. People with type 1 were noted to be more driven by their HbA_1c_ targets.

> H1: “Patients with type 1, they often know their HbA_1c_, they’re driven by their targets. […] People with type 2 diabetes […] most of the time, the patient will not know their target. […] Type 1 may see the kind of risks associated with the high HbA_1c_. Type 2, it seems that confusion is still there, maybe because it’s quite new to them.”

Alongside differences in understanding of HbA_1c_, psychological issues impacting people with diabetes were noted to have differences depending on the diabetes type.

> H1: “It’s very different in type 1 and type 2 in terms of like very, very different with mental health.”

#### Patient autonomy

Current clinic processes in diabetes put emphasis on ensuring the completion of annual diabetes care processes which, whilst important, can absorb valuable clinic time and may not allow individuals to achieve their objectives for the consultation. Patient-guided consultations were seen by healthcare professionals as a method to improve patient autonomy, encourage engagement and allow patients to discuss issues they felt were important.

> H7: “I usually ask, “what do you want to achieve from this conversation?” and they bring up some issues <u>they</u> want to discuss. I will always start with whatever issues they want bring up.”

To further enhance autonomy, when considering individualised targets, the importance of allowing patient input into the decision process was highlighted by professionals, especially where the decision was considered to impact upon the relative benefits and risks of more intensive glycaemic control on patient quality of life.

> H1: “I think that it probably comes down to their quality of life, and how they feel about that treatment. […] [regarding complications, patients say] “I know the risks, but it’s my choice, it’s my life at the end of the day.””

#### Young adults

The day-to-day challenges of diabetes can be even more pronounced for young adults, who are often balancing many conflicting priorities and display symptoms of depression to a greater extent than older adults, often requiring more specifically personalised care with a greater focus on wellbeing within consultations [50,51]. Professionals were aware of the increased need for tailored care amongst their young adult patients.

> H4: “I think with the younger adult population, there’s that element of— ‘it’s not a priority to them’.”

### Diabetes and wellbeing – psychological aspects

Seven codes were used in this sub-theme to explore healthcare professionals’ perceptions of the psychological impact of diabetes on their patients: burden, complacency, coping mechanisms, family, mental health in consultations, guilt and failure, and hopes.

#### The burden of diabetes on their patients

In agreement with the experiences of patients, healthcare professionals noted having diabetes was described as a ‘weight’ individuals carry around, with professionals observing the many smaller aspects of having diabetes stacking up alongside the requirements of daily life.

> H1: “I think that the weighting of diabetes itself, like the information that they have, the treatment, the blood sugars, the kind of frequent consultations that they have, there is like a weight they’re carrying around on top of everything else that they have in their life to manage as well.”

Additionally, professionals reported feedback on sub-optimal self-management results in the form of glycated haemoglobin or day-to-day blood glucose readings can be frustrating for patients, especially where the results did not match the effort invested.

> H2: “Patients can get very overwhelmed with the diabetes. They often say they get fed up when they see high numbers […] [It’s] very frustrating for them.”

In addition to perceived detrimental effects on mental health, professionals noted the requirements of daily self-management stretched over a lifetime added to the burden of diabetes experienced by their patients.

> H8: “It’s a lifelong condition that they have to live with every day for the rest of their lives. It does hit hard on the mental health of these patients. It is <u>difficult</u>.”

#### Noting complacency in patient self-management

Professionals reported that people with diabetes often normalised chronically high blood glucose readings, echoing patient statements.

> H1: “With type 2, if they’re normally quite high [HbA_1c_] for a long period of time, they can almost get complacent with it being high. […] If something’s kind of repeated too much, they just get used to that as a normal thing they’re going to hear.”

Professionals caring for young adults with diabetes reported that many individuals often neglected diabetes self-management focusing on other life priorities.

> H5: “When you start talking about diabetes with them, for example asking about their blood glucose readings, they just say it’s a thing that they don’t even care [about] and they just came to the clinic because they received a letter to say that if you don’t attend this clinic, for the second time or third time, you’ll be discharged from the clinic.”

#### Coping mechanisms

Coping with diabetes commonly manifested as denial, with professionals reporting that their patients often did not want to know their HbA_1c_ levels, anticipating a negative result or feelings of guilt.

> H6: “I also have patients that I’ll say, “actually, you know, you’ve not had your HbA_1c_ done in a while I’ll ask if the processing nurses can do it today.” And then they’ll be like, “okay, but I’m gonna go…” They don’t want to know!”

By engaging with patients in a non-clinical context, professionals noted more active strategies of avoidance and denial. Those with a long duration of diabetes were noted to demonstrate fatigue at diabetes appointments and utilised strategies to complete clinic visits as quickly as possible.

> H1: “They said to me, […] “I know what to say to get that consultation done.””

#### Family

Support networks for patients, including family, friends, colleagues, and others with diabetes were noted by professionals to be a significant aid in supporting the wellbeing of people with diabetes.

> H2: “If I know someone’s got a mental health problem, I just dig around and I see if the patient has good support, like a supportive family, or not. [For these people], family [support] is everything.”

Reciprocally, lack of a good support network or good understanding of the requirements of diabetes self-management within support networks was reported to be detrimental to wellbeing.

> H8: “I think probably in everyday life, for a lot of them, they don’t really feel like they can just talk to family members and friends about it, because they probably don’t feel like they really understand them.”

#### Mental health in consultations

Alongside the routine structure of diabetes appointments, many professionals acknowledged they used their consultations to act upon diabetes-specific psychological or mental health issues which can create barriers to optimal management.

> H1: “It is about picking up on the right things that they say—literally key things—that one word that they say will represent a lot to them. […] I do find that mental health is threaded throughout my consultations. […] The majority of my consultations is a kind of ‘safe environment’, the tissues are being used all the time!”

Presence of underlying psychological issues was a barrier to improving diabetes management.

> H5: “I think it’s all about, “what is the underlying problem?” It depends on what the reason is for having the anxiety and mental health problems, you know, because if you don’t treat the underlying problem, you’re never able to make the diabetes better in this group.”

#### Guilt and failure

Recurrent themes of guilt and failure were noted in people with diabetes, with a perception that individuals were blamed for sub-optimal self-management depending on the approach of the professional in the consultation.

> H1: When people are sitting face to face in consultation there might be [a perception] that there’s a blame on that individual, depending on how the terminology used within that consultation as well.”

#### Hopes

Use of glycated haemoglobin as a marker of performance was used by professionals to encourage patients to focus on positive outcomes, even where targets were not achieved.

> H4: “You don’t want to leave a consultation feeling flat, I always try and give them positive focuses. So, very often, it could be that their HbA_1c_ is quite a bit better.”

When professionals discussed glycated haemoglobin in a positive context, patients were noted to demonstrate increased engagement and motivation around its use.

> H2: “The patients that I do see again, they’re always keen to know when they come back to see me: “Well, what’s my HbA_1c_?””

Alternative metrics, such as ‘time in range’, available with modern glucose monitoring technology (isCGM and real-time continuous glucose monitoring [rtCGM]) were favoured by some professionals over glycated haemoglobin readings due to their instant availability on patient devices, allowing patients to track their progress independently of the diabetes clinic and requirement for interaction with healthcare professionals.

> H6: “I know HbA_1c_ still important, but sometimes I like the fact of maybe letting the patient know that even a 5% increase in ‘time in range’ is where we’re aiming for but actually—even if you can improve it slightly—it’s gonna make quite a big difference.”

### Diabetes and wellbeing – physical aspects

Four codes were used in this sub-theme to explore healthcare professionals’ perceptions of the impact diabetes has on physical aspects of patients’ lives: blood sugars, complications, side-effects, and treatments. The impact of the coronavirus pandemic on people with diabetes arose without prompt. An additional fifth code of ‘Covid-19’ was added to this sub-theme to further review the impact of the pandemic on the wellbeing of people with diabetes.

#### Blood sugars

For many, day-to-day diabetes self-management relies on high awareness of current blood glucose values, insulin administration doses in the previous 24 hours and the anticipated insulin dose requirements for upcoming meals. Individuals with type 1 diabetes may take this one step further to improve blood glucose values by using a technique called carbohydrate counting, requiring a specific quantity of insulin administration per gram of carbohydrate consumed. Individuals using this technique become adept at judging the grams of carbohydrates on the plate, though potentially impairing the joy from eating.

> H1: “They would feel it’s like their fault. […] [they say to me] “will I ever see food as food? All I ever see is just numbers””

Whilst many people with diabetes reported improved anxiety with the recent addition of new glucose monitoring techniques such as isCGM and rtCGM, professionals noted the effect of these technologies was not always positive.

> H6: “[with isCGM and rtCGM] sometimes we see a lot of panic and constantly correcting and getting a yo-yo effect with their blood sugars.”

#### Complications in diabetes

With the knowledge and training received to obtain their roles, professionals had a heightened awareness of the complications of diabetes.

> H3: “What’s going to happen, you know, are the kidneys still going to be working? Are they still going to be able to see? […] Not knowing, are they going to end up losing limbs because of the diabetes, you know… <u>that</u> many things can go wrong.”

Many professionals discussed the risk of diabetes complications with their patients in relation to HbA_1c_ levels and targets to help them make informed decisions on self-management.

> H5: “It has to be a slow process, but I say to them, “I don’t want you to be worried just because of the HbA_1c_, but I don’t want you to get the complications of the diabetes.””

#### Side-effects of diabetes treatment

People with diabetes had high levels of anxiety about hypoglycaemia. Professionals noted the fears patients had, acknowledging the association between hypoglycaemia and stricter, more intense HbA_1c_ targets.

> H8: “Fear of hypos, I suppose because the [blood sugar and HbA_1c_] targets are so tight, they do worry about increasing their insulin.”

Professionals identified feelings of confusion among patients, with the unpredictable nature of hypoglycaemia and severe hypoglycaemia often contributing to negative feelings related to this side-effect.

> H3: “After speaking with people that have [had severe hypo], and they were like, “This has never happened before. Why is it happened now?” you know, and, “I don’t understand how it’s happened.””

#### Treatments

Noted as a burden and source of anxiety in patients, the treatment burden in diabetes was acknowledged by professionals.

> H1: “One of the most common reasons that patients want to start it [GLP-1 RAs], is because it means they won’t have to start insulin. […] A lot of the time that potential fear comes from seeing other people on insulin and how they don’t want to end up on insulin.”

Negative perceptions of insulin use remain in those with type 2 diabetes, highlighting the importance of ensuring patients are well-informed about the relative benefits and drawbacks of treatment options as part of informed decision-making.

> H2: When you tell somebody, they might need to take insulin for their diabetes, their automatic reaction is, “I’m not gonna be able to do this. It’s a big injection, it’s a needle.” […] Educating them, that kind of gets over that initial barrier.”

#### Covid-19

Professionals acknowledged the loss of face-to-face patient peer contact during the coronavirus pandemic contributed to increased patient isolation.

> H2: “So, patients speaking to other people with diabetes [can be really helpful] and is probably something that we might have lost a little bit over the past few years during Covid. […] They felt like we weren’t here [to support them]. Isolated… yeah.”

## Discussion

This study has explored the feasibility of undertaking a larger project of this nature to provide conclusive results on the effect of setting intensified or relaxed glycaemic targets on psychometric and glycaemic outcomes in people with diabetes. Alongside this, the preliminary impact of these targets on PROs and glycaemic outcomes have been reported coupled with the experiences, views, and opinions of patients and healthcare professionals on study acceptability, the impact of diabetes on wellbeing, and on glycated haemoglobin target-setting.

Study feasibility data demonstrated a sufficient population available to screen for study entry (n = 671), a sufficient number of eligible individuals (n = 109/671, 16.2%) and recruitment of the required number of participants to take part in the quantitative study (n = 50/109, recruitment rate 45.9%) within the pre-specified recruitment window. Drop-out rate was higher than anticipated at 34%, though not unexpected when compared with a meta-analysis of other interventional trials in chronic disease having an average attrition rate of 43% (95% CI 29–57) [52].

Whilst no agreed consensus on an ‘ideal’ response rate is reported in the medical literature, alignment of a mean response rate of patient surveys with the response rate of this study would seem a reasonable standard for comparison. Whilst evidence that a lower response rate can result in non-response bias is conflicting [53], determining a consensus on the mean response rate from medical literature was important to establish validity of the results reported in line with similar studies. In our study, questionnaire response rate was 70.8% when including withdrawers in the analysis. A systematic review by Meyer *et al.* [54] demonstrated a mean (±SD) response rate of 70.0% (±18.4) in 811 patient surveys reviewed between 2007 and 2020, aligning with our findings.

Bearing in mind these feasibility data, a series of recommendations and amendments should be made to the protocol of a future definitive study. Enhanced participant retention has been demonstrated in a Cochrane review by Gilles *et al.* [55] by using an open trial design, telephone reminders, monetary reimbursements and monetary incentives. Whilst this research was open in design due to the nature of the intervention, further enhancement of retention could be achieved by considering additional suggestions from the Cochrane review during future research planning stages. Adjustments to the handling of questionnaire non-responders to increase response rate should also be considered, taking into account qualitative feedback focussing on questionnaire length and ease of understanding, and considering published literature noting enhanced response rates using telephone follow-up, recorded delivery postal questionnaires with return stamped addressed envelopes, and additional rewards and incentives for questionnaire completion in non-responders.

Levels of acceptability of the study intervention were high with study participants being motivated to achieve their glycaemic goals with better self-management demonstrated by improvements seen in metrics of diabetes-related distress, self-efficacy and glycated haemoglobin levels which were apparent irrespective of whether they were randomised to receive relaxed or intensified HbA_1c_ targets.

Estimating the sample size for future studies, sample size calculations were carried out based on the mean delta values for outcome measures of interest, as displayed in Table 7. Using these figures, with an alpha value (probability of type-I error) of 0.05 and a power (probability of type-II error) of 80%, sample size estimates varied between 415 and 1,212 per group.

Evidence of the psychometric impact of HbA_1c_ targets on people with diabetes is limited. Prior studies on this topic have not directly measured PROs when reviewing the impact of HbA_1c_ targets, instead using surrogate markers [56]. Though preliminary, PRO data were of interest to evaluate data trends and generate hypotheses to test in future studies. In general, irrespective of the randomisation group, statistically significant improvements in point-of-care glycated haemoglobin levels, systolic blood pressure, diabetes-related distress, managing psychosocial aspects of diabetes and overall self-efficacy were seen 3 months post-intervention (Table 6). These findings remained apparent when considering the results stratified by intervention group, though without statistical significance. Improvements in systolic blood pressure were above the threshold of MCID [57], with the remainder of statistically significant improvements lying below the level of MCID, or having no MCID reported in the literature.

Changes to health-related quality of life were equivocal and below the threshold of MCID. The main notable between-group differences were seen in self-care, with improvements seen in those randomised to receive intensified targets when compared with deteriorations seen in those randomised to receive relaxed targets.

With improvements seen in many of the PROs measured in this study in both randomisation groups, it can be argued that the process of setting specific glycated haemoglobin targets in people with diabetes—irrespective of the target—is beneficial to PROs. Current evidence suggests improvements in PROs such as the metrics tested in this study are strong predictors of improved healthcare outcomes.

Whether the lower levels of distress seen are related to relationships with HbA_1c_ levels seen in previous studies [58], or indeed are impacted by the study intervention independent of HbA_1c_ levels is difficult to say. Conversely, prior literature has demonstrated no relationship between self-efficacy and HbA_1c_, yet both metrics improved in this study. It could therefore be argued that the improvements in self-efficacy seen are related to the study intervention. This finding would agree with previous studies demonstrating that improved patient understanding of their disease translates to improved self-efficacy and, in turn, is an important enabler of good self-management in diabetes [59–61].

When considering health state by diabetes type, those with type 1 diabetes experienced a significant deterioration in health state post-intervention according to EQ-5D-5L index scores when compared with type 2 diabetes (mean difference .114 [95% CI .006–.222, p=.038]).

This may indicate that target-setting has a significantly different psychological impact on those with type 1 diabetes compared with type 2 diabetes. Further work is needed to evaluate this phenomenon in greater detail. No other significant between-group differences were seen in PROs for diabetes type, gender or whether participants achieved the target set during the study period.

Biomedical outcomes of HbA_1c_, blood pressure and BMI are preliminary and considered hypothesis-generating. Improvements were seen in both randomisation groups with all three biomedical outcomes, with minimal differences between groups. It can be argued that the improvements seen were due to increased participant motivation from receiving specific goals to achieve, increased participant understanding of the utility of HbA_1c_ targets, increased medication adherence, the Hawthorne effect [62], a delayed effect of medications commenced prior to run-in, or by chance. Whether the lower levels of distress seen are related to relationships with HbA_1c_ levels seen in previous studies [58], or indeed are impacted by the study intervention independent of HbA_1c_ levels is difficult to say.

Regardless of the reason, the findings suggest setting a specific glycaemic goal for people with diabetes is associated with improved glycated haemoglobin levels, blood pressure and BMI in our study. The improvements seen in biomedical and psychometric outcomes are likely to be multifactorial and will require further study.

Analysis of interviews carried out with study participants and diabetes healthcare professionals were triangulated with quantitative data and added greater depth and understanding to the findings. Findings from interviews demonstrate considerable crossover between quantitative outcomes and participants’ experiences in the study, both in terms of study acceptability and PROs. Three main themes emerged from these data: study feasibility, glycated haemoglobin targets, and diabetes and wellbeing. Participants discussed factors which aided their decision to consent to participate in the study including face-to-face interaction with the researcher with the opportunity to ask questions prior to consent, and whether the research aims aligned with their ideology. Participants also discussed study retention, with reasons for continued engagement with the research being perceived health benefits, increased mood, additional healthcare interaction, and personal interest in the research topic. Participants discussed the use of glycated haemoglobin targets as part of their diabetes care, with many acknowledging limited understanding of its utility prior to study entry. With additional understanding brought motivation to achieve their target. Motivators were either circumstantial (e.g., the need to reduce HbA_1c_ before surgery) or more general (e.g., to reduce future risk of diabetes complications). Having targets that were achievable in order to maintain motivation for improved self-management emerged as a recurrent theme in interviews. Specific demotivators were numerous, highlighting the persistent effort often required to maintain good levels of self-management in diabetes. Responses coded to the final theme of ‘diabetes and wellbeing’ were further divided into psychological, physical and social (‘living with diabetes’) sub-themes. Experiences were widely varied, with some participants noting minimal impacts of diabetes on their wellbeing and others noting life-changing psychological, physical or social effects.

Interviews with diabetes healthcare professionals demonstrated significant understanding of the impact diabetes and glycated haemoglobin targets have on the wellbeing of their patients, echoing findings from patient interviews. Two main themes emerged: managing diabetes, and diabetes and wellbeing. Healthcare professionals expressed the importance of discussing HbA_1c_ targets integrated as part of a positive dialogue with patients in order to maintain engagement and motivation. Interviewees noted the importance of targeting HbA_1c_ levels to improve biomedical outcomes for their patients but acknowledged the importance of consideration of the patient perspective where functional or social goals may take precedence. The importance of building rapport with patients was highlighted, especially where psychological issues associated with diabetes were anticipated. Many noted limited training or experience in managing psychological aspects of diabetes care and the limited availability of clinical psychologists with experience in diabetes-specific psychological issues.

## Strengths and limitations

The confidence intervals for these data are wide, reflecting that the study was underpowered to demonstrate statistical significance. By nature of design, the study presented aimed to establish feasibility, and as such was not powered to output statistically significant PRO and biomedical data. Sufficient demographic information has been reported to determine the generalisability of findings, though recruiting from a single centre reduces the overall generalisability A wide range of participants were recruited in terms of age, diabetes duration, and gender. The population from which study participants were recruited has a higher diabetes prevalence, higher deprivation score, lower life expectancy and higher rates of obesity [63] than the national average in the UK. Additionally, the local population is predominantly white (98.4%) compared with 85.4% nationally [64] resulting in difficulties in recruiting people from ethnic minority groups. On this basis, the findings presented are generalisable to adults with type 1 or type 2 diabetes under secondary care diabetes services in other regions in the UK with comparable demographics. This limitation could be addressed by recruiting from additional centres in England to achieve a broadly representative sample in a future study.

Multiple linear regression was conducted to identify any potential confounding factors (age, gender, diabetes duration, IMD decile, BMI) affecting outcomes. The majority of outcomes were not influenced by potential confounders due to robust randomisation processes. BMI was noted to be a predictor of foot care processes according to the SDSCA. Reduction in foot care in those with increasing BMI may be due to functional limitations (e.g., reduced spine flexibility, limited range of movement of major joints, reduced capacity to hold prolonged fixed postures) seen in adults with obesity [65] limiting foot care processes.

Using a mixed-methods approach, the findings from the quantitative and qualitative study aspects have been considered together as a whole to increase confidence in data trustworthiness. Trustworthiness of the results was determined using Lincoln and Guba’s Evaluative criteria [42]. Credibility of the findings is enhanced using method triangulation (a combination of both qualitative and quantitative methodologies to check consistency of findings), source triangulation (studying both people with diabetes and healthcare professionals), analytical triangulation (utilising experience from the research team to confirm coding and analytical strategy), and member-checking (testing the validity of participant accounts using probing, clarifying or summarising lines of inquiry).

## Conclusion

The results of this research contribute to the limited evidence base on the nuanced topic of psychometric outcomes in diabetes and the impact of treatment targets. Based on the findings in this study, the results suggest that undertaking a larger, conclusive study is feasible with protocol amendments. Additionally, preliminary outcome results suggest setting explicit glycated haemoglobin targets in people with diabetes is associated with improvements to PROs, biomedical outcomes, and patient experience.

To implement individualised glycated haemoglobin targets in practice, a shared decision-making process between patients and clinicians taking into account risks of future complications alongside patient preferences should be utilised [66].

Evidence for the use of individualised targets in response to patient characteristics has been established before this study. Prior to this study, evidence of the impact of their use on PROs and patient experiences was limited. The results of this study demonstrate the feasibility of conducting a larger trial to provide conclusive answers on the impact explicit glycated haemoglobin targets have on PROs evaluating health-related quality of life, self-care, distress, wellbeing, and self-efficacy.

Preliminary findings indicate that glycated haemoglobin targets may benefit these PROs and indeed may improve self-management and glycaemic control through improved patient motivation and focus.

## Supporting information

S3 approved protocol

S4 patient interview topic guide

S5 Healthcare professional interview topic guide

S1 CONSORT 2010 Checklist

S2 HRA letter of approval

## Data Availability

Data are available from the Edge Hill University Institutional Data Access / Ethics Committee (contact via) for researchers who meet the criteria for access to confidential data.

## Acknowledgements

This work forms part of the first author’s PhD. S.J.W. is supported by the Department of Diabetes and Endocrinology, St Helens and Knowsley Teaching Hospitals NHS Trust, UK.

## Author contributions

S.J.W., K.H., S.W., R.P.N. and G.I. conceptualised the research aims. S.J.W prepared the manuscript for publication. S.J.W., K.H., S.W., R.P.N and G.I. reviewed and edited the manuscript. K.H., R.P.N, S.W. and G.I. mentored and supervised S.J.W. in the PhD student role. The Department of Diabetes and Endocrinology, St Helens and Knowsley Teaching Hospitals NHS Trust provides the study materials, laboratory instrumentation, computing resources and facilities for use in the research.

