## Supplementary material for "Psychometric and biomedical outcomes of glycated haemoglobin target-setting in adults with type 1 and type 2 diabetes: A mixed-methods parallel-group randomised feasibility study": S3 approved protocol

**FULL/LONG TITLE OF THE STUDY:**

A mixed-methods feasibility study evaluating the impact HbA1c target-setting has on psychometric outcomes in people with diabetes in Northwest UK

**SHORT STUDY TITLE / ACRONYM:**

HbA1c TargeT AchIevemeNt in diabeteS study (ATTAINS study)

**PROTOCOL VERSION NUMBER AND DATE**

Version: 2.0.5

Date: 26th February 2021

**RESEARCH REFERENCE NUMBERS**

**IRAS ID:** **291254**

**SPONSORS (STHK) Number:** STHK-2021-003

**Student Number:** 23723122

#### Table of Contents

**TABLE OF CONTENTS** ...... **II**

**KEY STUDY CONTACTS** ...... **IV**

**STUDY SUMMARY** ...... **IV**

**ABSTRACT** ...... **VI**

**LAY SUMMARY** ...... **VII**

**FUNDING AND SUPPORT IN KIND** ...... **VIII**

**ROLE OF STUDY SPONSOR AND FUNDER** ...... **VIII**

**ROLES AND RESPONSIBILITIES OF STUDY MANAGEMENT COMMITEES/GROUPS & INDIVIDUALS** ...... **VIII**

**PROTOCOL CONTRIBUTORS** ...... **VIII**

**KEY WORDS** ...... **IX**

**STUDY FLOW CHART** ...... **X**

11.2 APPENDIX 2 – SCHEDULE OF PROCEDURES ........ 102~~101~~
11.3 APPENDIX 3 – AMENDMENT HISTORY ........ 103~~102~~
11.4 APPENDIX 4 – STUDY ESTIMATED COSTS SUMMARY ........ 104~~103~~
11.5 APPENDIX 5 – TIMELINE ........ 105~~104~~

#### KEY STUDY CONTACTS

| | |
|---|---|
| Chief Investigator | Dr Niall Furlong*<br>Consultant physician and endocrinologist, St Helens and Knowsley Teaching Hospitals NHS Trust.<br> |
| Study Co-ordinator, protocol author | Dr Sam Westall*<br>Clinical research fellow, St Helens and Knowsley Teaching Hospitals NHS Trust.<br>PhD student researcher, Edge Hill University. Subsequently referred to in this document as "the researcher".<br> |
| Sponsor<br>(designated contact) | Mrs Jeanette Anders, Research Development and Innovation Manager, St Helens and Knowsley Teaching Hospitals NHS Trust<br> |
| Funder(s) | Department of Endocrinology and Diabetes, St Helens and Knowsley Teaching Hospitals NHS Trust |
| Key Protocol Contributors | Professor Kevin Hardy*<br><br>Dr Prakash Narayanan*<br><br>Dr Simon Watmough*<br><br>Dr Greg Irving*<br> |
| Committees | Edge Hill University Health Research Ethics Committee<br>Contact: Philip Bentley, Research Coordinator, Edge Hill University<br> |

*CV in appendix 1

#### STUDY SUMMARY

| | |
|---|---|
| Study Title | A mixed-methods feasibility study evaluating the impact HbA1c target-setting has on psychometric outcomes in people with diabetes in Northwest UK |
| Internal ref. no. (or short title) | ATTAINS Study |
| Study Design | Mixed-methods feasibility study |

| | | | | |
|---|---|---|---|---|
| Study Participants | People with diabetes and healthcare professionals involved in caring for people with diabetes | | | |
| Planned Size of Sample (if applicable) | Study A: 50 people with type 1 and type 2 diabetes<br>Study B: 15 people with type 1 and type 2 diabetes from study A<br>Study C: 15 healthcare professionals<br>Study D: 15 people with type 1 and type 2 diabetes declining entry into Study A | | | |
| Follow up duration (if applicable) | 6 months | | | |
| Planned Study Period | This is educational research to be undertaken towards a PhD. Commencement of a full-time 3-year PhD for the researcher at the Faculty of Health, Social Care and Medicine, Edge Hill University began in October 2019. | | | |
| Timetable (PhD and research) | Commencement of PhD Studies | October 2019 | | |
| | Programme of related studies | October 2019 | to | March 2020 |
| | Interruption of studies (temporary return to clinical work) | April 2020 | to | September 2020 |
| | Protocol development | September 2020 | to | November 2020 |
| | Data collection (estimated) | May 2021 | to | Feb 2022 |
| | Data analysis/write-up/publication (estimated) | July 2021 | to | May 2022 |
| | Return to specialist training programme (actual) | 5$^{th}$ September 2022 | | |
| | Thesis submission/final viva (estimated) | February – March 2023 | | |
| Research Question/Aim(s) | Aims<br>Using a mixed-methods approach, this study aims to explore:<br>1. the feasibility of evaluating the use of explicit glycaemic targets in people with diabetes.<br>2. the preliminary impact of setting explicit glycaemic targets on psychometric and glycaemic outcomes in people with diabetes.<br>3. the experiences, views and opinions of people with diabetes on the acceptability of study processes and glycaemic target-setting in the context of their diabetes care.<br>4. the experiences, views and opinions of diabetes healthcare professionals on the use of glycaemic target-setting and the individualisation of glycaemic targets in people with diabetes. | | | |

| | |
|---|---|
| | 5. the experience, views and opinions of people declining entry into study A on barriers to participation in diabetes research.<br>Research Questions<br>1. What is the feasibility of undertaking a randomised control trial evaluating the effect of setting explicit glycaemic targets on psychometric and glycaemic outcomes in people with diabetes? (**Study A**)<br>2. What is the preliminary impact of setting explicit glycaemic targets above and below current HbA1c readings on psychometric and glycaemic outcomes in people with diabetes? (**Study A**)<br>3. What are the experiences, views and opinions of people with diabetes on the acceptability of the study processes and on glycaemic target-setting? (**Study B**)<br>4. What are the experiences, views and opinions of healthcare professionals on glycaemic target-setting and on the use of individualised treatment targets in people with diabetes? (**Study C**)<br>5. What are the experiences, views and opinions of people declining entry into study A on barriers to participation in diabetes research? (**Study D**) |

#### ABSTRACT

**Background**: Individualised treatment targets are commonly used in secondary care diabetes services in the UK in order to personalise the care of people with diabetes. Still, achievement of blood glucose targets in people with diabetes is often poor. Generic and diabetes-specific mental health issues are a known driver of poor blood glucose target achievement and increased morbidity and mortality. Little is known, however, of the reciprocal effect that blood glucose target-setting has on the psychology and emotional health of people with diabetes.

**Aim**: In a series of related studies, the feasibility of evaluating psychometric outcomes when setting explicit blood glucose targets in people with diabetes will be tested.

**Methods**: Using a mixed-methods approach, a single-centre, randomised feasibility study will be conducted, aiming to recruit 50 eligible people with diabetes due to attend a secondary care diabetes clinic in the Northwest UK. Participants will be randomised into one of two groups. Group A will have explicit average blood glucose (HbA1c) targets set 5mmol/mol above their present HbA1c reading. Group B will have explicit HbA1c targets set 5mmol/mol below their present HbA1c reading. Standard clinical care will continue. Measures of feasibility (eligibility rate, recruitment rate, response rate, retention rate, acceptability of study processes for participants and investigators) and preliminary outcome (psychometric questionnaire scores and HbA1c) will be evaluated at 0, 3 and 6 months. Semi-structured interviews with a purposive sample of study participants and healthcare professionals will capture experience and perspective on the study processes, evaluate barriers to participation in research and bring further meaning to the preliminary study findings. Interviews will be analysed thematically using the framework approach.

**Original contribution to knowledge**: Whilst the current literature points to the clear physical health benefits of having individualised targets for diabetes, there is minimal understanding of the psychological effect of their use. The proposed research aims to test the feasibility of conducting a full-scale randomised control trial in evaluating psychometric outcomes when using explicit glycaemic targets in people with diabetes.

#### LAY SUMMARY

For diabetes doctors, the importance of providing personalised care and treatment goals to people with diabetes is seen to be increasingly important. A diabetes treatment goal known to be important in predicting later health problems is average blood sugar (also known as HbA1c or 'A1c'). The A1c tells us what the blood sugar of people with diabetes has been like over the past 8-12 weeks. Over time, A1c helps predict what the chances are of developing health problems later in life for someone with diabetes.

Despite an improved selection of medications and increased awareness of A1c goals amongst people with diabetes, the achievement of A1c goals has improved very little over the past decade.

Many things can get in the way of achieving A1c goals in people with diabetes. Mental health issues are known to be an obstacle to optimal blood sugar levels. Research tells us that people with diabetes who also have mental health problems struggle to reach their A1c goals. Despite the frequent use of treatment goals such as A1c in the care of people with diabetes, little is known of the reciprocal effect that goal-setting has on the psychological well-being of individuals.

The ATTAINS Study will look at the effect that A1c goal-setting has on the psychological well-being of people with diabetes. This is an early project to see if a bigger project is justified in the future. The psychological impact that A1c goal-setting has on people with diabetes is unknown. A better understanding of this could help people with diabetes achieve their A1c goals.

**FUNDING AND SUPPORT IN KIND**

| FUNDER | FINANCIAL AND NON FINANCIAL SUPPORT GIVEN |
|---|---|
| **Department of Endocrinology and Diabetes, St Helens and Knowsley Teaching Hospitals NHS Trust** | **Funded clinical research fellow post and PhD research degree fees at Edge Hill University** |

**ROLE OF STUDY SPONSOR AND FUNDER**

The sponsor is St Helens and Knowsley Teaching Hospitals NHS Trust (STHK). STHK is a research-active NHS trust in the Northwest UK. As the sponsor, STHK is responsible for the initiation and management of the research and takes primary responsibility of ensuring that the design of the study meets appropriate standards and that arrangements are in place to ensure appropriate conduct and reporting.

The funder will provide the working environment and resources for the clinical research fellow post.

**ROLES AND RESPONSIBILITIES OF STUDY MANAGEMENT COMMITEES/GROUPS & INDIVIDUALS**

Edge Hill University service user group were consulted in the design of patient-facing study documents and on the acceptability undertaking this study. Feedback from the service user group has influenced these processes.

Consideration of the James Lind Alliance research priorities for diabetes were taken into account in the development of this research.

**PROTOCOL CONTRIBUTORS**

Author of the protocol: Dr Sam Westall

Supervision/advice: Professor Kevin Hardy, Dr Simon Watmough, Dr Prakash Narayanan, Dr Greg Irving.

The PhD student will undertake this research as part of a research PhD at Edge Hill University. The study sponsor and funder will control the final decision on study design, conduct, data analysis and interpretation, manuscript writing and dissemination of results.

The James Lind Alliance (JLA) research priorities for diabetes were consulted to identify key research topics. The JLA is an organisation funded by the National Institute for Health Research (NIHR) mandated to highlight key research priorities. These are developed in a collaborative process in consultation with clinicians, patients and carers.

Edge Hill University service user group input was sought to develop the patient-facing study documents and to check the acceptability of the proposed study processes. Feedback from the service user group has resulted in adaptation of the study documents.

**KEY WORDS**

**diabetes, well-being, target, feasibility, psychometric, glycaemic**

#### STUDY FLOW CHART

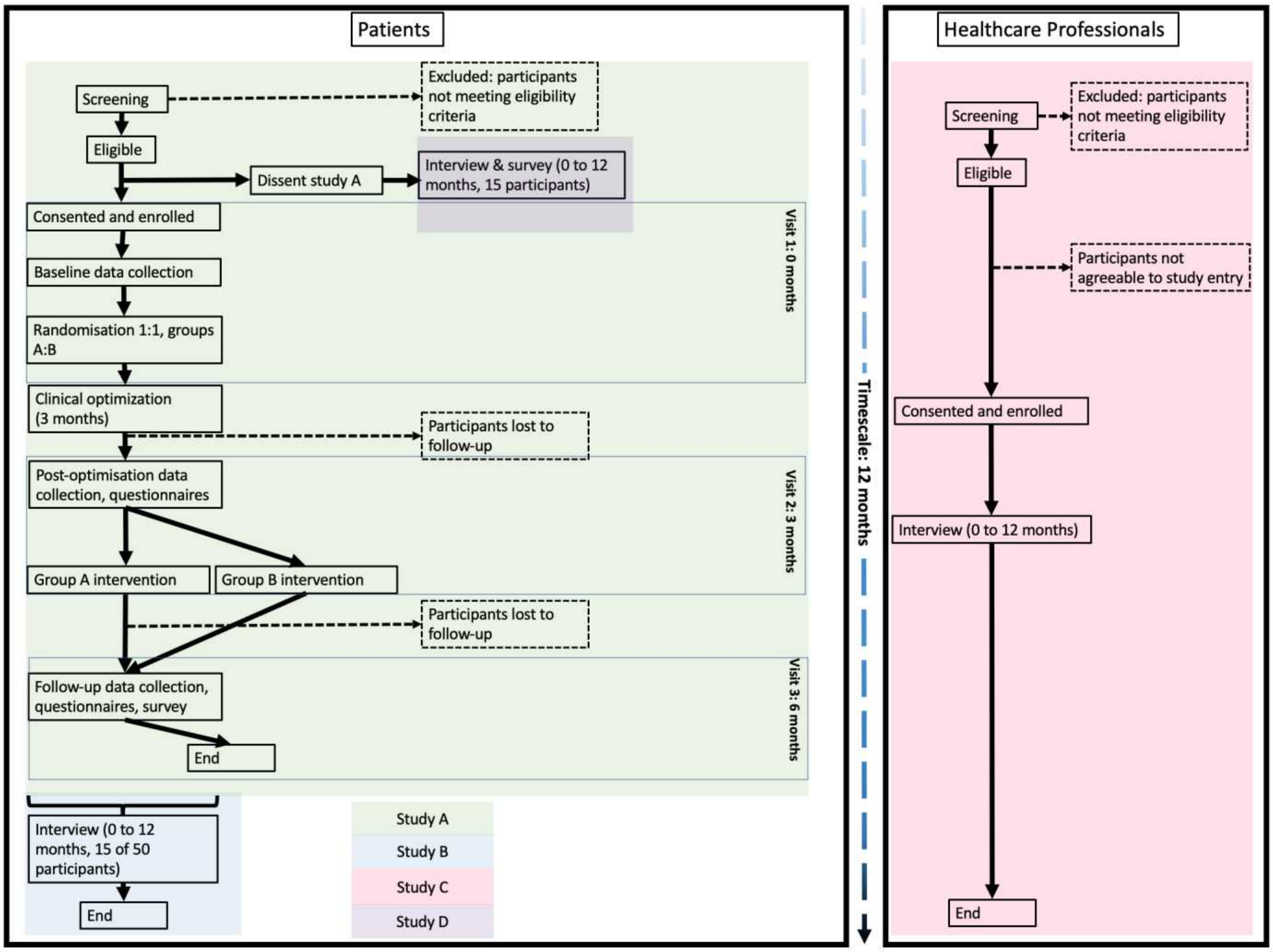

Figure 1: flowchart demonstrating the feasibility study process and timeline

**STUDY PROTOCOL: ATTAINS Study**

#### 1 BACKGROUND

For diabetes healthcare professionals, the importance of a personalised approach to management and goal-setting in people with diabetes has never been clearer, evidenced by the release of the joint ADA/EASD consensus statement (Davies et al., 2018). A diabetes treatment goal known to be important in predicting later health problems is average blood glucose (HbA1c). HbA1c tells us what the blood glucose of people with diabetes has been like over the past 8-12 weeks (Welsh, Kirkman and Sacks, 2016). Over time, HbA1c helps predict what the chances are of developing health problems later in life for someone with diabetes (World Health Organization, 1999). Chronically high HbA1c readings have been shown to negatively impact upon the physical health of people with diabetes: diabetes is one of the leading causes of adult blindness in the UK (Liew, Michaelides and Bunce, 2014), increases the risk of cancer (Noto et al., 2011, Ohkuma, Peters and Woodward, 2018) and dementia (Cheng et al., 2012, Chatterjee et al., 2016), more than doubles the risk of heart attack (NHS Digital, 2016), trebles the risk of stroke (NHS Digital, 2016) and is the commonest cause of end-stage renal failure (NHS Digital, 2016) and non-traumatic lower limb amputation in Europe (NHS Digital, 2016).

Several factors have a bearing on the achievement of glycaemic goals in people with diabetes (Ali et al., 2013). From a patient's perspective, lower levels of motivation and knowledge and higher levels of mental health comorbidity can create barriers to achieving optimal HbA1c levels (Khunti et al., 2018). From a healthcare perspective, contradictory guidelines and complex management options can cause confusion for clinicians. Underfunded NHS healthcare systems struggling to achieve national audit standards are often overwhelmed by the quantity and cost of diabetes-related healthcare interventions (Ali et al., 2013).

Although awareness of the importance of achieving glycaemic targets amongst patients with diabetes is improving with patient education (Berikai et al., 2007), many patients remain unaware of the preventable excess risk posed by inadequate HbA1c control. Despite many new pharmacological interventions that have become available in the past decade, achievement of HbA1c treatment targets in England and Wales has remained stubbornly below 30% for T1DM and below 70% for T2DM (NHS Digital, 2018). Evidence for lack of progress in the achievement of optimal glycaemic control is reflected in studies globally (Khunti et al., 2018). Outdated guidelines and national audit standards in England have limited progress towards the personalisation of diabetes care. People with diabetes have historically struggled to access evidenced-based diabetes education (though this is improving) and negative patient perceptions on the intensification of diabetes treatment remain prevalent (O' Connor, Crabtree and Yanoshik, 1997, Grant et al., 2011, Khunti et al., 2018).

Awareness amongst healthcare professionals of the importance using treatment targets to drive improvement in micro- and macro-vascular outcomes is good. Compelling evidence on the use of personalised glycaemic treatment targets in response to patient factors is gaining traction amongst clinicians in primary and secondary care. Little evidence exists, however, on the long term impact that clinician-initiated glycaemic target-setting has on psychological and self-management abilities of patients. Further work is required.

The ATTAINS Study will look at the effect that A1c goal-setting has on the psychological well-being of people with diabetes. This is feasibility study to see if a bigger project is justified in the future. The psychological impact that HbA1c goal-setting has on people with diabetes is unknown. A better understanding of this could help people with diabetes achieve their A1c goals.

#### 2 RATIONALE

The current literature points to the clear reduction in the risk of the complications of diabetes by having individualised treatment targets (Diabetes Control and Complications Trial Research Group et al., 1993, Turner, 1998, UK Prospective Diabetes Study (UKPDS) Group, 1998, The ACCORD Study Group, 2008, Ismail-Beigi et al., 2011). There is minimal understanding of the psychological effect of treatment targets in people with diabetes.

Depending on individual patient factors, current local practice at STHK results in setting glycaemic targets that are either higher (more relaxed) or lower (more stringent) than patient's current HbA1c readings. It is felt that studying the psychological impact of setting HbA1c targets in people with diabetes is essential. Risk assessments of higher and lower HbA1c target use are covered in section 8.1, 'Assessment and management of risk'.

The rationale for studying the use of explicit targets both above and below current HbA1c readings stems from current practice, testing whether "over-achievement" (i.e. the participants HbA1c being better than their target) or "under-achievement" have differing psychological implications.

In the planning of a large study, such as a randomised control trial, investigators draw upon many resources. This often includes past experience, findings from other studies and the views of patients and colleagues. Where uncertainties lie on the viability of a large study, preparatory studies are often needed (NIHR, 2016). The proposed feasibility study aims to see if it is feasible to undertake a larger trial in evaluating the psychological impact of HbA1c target-setting in people with diabetes.

#### 3 THEORETICAL FRAMEWORK

This study will address the research questions using a mixed-methods approach, relying on both qualitative and quantitative techniques. As described succinctly by Johnson and Onwuegbuzie (2007), mixed methods research is "the class of research where the researcher mixes or combines quantitative and qualitative research techniques, methods, approaches, concepts or language into a single study or set of related studies." Johnson et al. (2007) summarise mixed methods research as offering a "third methodological paradigm" alongside traditional qualitative and quantitative methods. This can help bring a richness to quantitative data and help to usefully triangulate qualitative data to produce research that becomes more than the sum of its parts (Johnson, Onwuegbuzie and Turner, 2007, Fielding, 2012). From the perspective of this research, employing a mixed method approach will bring greater real-world relevance to the data and will consequently produce findings that are meaningful for both patients and healthcare professionals. Applying methodological approaches using both qualitative and quantitative methodologies will bring enhancements to the overall project in excess of the sum of the original datasets, each addressing the limitations of the other.

Answering the research questions using quantitative feasibility outcomes defined by the NIHR (2016) and using semi-structured interviews in both people with diabetes and healthcare professionals, the findings can be triangulated to bring richness to the data and to help determine if this preliminary study justifies a larger study of this nature in the future. Semi-structured interviews will be carried out from a phenomenological (i.e. study of the participants' lived experiences of the events in the study) perspective (Creswell and Poth, 2016) and analysed thematically using the framework method of data analysis (Gale et al., 2013).

From a quantitative perspective, alongside the primary outcomes of feasibility (rates of eligibility, recruitment, retention, response), preliminary data on the impact of A1c goal-setting has on the psychological well-being of people with diabetes will be gathered.

#### 4 RESEARCH QUESTION/AIM(S)

1. What is the feasibility of undertaking a randomised control trial evaluating the effect of setting explicit glycaemic targets on psychometric and glycaemic outcomes in people with diabetes? (**Study A**)
2. What is the preliminary impact of setting explicit glycaemic targets above and below current HbA1c readings on psychometric and glycaemic outcomes in people with diabetes? (**Study A**)
3. What are the experiences, views and opinions of people with diabetes on the acceptability of the study processes and on glycaemic target-setting? (**Study B**)

4. What are the experiences, views and opinions of healthcare professionals on glycaemic target-setting and on the use of individualised treatment targets in people with diabetes? (**Study C**)

5. What are the experiences, views and opinions of people declining entry into study A on barriers to participation in diabetes research? (**Study D**)

##### 4.1 Objectives

Using a mixed-methods approach, these related studies aim to explore:

1. the feasibility of evaluating the use of explicit glycaemic targets in people with diabetes.
2. the preliminary impact of setting explicit glycaemic targets on psychometric and glycaemic outcomes in people with diabetes.
3. the experiences, views and opinions of people with diabetes on the acceptability of study processes and glycaemic target-setting in the context of their diabetes care.
4. the experiences, views and opinions of diabetes healthcare professionals on the use of glycaemic target-setting and the individualisation of glycaemic targets in people with diabetes.
5. the experience, views and opinions of people declining entry into study A on barriers to participation in diabetes research.

##### 4.2 Outcomes

From a quantitative perspective, primary outcome data will be captured on:

- the percentage of patients due to attend the secondary care clinic meeting eligibility criteria.
- the recruitment rate (including assessment of barriers to participation).
- the retention rate.
- questionnaire response rate (including rate of question non-response, rate of questionnaire completion).
- the acceptability of the study processes (randomisation, intervention, procedures, questionnaires, time commitments).

Other (preliminary) outcome data will be captured on:

- Change in HbA1c (0, 3 and 6 months – pre-clinic, pre-intervention and post-intervention).
- Change in psychometric questionnaire scores (3 and 6 months – pre-intervention and post-intervention).

Qualitative semi-structured interview data will be capture patient and healthcare professional perspectives on the topics of study process acceptability, A1c target use and the personalisation of A1c targets. In those declining entry into study A, quantitative semi-structure interview data will be used to capture patient data on barriers to diabetes research participation.

#### 5 STUDY DESIGN and METHODS of DATA COLLECTION AND DATA ANALYSIS

This is a mixed-methods study focussing on determining the feasibility of evaluating psychometric outcomes when setting explicit glycaemic targets in people with diabetes.

Quantitative data will be captured on feasibility outcomes and preliminary psychometric outcomes and the acceptability of the study intervention (**Study A**). Recruited participants will be surveyed on their perceived barriers to ongoing study participation.

Qualitative semi-structured interviews will obtain the experiences, views and opinions of people with diabetes (**Study B**) and healthcare professionals (**Study C**) on the study processes and the use of and impact of glycaemic target setting on the psychological well-being of people with diabetes.

Patients eligible for study A but declining participation will be asked if they would be willing to take part in a follow-up semi-structured interview study evaluating barriers to participation (**Study D**).

##### 5.1 Study A

Design: Single-centre, parallel group, randomised feasibility study. This study will test the feasibility of conducting a randomised controlled trial (RCT) evaluating the psychometric impact of setting explicit glycaemic targets in people with diabetes (type 1 and type 2). There is deliberate separation of the initial clinic appointment (visit one; 0 months) and delivery of the intervention (visit two; 3 months) to allow for clinical care to be optimised and avoid conflict between the clinician advice and the study target. A short follow-up period 3 months post intervention has been selected to allow testing of feasibility outcomes and preliminary psychometric and glycaemic outcomes whilst minimising the health impact of the HbA1c target change to the participants.

Study Processes: people with diabetes meeting inclusion criteria and not excluded by exclusion criteria who are due to attend new patient diabetes clinic will be screened for entry into study A. Patients meeting eligibility criteria will be invited to entry into the study by postal letter with an included participant information sheet. Follow-up telephone calls will check patient interest and coordinate screening visits.

*At visit one (0 months),* invited patients will attend for a screening visit and informed consent process at the time of attendance to their diabetes clinic appointment. Those declining to take part will be asked to complete a short survey evaluating barriers to participation (Barriers to Participation survey, Section 11.1, Appendix 1). Baseline characteristics will be recorded: NHS number, age, sex, ethnicity, diabetes type, diabetes duration, postcode, point of care (POC) HbA1c, Blood Pressure (BP), BMI, educational status and prescribed glucose-lowering drugs. Participants will attend their usual diabetes clinic appointment for optimisation of their diabetes care. Participants will be randomised (stratified [strata: type 1 diabetes, type 2 diabetes]] random permuted block strategy) 1:1 into two groups using an online randomisation service (Altman and Bland, 1999, Suresh, 2011, Sealed Envelope Ltd, 2020).

*At visit two (3 months)*, participants will visit to have a POC HbA1c reading and complete a series of validated baseline psychometric questionnaires (EQ-5D-5L – Health-related quality of life measure, SDSCA – Summary of Diabetes Self-Care Activities, PAID – Problem Areas in Diabetes, DES-LF – Diabetes Empowerment Scale, WBQ-12 – Well-being Questionnaire). The study intervention will be delivered as described using the TIDieR Checklist (Hoffmann et al., 2014):

- Intervention: Participants in group A are given an explicit HbA1c target 5mmol/mol above their current HbA1c reading. Participants in group B are given an explicit HbA1c target 5mmol/mol below their current HbA1c reading.
- Rationale: The feasibility of studying the use of glycaemic target-setting on psychometric outcomes in people with diabetes is unknown. Over-achievement and under-achievement of glycaemic targets has an unknown effect on psychometric outcomes and long term glycaemic control in people with diabetes.
- Materials: Participants will be provided with a sterilised, laminated leaflet (see Appendix 2) explaining in lay terms the rationale for HbA1c monitoring in diabetes. The leaflet will include a diagrammatic representation of their current HbA1c reading against their explicit target. A short scripted HbA1c summary will be read to participants by the researcher, along with an opportunity for participants to ask questions.
- Intervention provider: the intervention will be provided by the researcher employed by St Helens and Knowsley Teaching Hospitals NHS Trust. The researcher has a background working as a specialist trainee in diabetes and endocrinology and is fully trained in infection control measures, including precautions related to COVID-19.
- Mode of intervention delivery: the intervention will be provided face-to-face on an individual basis, following the latest UK government guidance on personal protective equipment and social distancing.
- Location: the intervention will be in a consultation room in the Diabetes Centre in St Helens Hospital.
- Time requirements/timing: the intervention will take place 3 months after study entry to allow for initial secondary care clinical management (undertaken at study entry) to be fully

established. Patients will attend the Diabetes Centre to complete a series of baseline questionnaires (30 minutes), a blood sample (10 minutes) and the intervention (5 minutes).

- Both the participants and researcher will be unmasked to intervention allocation. Clinical management of the patient will not be altered.

*At visit 3 (6 months)*, participants will have a POC HbA1c reading and repeat the psychometric questionnaires. Participants will complete a study acceptability survey. Participants will attend their follow-up diabetes clinic appointment as normal to ensure optimal management is maintained. Glycaemic targets will be reset to the evidence-based appropriate target by the clinician. End of study A.

Data collection: A password-protected Microsoft Excel dataset template will be created for data collection purposes. The dataset master copy will be saved on an NHS-encrypted computer. Daily cloud backups are made to minimise data loss (see research data management plan). Data will be obtained from the following sources:

1. Eligibility/recruitment/retention/response rate data will be counted per participant and recorded.
2. Blood test POC HbA1c data will be captured from electronic patient records systems.
3. Baseline demographic data will be obtained firstly from electronic patient records systems and if unavailable, directly from the patient.
4. Physiological measurement (blood pressure, height, weight) will be obtained at participant visits.
5. Paper questionnaires will be completed by hand by study participants and subsequently inputted into the dataset by the study investigator.

Quality control of all equipment will be as per NHS standards.

Data storage: Key research data files containing any sensitive information (e.g. data collected) will be stored on an NHS-issued, encrypted, password-protected NHS laptop. Data will be stored via the laptop on NHS trust cloud storage associated with the PhD student's clinical research fellow post in the NHS. Therefore, if data were to be lost or become corrupt, for whatever reason, regular (daily) backups are accessible from the NHS trust network. The PhD student, Dr Sam Westall, will be responsible for maintaining data storage, backups and security. Physical data (e.g. completed questionnaires, audio recording devices, consent forms) will be catalogued, filed securely and placed in a separate, locked room on NHS property within one day of receipt. Physical data will be accessed when required on the hospital site. Physical data will remain on-site until no longer required, at which point it will be destroyed. Patient identifiers will be held in a separate, password-protected database and linked through a unique study code. Separation of patient identifiers from other data will be at the point of entry into the database.

Data analysis: Participant characteristics and feasibility outcomes will be reported with descriptive statistics, presenting continuous variables as means (standard deviation, SD) and categorical variables as numbers (%). For psychometric outcomes, paired baseline and endpoint data will be presented as mean differences (95% confidence interval, p-value). The paired two-tailed student's t-test will be used to compare means of parametric datasets (EQ-VAS index scores, PAID score, SDSCA scores, DES-LF scores, WBQ-12 scores). Differences between categorical variables will be analysed using the chi-squared test (EQ-5D-5L dimension responses). The Wilcoxon rank-sum test will be used to compare non-parametric data sets (EQ-5D-5L index scores). Correlations between different psychometric outcomes will be determined using spearman's rank correlation coefficient. Statistical methods will use SPSS v.25 statistics software (IBM Corp., 2017) and expert statistician input.

##### 5.2 Study B

Design: At study A visit 1, participants will be invited to take part in a telephone semi-structured interview. Participant information sheets will be supplied. A purposive sample of eligible participants will be screened and for entry into study B. On entry into the study, participants will be assigned a unique study ID. A convenient telephone appointment for the interview will be offered. To fully explore a range of views, experiences and opinions, semi-structured interviews will be carried out until adequate information power is achieved.

Data collection: Telephone semi-structured interviews will be audio-recorded. Interview structure will be guided by the study B interview guide (see appendix 1). Audio data will be anonymised for place- and person-identifiable information. Recordings will be transcribed verbatim into a password-protected Microsoft Word document on an NHS-encrypted computer for analysis. Direct quotations will be anonymised. Any identifiable free-text data from participants' questionnaires or transcriptions will have their meaning generalised to prevent disclosure.

Data storage: Audio data will be stored on a password-protected electronic dictaphone which will be kept on NHS premises in a secure research room in the diabetes centre.

Data analysis: Transcripts will be coded to highlight key concepts and analysed using the framework method for the content analysis of qualitative data (Gale et al., 2013) and using the qualitative data analysis support software, NVivo (QSR International Pty Ltd., 2018).

##### 5.3 Study C

Design: Healthcare professionals working in secondary care diabetes services in STHK who are directly involved in the care of people with diabetes will be invited via email to participate in the research. Participant information will be attached to the emails. Replies to the email invitation will be followed up with agreement of a suitable time and place for the interview. Non-repliers will receive a follow-up email one week after the initial invite. On agreement of entry into the study, written, informed consent will be taken and participants will be assigned a unique study ID. Completed electronic copies of consent forms will be accepted if they are sent from a corresponding staff email address. A telephone or videocall interview appointment will be offered. To fully explore a range of views, experiences and opinions, semi-structured interviews will be carried out until adequate information power is achieved.

Data collection: Telephone semi-structured interviews will be audio-recorded. Interview structure will be guided by the study C interview guide (see appendix 1). Audio data will be anonymised for place- and person-identifiable information. Recordings will be transcribed verbatim into a password-protected Microsoft Word document on an NHS-encrypted computer for analysis. Direct quotations will be anonymised. Any identifiable free-text data from participants' questionnaires or transcriptions will have their meaning generalised to prevent disclosure.

Data storage: Audio data will be stored on a password-protected electronic Dictaphone which will be kept on NHS premises in a secure research room in the diabetes centre.

Data analysis: Transcripts will be coded to highlight key concepts and analysed using the framework method for the content analysis of qualitative data (Gale et al., 2013) and using the qualitative data analysis support software, NVivo (QSR International Pty Ltd., 2018).

##### 5.4 Study D

Design: Study D will recruit eligible patients declining entry into study A. Patients will be invited by telephone to take part in a semi-structured interview. A participant information sheet and consent form with return stamped, addressed envelope will be sent to those interested in taking part. On receipt of consent, participants will be assigned a unique study ID. A convenient telephone appointment for the interview will be arranged. To fully explore a range of views, experiences and opinions, semi-structured interviews will be carried out until adequate information power is achieved.

Data collection: Telephone semi-structured interviews will be audio-recorded. Interview structure will be guided by study D interview guide (see appendix 1). Audio data will be anonymised for place- and person-identifiable information. Recordings will be transcribed verbatim into a password-protected Microsoft Word document on an NHS-encrypted computer for analysis. Direct quotations will be anonymised. Any identifiable free-text data from participants' transcriptions will have their meaning generalised to prevent disclosure.

Data storage: Audio data will be stored on a password-protected electronic Dictaphone which will be kept on NHS premises in a secure research room in the diabetes centre.

Data analysis: Transcripts will be coded to highlight key concepts and analysed using the framework method for the content analysis of qualitative data (Gale et al., 2013) and using the qualitative data analysis support software, NVivo (QSR International Pty Ltd., 2018).

##### 5.5 All studies

Data preservation: All data collected could be of potential value for future research. As such, the collected, anonymised computer data will be submitted to the Edge Hill University research data repository for use.

Physical copies of data will be kept for the duration specified by St Helens and Knowsley Teaching Hospitals NHS Trust (STHK) Research Guidelines, at which point further retention of the data can be reviewed. Consent forms will be kept for as long as the research data are retained in the data repository. Following the completion of the research, they will be digitised and stored securely (encrypted) with the university. At this point, original physical copies will be destroyed securely (https://www.ukdataservice.ac.uk/manage-data/legal-ethical/consent-data-sharing/consent-forms).

COVID-19 considerations: At the time of writing (December 2020), there is a great deal of concern and uncertainty within the healthcare and research communities related to the global COVID-19 outbreak. It is noted that particular care must be taken in vulnerable patient groups such as in the elderly, in those from black, Asian and ethnic minority groups and in those with diabetes. Concomitantly working in a healthcare environment, the researcher is up-to-date with the current *social distancing* and *personal protective equipment (PPE)* practices which will guide interactions with patients.

During lockdown periods, due to the complex nature of diabetes, people with diabetes will still require face-to-face clinic time. Should any further lockdown be announced by the UK government, it is still anticipated that patients will attend the diabetes centre for their appointments. Recruitment of participants into this study is therefore not anticipated to be significantly impacted upon by future COVID-related lockdowns.

Advice and guidance on the response to COVID-19 is changing rapidly and so the utmost care will be taken in complying with the current Public Health England/National Institute of Health Protection guidance on reducing the spread of COVID-19.

### 6 STUDY SETTING

DATA COLLECTION SITE (single centre):

Diabetes centre, St Helens Hospital, Marshalls Cross Road, St Helens, WA9 3DA, UK

SITE ACTIVITIES:

Participant identification, recruitment, data collection, data analysis, data storage.

### 7 SAMPLING AND RECRUITMENT

#### 7.1 Eligibility Criteria

##### 7.1.1 Inclusion criteria

Study A: All patients with poor glycaemic control (HbA1c 64 to 125 mmol/mol) aged ≥ 18 due to attend new patient clinic who are able to provide informed consent and have a confirmed diagnosis of diabetes mellitus (including Type 1 and Type 2) will be screened for entry into study A.

Study B: Patients enrolled in study A will be considered for inclusion in study B.

Study C: Healthcare professionals (including consultant, specialist nurse, nurse consultant, dietician, specialist registrar and specialty doctor grades) working at St Helens and Knowsley Teaching Hospital NHS Trust who are directly involved in the care of people with diabetes will be considered for inclusion in study C.

Study D: Eligible patients declining entry into study A will be approached for inclusion in study D.

##### 7.1.2 Exclusion criteria

Study A:

1. Patients at risk of CVD events.
2. Patients with an episode of severe hypoglycaemia with the past 12 months.
3. Patients with hypoglycaemia unawareness (defined as a GOLD score ≥4).
4. Patients unwilling to self-monitor blood glucose at home (if clinical management requires).
5. Patients unwilling to inject insulin (if clinical management requires).
6. BMI ≥ 45 kg/m$^2$.
7. Patients who have opted-out from being contacted by researchers under the NHS national data opt-out service. (NHS Digital, 2020)
8. Patients with other serious illnesses which may limit survival or factors which may limit adherence to study interventions, such as:

- Malignancy within the past 2 years
- Dementia
- Alcohol abuse
- Plans to move out of the area
- Unreliability
- Frailty (Clinical Frailty Scale ≥ 5)

9. Patients currently participating in another clinical trial.
10. Patients with a transplanted organ.
11. Pregnancy.
12. Patients with requirements for regular blood transfusion or venesection.
13. Ongoing medical therapy known to cause difficulties with glycaemic control (e.g. corticosteroid therapy).

Study B: patients unable to communicate via telephone for the purposes of an interview (e.g. due to deafness).

Study C: clinicians involved in this research will be excluded from the study.

Study D: patients unable to communicate via telephone for the purposes of an interview (e.g. due to deafness).

#### 7.2 Sampling

##### 7.2.1 Size of sample

Study A: Influential papers (Julious, 2005, Sim and Lewis, 2012) on the sample size requirements for feasibility studies have suggested between 24 and 50 participants are required to estimate the standard deviation of an outcome measure (often needed in the calculation of sample sizes in larger studies).

Given an estimated study participation rate of 70% of those eligible, based on participation rates of eligible patients in the DAFNE (DAFNE Study Group, 2002) and DESMOND (Davies et al., 2008) trials, 72 eligible patients will be selected for entry into the study in order to enrol 50 participants (70% x 72 ≈ 50).

Depending on eligibility and recruitment rates, the sample size may have to be adapted.

Study B: A stratified (by type 1 and type 2 diabetes), purposive estimated sample of 15 participants will be required. Sample adequacy will be determined using the concept of information power, described by

Malterud et al. (2016). Estimates of sample size required in similar studies suggest a requirement of 10 – 15 participants. Adjustments to sample size may be required during the study, depending on the information power of the sample.

Study C: A convenience sample of 15 participants will be recruited. Determination of sample adequacy as per study B. Adjustments to sample size may be required during the study, depending on the information power of the sample.

Study D: A stratified (by type 1 and type 2 diabetes), purposive estimated sample of 15 participants will be required. Sample adequacy will be determined as per study B. Estimates of sample size required in similar studies (Taylor et al., 2007, Harrop et al., 2016, Hughes-Morley et al., 2016) suggest a requirement of 10 – 39 participants. Adjustments to sample size may be required during the study, depending on the information power of the sample.

###### 7.2.2 Sampling technique

Study A: Patients due to attend new patient diabetes clinic at St Helens Hospital Diabetes centre will be screened for entry into study A. Patients meeting eligibility criteria will be invited to enter the study by postal letter with an included participant information sheet (see appendix 1). Invites will be followed up with telephone calls. Patients who express interest in taking part in the study will be approached for recruitment into the study on attendance to their diabetes clinic appointment. Sampling will continue until the required number is achieved.

Study B: fifteen participants who took part in study A will be recruited in a stratified (for type 1 and type 2 diabetes), purposive manner.

Study C: a convenience sample of 15 healthcare professionals directly involved in secondary care diabetes services (including consultant, specialist nurse, nurse consultant, dietician, specialist registrar and specialty doctor grades) will be selected for interview.

Study D: fifteen patients who were eligible for and declined participation in study A will be recruited in a stratified (for type 1 and type 2 diabetes), purposive manner.

##### 7.3 Recruitment

###### 7.3.1 Sample identification

Study A: upcoming new-patient diabetes clinic lists are available on the hospital patient administration system. Hospital electronic patient records will be reviewed to identify potentially eligible participants on this list. Potentially eligible participants will be invited to a screening visit occurring at the time of their clinic appointment. Following screening, eligible patients who provide written, informed consent to take part in the study will be enrolled.

The researcher is a medical doctor in the diabetes centre at St Helens Hospital and therefore already a member of the patient's existing clinical care team. The list of patients due to attend a face-to-face diabetes clinic appointment can be obtained by the researcher in his NHS role. Clinical care of patients involved in the study will be undertaken by a clinician external to the research.

Study B: A purposive sample of participants enrolled in study A will be provided with participant information sheets on study B and selected for telephone interview. A mutually agreed date and time will be agreed with the participants on completion of the final visit of study A.

Study C: the hospital internal email system will be used for recruiting diabetes healthcare professionals for the interviews.

Study D: eligible patients declining entry into study A will be invited to study D via letter with enclosed participant information sheet. A purposive sample will be selected for telephone interview. A mutually agreed date and time will be agreed with participants.

##### 7.3.2 Consent

Consent forms (see appendix 1):

Study A: written, informed consent will be obtained using an adapted template from the HRA website.

Study B: written, informed consent will be obtained using an adapted template from the HRA website.

Study C: completion of electronic consent forms (.pdf) from healthcare professionals' email addresses will be obtained using an adapted template from the HRA website.

Study D: written, informed consent will be obtained using an adapted template from the HRA website.

Service user group feedback noted that the consent form documents were easy to understand, taking into account reading-ease, length, clarity and content.

Consent process:

Study A: participant information sheets will be sent in advance. On arrival to the clinic, the researcher will greet the patient and ensure that they have read and fully understand the information. Any questions will be answered by the researcher. In those agreeing to take part in the research, written, informed consent will be obtained and countersigned by the researcher. Three separate copies of the consent form will be signed: one for the participant, one for the research records and one for the patient's healthcare records.

Study B: participant information sheets will be provided during study A. At the final study A visit, the researcher will ensure that those taking part in study B have read and understood the participant information sheet. Any questions will be answered by the researcher. In those agreeing to take part, written, informed consent will be obtained and countersigned by the researcher. Three separate copies of the consent form will be signed: one for the participant, one for the research records and one for the patient's healthcare records.

Study C: participant information sheets will be sent out via email. Any questions will be answered by the researcher via email. Electronically-completed consent forms will be accepted from healthcare professionals from their professional email addresses.

Study D: participant information sheets will be sent out in advance. Questions from participants will be answered in a follow-up telephone call. In those agreeing to take part, written, informed consent will be obtained. Forms will be returned via enclosed stamped, addressed envelopes.

Participant information sheets: service user group feedback mentioned that the participant information sheets were clear and easy to understand. The service user group suggested greater clarity on the consent process (that it should mention that informed consent will be obtained from each participant and that it can be withdrawn at any time). Participant information sheets were amended following service user group feedback.

### 8 ETHICAL AND REGULATORY CONSIDERATIONS

#### 8.1 Assessment and management of risk

A thorough risk assessment has been carried out. Risks are scored according to severity (1 - low, 2 - medium, 3 - high) and likelihood (1 - unlikely, 2 - potentially likely, 3 - likely). This gives a risk score. 1-3: low risk, 4-6 medium risk, 7-9 high risk.

RISKS TO PARTICIPANTS.

1. Adverse event (AE) or serious adverse event (SAE) occurs.

Pre-mitigation risk:

Severity: 2 (AE), 3 (SAE). Likelihood: 1. Risk: 2 - 3 (Low)

Mitigation:

The researcher is trained in good clinical practice to ensure that AE’s and SAE’s are dealt with appropriately. The researcher is trained in conflict resolution, safeguarding adults and children, health and safety, fire safety and advanced life support.

Post-mitigation risk:

Severity: 2 (AE), 3 (SAE). Likelihood: 1. Risk: 2 - 3 (Low)

2. Confidential data breach.

Pre-mitigation risk:

Severity: 3. Likelihood: 1. Risk: 3 (Low)

Mitigation:

The researcher is trained in good clinical practice to minimise the risk of confidential data breaches. The researcher is trained and up-to-date with NHS information governance training. Research data management policy has been created to plan the safe and secure storage of data, with appropriate data anonymisation practices in place.

Risk rationale:

The use of confidential patient information is required to contact patients and analyse their blood results.

Post-mitigation risk:

Severity: 3. Likelihood: 1. Risk: 3 (Low)

3. Impact on emotional wellbeing.

Pre-mitigation risk:

Severity: 3. Likelihood: 1. Risk: 3 (Low)

Mitigation:

Patients will be consented for the reporting of health-related findings. Alongside the discovery of health-related findings, should the studies find that a participant's mental health has been affected by study processes, or that the participant had a pre-existing undiagnosed mental health problem, the participant will be sign-posted to appropriate support options and will be asked if we can let their GP know. This may help participants who were unaware that they could benefit from support in improving their psychological wellbeing.

Risk rationale:

The study is designed to evaluate the feasibility of testing the psychometric outcomes of goal-setting in participants. An integral part of testing psychometric outcomes is an evaluation of a participant's wellbeing, health-related quality of life and distress.

Post-mitigation risk:

Severity: 3. Likelihood: 1. Risk: 3 (Low)

4. Discovery of health-related findings.

Pre-mitigation risk:

Severity: 3. Likelihood: 1. Risk: 3 (Low)

Mitigation:

Participants will be consented for health-related findings on entry into the study. Should a health-related finding be uncovered by the researcher, they will (with the participant’s consent) inform the participant's GP so that appropriate evaluation of the finding can be initiated. By taking part in the research, the participants benefit from discoveries related to their health due to increased exposure to clinicians. This may lead to earlier diagnoses than would otherwise have occurred.

Post-mitigation risk:

Severity: 3. Likelihood: 1. Risk: 3 (Low)

5. Risk of capillary blood sampling to the patient (pain, bruising, bleeding).

Pre-mitigation risk:

Severity: 1. Likelihood: 3. Risk: 3 (Low)

Mitigation:

Capillary blood sampling via skin puncture with a lancet (“finger-pricking”) is generally considered as a low-risk procedure performed routinely in clinical practice by medical and nursing staff and is commonly performed by patients who self-monitor their blood glucose. Generally accepted complications of finger-pricking are: mild and temporary pain of skin puncture; minor bruising to the site following the procedure, and minor bleeding following the procedure.

Risk rationale:

The use of blood samples is required as part of the study intervention.

Post-mitigation risk:

Severity: 1. Likelihood: 3. Risk: 3 (Low)

6. Blood sample analysis error (e.g. sample insufficient).

Pre-mitigation risk:

Severity: 1. Likelihood: 1. Risk: 1 (Low).

Mitigation:

It is accepted that occasionally, an error can occur with a blood sample that means that the sample cannot be analysed. This could be due to many different reasons but invariably means that, on an infrequent basis, repeat blood samples are required.

Post-mitigation risk:

Severity: 3. Likelihood: 1. Risk: 3 (Low)

7. Time commitments.

Pre-mitigation risk:

Severity: 1. Likelihood: 3. Risk: 3 (Low).

Mitigation:

Study processes occur when patients are due to come to the diabetes clinic. Psychometric questionnaires have been selected based on their content validity, reliability, acceptability, precision and responsiveness. Time commitments of completing questionnaires were considered during the questionnaire selection process.

Post-mitigation risk:

Severity: 1. Likelihood: 3. Risk: 3 (Low).

8. Additional COVID-related risks to patient research participants (risk of contracting/spreading COVID-19).

Pre-mitigation risk:

Severity: 3. Likelihood: 2. Risk: 6 (Medium).

Mitigation:

A large proportion of patients are planned to continue attending their face-to-face clinic appointments in the diabetes centre (despite the ongoing pandemic) due to the complexities of diabetes care. Patients selected for participation in the research will be those who are already due to attend a face-to-face diabetes clinic appointment. Participants are reminded in letters from the hospital to avoid attendance if they are self-isolating or displaying symptoms of coronavirus. Regular hand-washing and sanitising are encouraged. The diabetes department has been adapted to take into account the latest Public Health England Guidance on social distancing and PPE. Everyone attending hospital is supplied with a surgical mask at site entrances. Mask wearing is mandatory unless specific written exemption certificates are demonstrated.

Risk rationale:

Patient attendance is required to enable the delivery of the study intervention (setting an explicit average blood glucose (A1c) target against their current A1c reading).

Post-mitigation risk:

Severity: 3. Likelihood: 1. Risk: 3 (Low).

9. Risk of research intervention.

Pre-mitigation risk:

Severity: 1. Likelihood: 3. Risk: 3 (Low)

Mitigation:

The research intervention is to set an explicit A1c target for the patient. It is common for patients to be set glycaemic targets as part of secondary care diabetes clinic procedures. Higher A1c targets could result in participants relaxing their glycaemic control. Lower A1c targets could result in participants tightening their glycaemic control. Their clinical management by a senior diabetes clinician will remain unchanged. By taking part in the feasibility study, the participants will have the benefit of two visits to see a diabetes clinician where they would commonly be seen only once. At the patient's last clinic visit, an appropriate A1c will be discussed with them to enable optimisation of their management following completion of the study.

Risk rationale:

The patient will have a study-set A1c target for only three months. Whether their target is set above or below their current A1c reading, the three-month window of A1c target alteration would be unlikely to have any impact on the patient's physical health.

Post-mitigation risk:

Severity: 1. Likelihood: 3. Risk: 3 (Low)

10. Interview is upsetting/distressing for the participant.

Pre-mitigation risk:

Severity: 3. Likelihood: 1. Risk: 3 (Low).

Mitigation: Distress protocol (Draucker, Martsolf and Poole, 2009) will be followed should any distressing issues arise during the interview process.

Post-mitigation risk:

Severity: 2. Likelihood: 1. Risk: 2 (Low).

Service user group feedback suggested that the study processes were acceptable and did not pose an unacceptable risk and burden.

RISKS TO RESEARCHER

1. Working in the diabetes department in-hospital will carry the risk of increased exposure to COVID-19.

Pre-mitigation risk:

Severity: 2. Likelihood: 2. Risk: 4 (Medium).

Mitigation:

The researcher already works in hospital – no increased risk from baseline. The researcher is trained and fully up-to-date with the use of PPE and the latest PHE guidance on social distancing and hand-washing. The researcher has been risk-scored for COVID-19 by STHK and has been deemed low risk, taking into account age, gender, ethnicity, shielding status, childcare arrangements, underlying health conditions and work environment.

Post-mitigation risk:

Severity: 1. Likelihood: 1. Risk: 1 (Low).

OTHER RISKS.

1. Identification and screening of participants (use of patient-identifiable data).

The national data-opt out service enables the public to opt-out of their confidential information being used for the purposes beyond their individual care and treatment. The public can change their national data opt-out choice at any time. National data opt-outs apply to a disclosure of confidential information when a researcher confirms they have approval from the Confidentiality Advisory Group (CAG) for use of patient-identifiable information, unless, as is the case with this project, the researcher is already an existing member of the study population's existing clinical care team.

All NHS trusts are compliant with the national data-opt out policy as of 31/3/2020.

Ethical approval by Edge Hill University Health Research Ethics Committee, and the Health Research Authority is required prior to research commencement. Use of contact details of patients who have been referred to the Department of Diabetes and Endocrinology at St Helens and Knowsley Teaching Hospitals NHS Trust (for the purposes of contacting a patient to consent) is allowed as the researcher is a member of the existing clinical care team.

Service user group feedback has been sought on the use of patient-identifiable information for the purposes of contacting potential research participants. General feedback from the group, including people with diabetes, was that contacting patients for the purpose of research recruitment (as a member of the patient's existing clinical care team) is justifiable as the knowledge gained from the research is hoped to improve patient care.

2. Consent.

Good clinical practice (GCP) guidelines will be followed throughout the study. The researcher has received online training and certification from the National Institute of Health Research (NIHR) on GCP.

The researcher works as a healthcare professional within St Helens and Knowsley Teaching Hospitals NHS Trust (research sponsor). The researcher will maintain clinical knowledge by undertaking routine clinical work in diabetes, endocrinology and general internal medicine. The researcher will maintain full registration with the General Medical Council with a licence to practice. The research will maintain medical indemnity with the Medical Defence Union.

##### 8.2 Research Ethics Committee (REC) and other Regulatory review & reports

Before the start of the study, a favourable opinion will be sought from Edge Hill University REC and the Health Research Authority (HRA).

##### 8.2.1 Regulatory Review & Compliance

Before any site can enrol patients into the study, the Chief Investigator/Principal Investigator or designee will ensure that appropriate approvals from participating organisations are in place. For any amendment to the study, the Chief Investigator or designee, in agreement with the sponsor (STHK) will submit information to the appropriate body in order for them to issue approval for the amendment. The Chief Investigator or designee will work with STHK Trust R&D so they can put the necessary arrangements in place to implement the amendment to confirm their support for the study as amended.

##### 8.2.2 Amendments

If a substantial amendment to the protocol or associated study documents is required, the sponsor must submit a valid notice of amendment to the REC for consideration. The REC will provide a response regarding the amendment within 35 days of receipt of the notice.

If applicable, other specialist review bodies (e.g. Edge Hill University Health Research Ethics Committee) need to be notified about substantial amendments in case the amendment affects their opinion of the study.

The requirement for amendments will be decided by the researcher and the researcher’s supervisory team. If amendments to the protocol or associated study documents is required, changes to the documents will be made by the research student with advice from the supervisory team. Changes will be communicated to the sponsor (STHK). It is the sponsor’s responsibility to decide whether an amendment is substantial or non-substantial for the purposes of submission to the REC.

Amendment history will be tracked. Study documents will be held in a hierarchical file system. Documents will be named in the following fashion:

'study title' - 'document name' - 'document version number (1.0.0)' - 'document creation/update date (YYYY-MM-DD)'.extension

Document version and date will be displayed in the file name and on each page of the document. Ratified master copies of the documents will be held as read-only copies on a cloud-shared NHS computer drive. Document version numbers are displayed as 'Major.Minor.Revision'. The first version of a document would be '1.0.0' with revisions displayed as '1.0.1', minor adaptations as '1.1.0' and major changes as '2.0.0'. Once updated documents have been ratified, old copies of the documents will be archived and the new copies will replace them in the master file. Document revisions will be recorded in a separate document.

#### 8.3 Peer review

Internal and external independent, proportionate (to the size and complexity of the study) and expert (reviewers should have knowledge of the relevant discipline and/or expertise to assess the methodological aspects of the study) peer review will take place and be submitted to the sponsor. The internal and external peer review documents are included in Section 11.1 'Appendix 1 – Required Documentation'. The internal peer reviewer (Consultant Physician/Diabetologist) approved the study protocol. The external peer reviewer (Professor of Medicine and Honorary Consultant Diabetologist) had a few minor comments and one major comment. These are summarised below, with accompanying amendments.

1. Clarify recruitment population and stratification (minor).
   - ⇒ Section 7.1.1 'Inclusion Criteria' updated.
2. Provide a stronger rationale for setting higher explicit HbA1c targets (major).
   - ⇒ Section 2 'Rationale' and section 5.1 'Study A - Study Design' updated.
3. Amend the HbA1c study leaflet infographic to include levels of HbA1c considered adequate by guidelines and non-diabetic HbA1c levels (minor).
   - ⇒ Section 11.1 'Appendix 1 – Required Documentation' HbA1c Leaflet amended.
4. Consider including healthcare professionals from outside STHK in interviews (minor).
   - ⇒ Following discussion with the supervisory team, it was felt that we could potentially include healthcare professionals if the study fails to recruit suitable numbers locally.

University research approval processes (research project registration) suggested the inclusion of additional qualitative aspects of the study in the event that poor recruitment is seen in the quantitative aspects of the project (Study A). Study D was added to the protocol to address these concerns.

#### 8.4 Patient & Public Involvement

Service user (SU) group feedback has been sought on the research proposal and the patient-facing study documents (participant information sheets, consent forms, leaflets, invitation letters). The service user group consists of a range of people from the general public, people with diabetes and expert patients. Feedback was received via email and video call. Documents have been updated with respect to the received comments:

- Comments suggested to clarify in the participant information sheet that taking part in research will not affect patient care. It is now explicit in the patient information sheets that involvement in the research does not affect their clinical care.

- One member on the SU group mentioned that the section on consent in the participant information sheets should be expanded upon to make it clear that individual consent is required and can be withdrawn at any time.
- Comments suggested evaluation of barriers to participation in those declining entry into the main study. 'Study D' has been added to the protocol to address this point.

Service user group feedback was sought on the acceptability of using patient identifiable data in the study without prior consent (namely for the purpose of contacting individuals to invite them to take part in the research). Feedback suggested that the use of patient-identifiable data for this purpose was reasonable. Specific comments:

- "would be happy to be contacted" (person with diabetes)
- "seems reasonable"
- "the invitation letter could improve recruitment if it comes from the consultant"
- "I wouldn't mind being contacted out of the blue for this project"
- "As long as it's highlighted that it is optional and not affecting healthcare"
- "it's important that you regularly update your participants as to the research progress and results"
- "it would be interesting if you followed-up non-responders to see why they didn't want to take part"

The SU group also suggested that it is important to regularly update people involved in the research. Following completion of the research, dissemination of the results to involved participants and the local diabetes service user group is planned.

##### 8.5 Protocol compliance

Publication of the protocol is planned to ensure greater transparency of the research process and also informs the research community about what research activity is currently being carried out in different fields, preventing unnecessary duplication of work and encouraging collaboration between research groups. Accidental protocol deviations can happen at any time. These will be adequately documented on the relevant forms and reported to the Chief Investigator and Sponsor immediately.

##### 8.6 Data protection and patient confidentiality

All data obtained will be anonymised for person- and place-identifiable information during transcription/database input by removal of direct identifiers. The secure holding of all data during the study period will be on password-protected NHS computers with NHS-level data encryption. Transfer of study data between computing devices will be either with an NHS-encrypted USB drive or a secure N3 (NHS network encrypted) email. Non-electronic data (e.g. consent forms, completed questionnaire) will be held in a secure file in a separate locked research office on NHS sites when not in use. Audio-recording data (.mp3/.wav) will be stored securely on a password-protected recorder device in a separate locked research office on NHS sites when not in use. Audio recordings will be held separately to signed consent form documents. All paper documentation will be held in a separate, locked file in a locked research office. Any detailed, identifiable free-text responses from surveys or semi-structured interviews will have their meaning generalised during transcription to prevent disclosure. Direct identifiers in all data obtained will be replaced with a code. Identifier code data will be located in a password-protected Microsoft Excel document on NHS-encrypted computers. Patient identifiers will be held in a separate, password-protected database and linked through a unique study code. Separation of patient identifiers from other data will be at the point of entry into the database.

University and Health Research Authority ethical approval will be obtained prior to recruitment/commencement of data collection.

##### 8.7 Indemnity

Sponsor indemnity: STHK NHS R&D indemnity.

Investigator indemnity: the researcher and chief investigator are individually indemnified personal medical indemnity.

##### 8.8 Access to the final study dataset

The final dataset will be accessed by the research team. All anonymised data collected could be of potential value for future research. As such, the collected, anonymised computer data will be submitted to the Edge Hill University research data repository for use upon completion and publication of the research. Participants will be consented for this.

Physical copies of data will be kept for the duration specified by St Helens and Knowsley Teaching Hospitals NHS Trust (STHK) Research Guidelines, at which point further retention of the data can be reviewed. Consent forms will be kept for as long as the research data are retained in the data repository. Following the completion of the research, they will be digitised and stored securely (encrypted) with the university. At this point, original physical copies will be destroyed securely (https://www.ukdataservice.ac.uk/manage-data/legal-ethical/consent-data-sharing/consent-forms).

##### 8.9 Progression criteria (go/no-go)

A green (proceed: there are no concerning issues that threaten the success of a future trial), amber (amend: where there are remediable issues, thereafter proceeding with caution), red (stop: there are intractable issues which cannot be remedied) system will be used to determine progression to a main trial.

Criteria informing the green/amber/red system will use:

a. Screening.
   ⇒ Ensuring a sufficient pool of patients to screen will be key in achieving the required sample.
   In STHK, six face-to-face diabetes clinics run per week with eight patients per clinic. Usually, at most, there are three ‘Did Not Attends’ (DNAs) or cancellations, leaving a usual minimum of five patients per clinic. This leaves 130 patients/month to screen.

b. Eligibility.
   ⇒ Determination of the expected eligibility rate of patients will allow for an estimation of the likelihood that investigators will have enough participants to recruit.
   For study A, it is estimated that 25% of screened patients will be eligible to participate in the study (cf. other diabetes trials) (Steiner et al., 2015, Neal et al., 2017, Wiviott et al., 2019). This means an estimated 32 patients/month will be eligible to study entry.

c. Recruitment.
   ⇒ Recruitment is the most commonly used criterion in establishing whether a feasibility study or pilot study should progress to a main trial. Suggested methods of determining adequate recruitment are to compare expected versus actual recruitment per centre per month.
   For this study, the recruitment period is planned to take place over a 4-month period. Fifty participants are required for the quantitative aspects of study A according to the protocol. Therefore a recruitment rate of 13 participants/month over four months will result in achieving the desired sample.

   It is estimated that 70% of eligible patients will agree to study enrolment (cf. other diabetes trials) (DAFNE Study Group, 2002, Davies et al., 2008). This means 22 patients/month of the 32 eligible patients are predicted to enrol.

d. Protocol non-adherence.
   ⇒ More commonly used in the clinical trials of investigational medicinal products (CTIMPs), the focus of evaluation of protocol non-adherence is on adherence to the trial intervention. Types of non-adherence are ‘cross-over’ or ‘off-protocol intervention’. Cross-over is where a participant does not receive their allocated intervention, instead receiving an alternate intervention. This will reduce the power of the study. Off-protocol intervention is where another intervention which is not included in the protocol is used. Non-adherence may arise due to side effects of the intervention or problems with the acceptability of the intervention.

In this study, cross-over and off-protocol intervention episodes will be monitored and recorded.

e. Outcome data.

⇒ Monitoring the quality and completeness of short-term outcome data provides valuable insight into issues which can be addressed for the main trial. Missing or incomplete data will impact upon the required sample size. Loss of data in this way can be due to attrition, acceptability of the outcome assessment protocol or the outcome measurement tools themselves. Generally, loss to follow-up should be < 20% to reduce threats to trial validity (Dettori, 2011). Greater than 60% of participants would need to provide response data to enable a trial to be powered to show an effect and reduce response bias (Fincham, 2008).

Considering these criteria, reasonable progression criteria (‘green’) for study A are:

- Identification of at least 19 eligible patients/month through screening.
- Recruitment of at least 13 patients/month.
- Monitoring of cross-over and off-protocol intervention episodes for significant protocol deviation.
- Retention of ≥ 80% at follow-up.
- Response rate of ≥ 60% for PROMs.

Failure to meet these criteria result in an amber situation. Identifiable remediable adjustments to allow trial progression will be identified. Failure to meet the progression criteria without the identification of remediable changes may result in a red situation where the trial will be prevented from progressing.

### 9 DISSEMINIATION POLICY

#### 9.1 Dissemination policy

On completion of the study, the data will be tabulated and analysed. Findings and discussions will form part of the researcher’s thesis document. As part of this process, publications in peer-reviewed journals are planned. The researcher has identified a number of suitable journals. Posters and abstracts are planned for conference submission. Following completion and publication of the research, the data will be owned by Edge Hill University. The data will be made available to Edge Hill University research data repository following the publication of the research results on completion of the PhD project.

### 11. APPENDICIES

#### 11.1 Appendix 1- Required documentation

The following pages contains the associated study documents. Documents can be found at the following links:

- PhD student researcher CV (link)
- Chief Investigator CV (link)
- Clinical/academic supervisor CV's (link)
- Peer review forms (link)
- Participant information sheets (link)
- Consent forms (link)
- HbA1c Leaflet (link)
- HbA1c Lay summary script (link)
- Letter templates (link)
- Email templates (link)
- Acceptability survey (link)
- Barriers to participation survey (link)
- GDRP patient information (link)
- Patient interview guide (link)
- Healthcare professional interview guide (link)
- Research data management plan (link)
- EQ-5D-5L HRQOL tool (link)
- Diabetes Empowerment Scale (DES-LF) tool (link)
- Summary of Diabetes Self-Care Activities (SDSCA) tool (link)
- Well-Being Questionnaire (WBQ-12) tool (link)
- Problem Areas in Diabetes tool (link)

**PhD student researcher CV**

**CURRICULUM VITAE**

| **Name: Samuel John Westall** | |
|---|---|
| **Present appointments:** | |
| Clinical Research Fellow, Department of Endocrinology and Diabetes, St Helens and Knowsley Teaching Hospitals NHS Trust<br><br>PhD Student Researcher, Faculty of Health, Social Care and Medicine, Edge Hill University | |
| **Address (work):** | |
| Diabetes Department<br>St Helens Hospital<br>Marshalls Cross Road<br>St Helens<br>WA9 3DA | |
| **Telephone number (work):** | **Email address (work):** |
| (+44) 01744 646249 | |
| **Qualifications:** | |
| MBChB (University of Liverpool), 2010<br>MRCP (UK), 2014<br>Diploma in Medical Leadership (Edge Hill University), 2018<br>MRCP (Diabetes and Endocrinology), 2019 | |
| **Professional registration:** | |
| General Medical Council, Registration number 7080659, full registration date 3/8/2011. | |
| **Previous and other appointments:** | |

| | | |
|---|---|---|
| Specialty Registrar | Mersey Deanery rotations in Diabetes/Endocrinology and Internal Medicine | 08/2016 to 03/2019<br>04/2020 to 08/2020 |
| Locum Medical Registrar | NHS Trusts in North West England | 08/2014 to 07/2016 |
| Expedition medic | Exile Medics and Operation Wallacea | 06/2015 to 08/2015 |
| Core Medical Training | Mersey Deanery rotations in Elderly Care, | 08/2012 to 07/2014 |

| | | |
|---|---|---|
| | Cardiology, Gastroenterology, Nephrology, Diabetes/Endocrinology, Respiratory | |
| Foundation Training | Mersey Deanery | 08/2010 to 07/2012 |

**Research experience:** *(Summary of research experience, including the extent of your involvement. Refer to any specific clinical or research experience relevant to the current application.)*

Study doctor in ORION-4 study (IRAS ID 240684, 2020 – ongoing)

- Responsible for inviting patients to screening and approval for randomisation

Co-CI in ACCREDIT study (IRAD ID 285093, 2020 – ongoing)

- Responsible for confirming eligibility, make study related medical decisions, make case report form entries/corrections, resolve data queries and maintain essential documents.

**Research training:** *(Details of any relevant training in the design or conduct of research, for example in the Clinical Trials Regulations, Good Clinical Practice, consent or other training appropriate to non-clinical research. Give the date of the training.)*

- Undertaken a period of core research training at Edge Hill University (2019-20) as part of an ongoing research PhD.
- Completed NIHR online certification in Good Clinical Practice (6/1/2020).

**Relevant publications:** *(Give references to all publications in the last two years plus other publications relevant to the current application.)*

**Westall, S.**, Sullivan, H., Rawsthorne, E., Beesley, C., Collins, J., Furlong, N., Narayanan, R. P. (2020, August). Ambulance alert system enabling pre-hospital parenteral steroids for patients with adrenal insufficiency. August 2020. In *22nd European Congress of Endocrinology* (Vol. 70). BioScientifica.

Sullivan, H., **Westall, S.**, McNulty, S., Furlong, N., Gilkes, C., Daousi, C., Narayanan, P. (2019). Nature's price for Cushing's disease: A blind eye and a hole in the (pituitary) Middle. April 2019. Endocrine Abstracts.

**Westall, S.,** Sullivan, H., McNulty, S., Bujawansa, S., & Narayanan, P. (2019, April). Partial cranial diabetes insipidus in breast cancer: invisible pituitary metastases or uncanny coincidence. April 2019. In *Society for Endocrinology Endocrine Update 2019* (Vol. 62). BioScientifica.

Heald A.H., Stedman M., **Westall S**., Kalansooriya A., Livingston M. Using population based real world experience and evidence in NHS England to identify opportunities to improve Type 2 diabetes outcomes and use of resources. Diabetic Medicine. Conference Abstract: Diabetes UK Professional Conference 2017. United Kingdom. 34 (Supplement 1) (pp 179), 2017.

| **Signature:** | **Date:** |
|---|---|
| 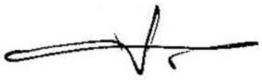 | 09/11/2020 |

#### Chief Investigator (Dr N Furlong) CV

**CURRICULUM VITAE**
**Dr N Furlong 03 March 2021**

**Name:**
Dr Niall Furlong

**Present appointment**
Consultant Physician and Diabetologist

**Address:**
Diabetes Centre, St Helens Hospital
St Helens & Knowsley Teaching Hospitals NHS Trust
Marshalls Cross Road
St Helens, Merseyside, WA9 3DA

| **Telephone number:** | **Email address:** |
|---|---|
| Work 01744 646500<br>Mobile 07803135077 | |

**Qualifications:**

| | |
|---|---|
| MB ChB | 1995 Liverpool |
| MD | 2005 Liverpool |
| FRCP | 2009 London |

**Professional Registration:**
GMC (full) 4190859 August 2005
GMC Revalidation Sept 2018 (due Sept 2023)

**Previous and other appointments:**
Consultant Physician and Diabetologist St Helens & Knowsley Teaching Hospitals NHS Trust (2005-presesent), including Clinical Director for Diabetes (2016-2018).

**Research experience:**
My MD (awarded 2005) focussed on insulin-oral hypoglycaemic agent combination therapy in Type 2 diabetes, including a systematic review with the Cochrane Collaboration & 2 prospective RCTs designed and conducted by myself.

Departmental lead for research (2005-); Principal Investigator for numerous multinational collaborative RCTs including major cardiovascular end-point studies; c £500K research income supporting multiple students (incl. PhD, MsC, BSc); c 20 original publications and 50 scientific abstracts; systematic reviewer with Cochrane collaboration, member of STHK Research, Development & Innovation group.

**Research training:**
Good Clinical Practice (GCP) Refresher: e-learning 16.09.2020

| **Signature:** | **Date:** |
|---|---|
| 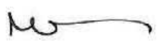 | **03 March 2021** |

Template CV for research applicants
Version 2, January 2007

#### Clinical/academic supervisor (Dr Prakash Narayanan) CV

##### CURRICULUM VITAE

**Name:** Dr Ram Prakash Narayanan

**Present appointment:**

1. Consultant Physician-Diabetes and Endocrinology/Internal Medicine, St Helens and Knowsley Teaching Hospitals NHS Trust (since Feb 2016)
2. Honorary Associate, Institute of Ageing and Chronic disease, University of Liverpool (since Feb 2016)

**Address:** Diabetes Centre,
St Helens Hospital,
Marshalls Cross Road,
St Helens,
Merseyside,
WA9 3DA

**Qualifications:**

MBBS (Nagpur University, India) 2003
MRCP(UK) 2008
PhD (The University of Manchester) 2013
FHEA (Higher Education Academy) 2014
MRCP (Diabetes and Endocrinology) 2016
CCT in Diabetes and Endocrinology/General Internal Medicine 2016
FRCP (London) 2019

**Professional registration:** General Medicine Council registration number 6064241

**Previous and other appointments:**

| | | |
|---|---|---|
| Clinical Lecturer | University of Liverpool/Aintree University Hospital | 01/2014 to 02/2016 |
| Specialty Registrar | Mersey Deanery rotations in Diabetes/Endocrinology and Internal Medicine | 08/2008 to 11/2009<br>11/2012 to 01/2014 |
| Clinical Research Fellow | The University of Manchester<br>Salford Royal Hospital | 11/2009 to 11/2012 |
| Senior House Officer | Arrowe Park Hospital | 08/2007 to 08/2008 |
| Senior House Officer | University Hospitals of Leicester | 08/2006 to 08/2007 |
| Foundation Year 2 | Carmarthenshire NHS Trust | 08/2005 to 08/2006 |
| Senior House Officer | Salford Royal Hospital | 01/2005 to 08/2005 |
| Locum House Officer | Salford Royal Hospital | 10/2004 to 11/2004 |
| SHO Intensive Care | Sundaram Medical Foundation, Chennai, India | 12/2003 to 4/2004 |
| Medical Officer | Jayaben Mody Hospital, Ankleshwar,India | 03/2003 to 08/2003 |
| Pre Reg House Officer | Mahatma Gandhi Institute of Medical Sciences, Wardha, India | 01/2002 to 01/2003 |

**Research experience:**

I have been awarded a PhD by The University of Manchester for my work studying genomic and proteomic interactions of the insulin-like growth factor system in type 2 diabetes. I have published upon longitudinal population trends in glycaemic control and cardiovascular risk factors in type 2 diabetes and researched associations of socioeconomic deprivation and painful neuropathy, as well as cardiovascular risk factors in patients with severe enduring mental illness.I am also researching associations of vitamin D with adipokines in obesity, as well as understanding adipokine changes to cytokine inhibition in rheumatoid arthritis.I have also been a sub-investigator in commercial trials in diabetes, endocrinology and renal medicine at Salford Royal Hospital.

**Research & other relevant training:** Good Clinical Practice (valid until Jan 2022)

**Relevant publications:**

Wong S, Yap YW, Narayanan RP, Al Jubouri M, Grossman A, Daousi CD, Mahgoub Y. Etomidate in the management of Cushing's disease and MRSA in a district general hospital in the UK. EDM Case Reports (Aug 2019)

Malipatil NS, Yadegarfar Y, Lunt M, Keevil B, Siddals K, Livingston M, Roberts S, Narayanan RP, Rutter M, Gibson JM, Donn R, Hackett G, Jones TH, Heald A. Male hypogonadism: 14 year prospective outcome in 550 men with type 2 diabetes. Endocrinol Diab Metab. 2019 Feb; e00064

Malipatil N, Lunt M, Narayanan RP, Siddals K, Cortés Moreno GY, Gibson MJ, Gu HF, Heald AH, Donn RP.Assessment of Global LINE1 DNA methylation in a longitudinal cohort of type 2 diabetes individuals. Int J Clin Pract. 2018 Oct 21:e13270

Steele T, Narayanan RP, James M, James J, Mazey N, Wilding JPH. Evaluation of Aintree LOSS, a community-based, multidisciplinary weight management service: outcomes and predictors of engagement. Clin Obes. 2017;7(6):368-376

Longworth H, McCallin K, Narayanan RP, Turner MA, Quenby S, Rycroft D, Charnley M, Abayomi J, Topping J, Weeks AD, Wilding JPH. Screening methods for obstructive sleep apnoea in severely obese pregnant women. Clin Obes. 2017; 7(4):239-244
Anderson SG, Narayanan RP, Radford D, Hodgson R, de Hert M, Heald AH. BMI independently relates to

glycaemia in patients with severe enduring mental illness. J Ment Health 2017; 26(3): 232-6

Anderson SG, Narayanan RP, Radford D, Hodgson R, de Hert M, Heald AH. BMI independently relates to glycaemia in patients with severe enduring mental illness. J Ment Health 2016; Aug 12(1-5).(epub ahead of print)

Heald AH, Martin JL, Payton A, Khalid L, Anderson SG, Narayanan RP, De Hert M, Yung A. Changes in metabolic parameters in patients with SMI over a 10 year follow-up period. Aust N Z J Psych 2016; Jan 14 (epub ahead of print)

Narayanan RP and Syed AA. Pregnancy following bariatric surgery-medical complications and management. Obes Surg. 2016 Oct;26(10):2523-9

Narayanan RP, Weeks AD, Quenby S, Rycrof D, Hart A, Longworth H, Charnley M, Abayomi J, Topping J, Turner MA, Wilding JPH. Fit for Birth- the effect of weight changes in obese pregnant women on maternal and neonatal outcomes: a pilot prospective cohort study. Clin Obes. 2016; 6(1):79-88

Livingston M, Robinson JC, Brown CE, Narayanan RP, Holland D, Fryer AA, Heald AH. Are cholesterol levels being checked and managed appropriately in UK primary care type 2 diabetes? Int J Clin Prac 69(11):1389-91

Narayanan RP, Samad S. Improving outcomes in diabetes in pregnancy. Practitioner 2015; 259(1782):25-28

Anderson SG, Narayanan RP, Malipatil N, Roberts H, Dunn G, Heald AH. Socioeconomic deprivation independently predicts symptomatic painful diabetic neuropathy in type 2 diabetes. Exp Clin Endocrinol Diabetes. 2015; 123(7):423-7

Narayanan RP, Fu B, Oliver RL, Siddals KW, Donn RP, Hudson JE, White A, Laing I, Ollier WER, Heald AH, Gibson JM. IGF-II and IGFBP-2 predict longitudinal elevations of HDL cholesterol in type 2 diabetes. Ann Clin Biochem 2014; 51(4):468-75

Narayanan RP, Fu B, Heald AH, Payton A, Hudson JE, Oliver RL, Anderson SG, Siddals KW, White A, Ollier WER, Gibson JM. IGF2 gene polymorphisms and IGF-II concentration are determinants of longitudinal weight trends in Type 2 diabetes. Exp Clin Endocrinol Diabetes 2013;121(6):361-7

Narayanan RP, Siddals KW, Heald AH, Gibson JM. Interactions of the IGF system with diabetes and its vascular complications. Exp Clin Endocrinol Diabetes 2013;121(5):255-61

Narayanan RP, Gittins M, Siddals KW, Oliver RL, Hudson JE, White A, Durrington P, Davies RR, Rutter MK, Gibson JM. Atorvastatin administration is associated with dose-related changes in IGF bioavailability. Eur J Endocrinol 2013; 168 (4): 543-8

Narayanan RP, Fu B, Heald AH, Siddals KW, Oliver RL, Hudson JE, Payton A, Anderson SG, White A, Ollier WER and Gibson JM. IGFBP2 is a biomarker for predicting longitudinal deterioration in renal function in type 2 diabetes. Endocr Connect 2012; 1(2): 95-102.

Anderson SG, Narayanan RP, Amlesh J, Heald AH. Type 1 diabetes in Cheshire: cardiometabolic risk factor trends. Prim Care Diabetes 2012; 6(2):123-6.

Anderson SG, Heald AH, Younger N, Bujawansa S, Narayanan RP, McCulloch A, Jones H. Screening for hypogonadism in diabetes: 2008/09: Results from the Cheshire Primary Care cohort. Prim Care Diabetes 2012; 6(2):143-8.

Narayanan RP, Bujawansa S, Qureshi MZ, Rimmer M, Heald AH. Hypogonadism secondary to hyperprolactinaemia: Successful treatment but adverse consequences. Exp Clin Endocrinol Diabetes 2012; 120(5):311-3.

Narayanan RP, Mason JM, Taylor J, Long AF, Gambling T, New JP, Gibson JM, Young RJ. Telemedicine to improve glycaemic control: 3 yr results from the PACCTS trial. Diabet Medicine 2012; 29(2):284-5.

Narayanan RP, Peasley M. Managing diabetic emergencies. Pharmacy Professional 2010;(6):36-40.

Narayanan RP, Kirk P, Lewis S. Uptake and perceptions of an e-learning package on blood transfusion by trainees in Wales. JR Coll of Physic Edinb 2008; 38: 298-301

Narayanan RP, James N, Ramachandran K, Jaramillo MJ. Guillain-Barre' syndrome presenting with bilateral facial nerve palsy - a case report. Cases J 2008; 1(1):379 and reproduced with permission in Progress in Neurology and Psychiatry 2009; 13(4): 06-09. Narayanan RP. Medical students leading social revolutions. The Clinical Teacher 2006; 3 (1):69-70

#### Student Director of Studies (Edge Hill University) (Dr S Watmough) CV

**CURRICULUM VITAE**

| | |
|---|---|
| **Name: Dr Simon Watmough** | |
| **Present appointment:** *(Job title, department, and organisation.)* | |
| Associate Head of Undergraduate Medicine and MBChB Programme Lead<br>School of Medicine<br>Edge Hill University | |
| **Address:** *(Full work address.)* | |
| Faculty of Health, Social Care and Medicine Rm 218<br>Edge Hill University<br>St Helens Road<br>Ormskirk<br>L39 4QP | |
| **Telephone number:** | **Email address:** |
| **01695-657043** | |
| **Qualifications:** | |
| BA History and Politics 2.1 – University of Southampton (June 1995)<br>MA Historical Research – University of Liverpool (December 1996)<br>PhD in Medical Education – University of Liverpool (July 2008)<br>Senior Fellow of the Higher Education Academy (July 2016) | |
| **Professional registration:** *(Name of body, registration number and date of registration.)* | |
| n/a | |
| **Previous and other appointments:** *(Include previous appointments in the last 5 years and other current appointments.)* | |
| Curent appointment Associate Head of Undergraduate Medicine and MBChB programme lead Edge Hill University from January 2020<br>Senior Lecturer in Medical Education and MBChB Programme Lead Edge Hill University June 2018 – December 2019<br>Senior Lecturer in Medical Education and Master in Medicine Programme Lead Edge Hill University March 2017 – June 2018<br>Lecturer in Health Cared Education – University of Liverpool Edge Hill University December 2015 – March 2017 | |
| Experience:<br>• have been involved in a variety of research projects including the influence of undergraduate and postgraduate medical education on competencies, medical careers, Physician Associate Education, simulation in medical education, Learning technology<br>• approx. 40 peer reviewed publications, approx. 30 conference abstracts, 1 editor of a book<br>• one of these research projects was as primary supervisors of a pharmacist from STHK undertaking a PhD on prescribing errors which led to a number of peer reviewed publications and conference abstracts. This involved extensive mixed methods research including a randomised control trial and involved pharmacists and al grades of doctors . This research with STHK is particularly relevant to the current application.<br>• supervised 1 PhD and 1 Mres to completion as primary supervisor and 8 PG dissertations.<br>• currently primary supervisor for 2 PhD students.<br>• taught research and ethics to both undergraduate medical students and postgraduate doctors in training over a 15 year teaching career<br>• Represent the Faculty of Health Social Care and Medicine on the Edge Hill University Research and Ethics Sub Committee and previously was a member of the Faculty of Health , Social Care and Medicine ethics committee and The University of Liverpool School of Medicine Ethics Committee. | |

**Research training:** *(Details of any relevant training in the design or conduct of research, for example in the Clinical Trials Regulations, Good Clinical Practice, consent or other training appropriate to non-clinical research. Give the date of the training.)*

Undertook extensive training in both qualitative and quantitative research methods for my PhD in medical Education at the University of Liverpool (2002-2008)
Trained in the HTA at Edge Hill University January 2018.

**Relevant publications:** *(Give references to all publications in the last two years plus other publications relevant to the current application.)*

Lloyd, M., Watmough, S. D., O'Brien, S. V., Hardy, K. & Furlong, N., Evaluating the impact of a pharmacist-led prescribing feedback intervention on prescribing errors in a hospital setting
16 Dec 2020, In: Research in Social and Administrative Pharmacy.
SandarsJ, Brown J, Nwolisa C, Patel M, Dogra N,Kaehne A, Garner J, Watmough S, Maden M, Duckworth V (2020) Can qualitative research help us understand the challenges of understanding equity in Medical Education? A scoping review of the literature *The Clinical Teacher* 17: (supplement 1) 66-67
https://doi.org/10.1111/tct.13238

SandarsJ, Brown J, Nwolisa C, Patel M, Dogra N,Kaehne A, Garner J, Watmough S, Maden M, Duckworth V The challenge of conducting qualitative research to understand the factors that influence equity in medical education: A scoping review
30 Apr 2020, In : MedEdPublish.

Lloyd, M, Watmough, S Bennett, (2018) Simulation-based training: applications in clinical pharmacy. *Clinical Pharmacist*, 10 (9). pp. 3-10. ISSN 1758-9061
DOI https://doi.org/10.1211/CP.2018.20205302

Lloyd, M., Watmough, Simon, O'Brien, S.V., Furlong, N. and Hardy, K. (2018)
Exploring the impact of pharmacist-led feedback on prescribing behaviour: A qualitative study. *Research in Social & Administrative Pharmacy*. 2018 Jun;14(6):545-554. doi: 10.1016/j.sapharm.2017.06.010. Epub 2017 Jun 29

Lloyd, Michael, Watmough, Simon, O'Brien, Sarah, Furlong, Niall and Hardy, Kevin (2017)
Exploring the impact of feedback on prescribing error rates: A pilot study.
*International Journal of Clinical Pharmacy*. pp. 1-5. ISSN 2210-7703 DOI 10.1007/s11096-017-0503-x

Lloyd, M, Watmough S, O'Brien S, Furlong N, Hardy K. Exploring Attitudes and Opinions of Pharmacists
towards delivering Prescribing Error Feedback : A Qualitative Case Study using focus group interviews. *Research in Social & Administrative Pharmacy* 2016 12(3) 761-74

Watmough, S, Box H, Bennett N, Stewart, S. Unexpected Medical Undergraduate Simulation Training (UMUST): Can unexpected medical simulation scenarios help prepare medical students for the Transition to Foundation doctor. *BMC Medical Education* 2016 16:110

Lloyd M, Watmough SD, O'Brien SV, Furlong N, Hardy K. How to give and receive constructive feedback. *The Pharmaceutical Journal,* 2016; Vol 296, No 7887,
online | DOI: 10.1211/PJ.2016.20200756

| **Signature:** | **Date:** |
|---|---|
| | 3/03/2021 |

#### Academic supervisor (Dr G Irving) CV

GREG JAMES IRVING

| | | |
|---|---|---|
| Address: | 65 Millbrook Lance, Eccleston, St.Helens, Merseyside, WA10 4QZ | |
| Phone number: | 07745148864 | |
| Email address: | | |
| DOB: | 10.10.80 | |
| Nationality: | British | |
| GMC No: | 6101830 | |
| Education: | The University of Liverpool, Liverpool, UK | (2007 - 2014) |
| | The University of Oxford, Oxford, UK | (2011 - 2013) |
| | The University of Nottingham, Nottingham, UK | (1999 – 2004 |
| Qualifications: | MA | (2017) |
| | FRCGP | (2017) |
| | PhD | (2015) |
| | MSc in Evidence Based Health Care | (2014) |
| | MPH | (2011) |
| | MRCGP | (2010) |
| | DRCOG | (2008) |
| | DFPSRH | (2008) |
| | Bachelor of Medicine / Surgery: | (2004) |
| | BMedSci (First class Honours): | (2002) |

Fellowships / Scholarships:

NIHR Clinical Lecturer in General Practice – University of Cambridge
NIHR Doctoral Research Fellow (GP): Personal award £339,763
Harvard Global Clinical Scholar Research Training Program

Appointments:

| | |
|---|---|
| Dec 15- Dec 19: | University of Cambridge |
| Oct 10 – Dec 15: | University of Liverpool – Senior Research Fellow |
| Aug 10 - Oct 10: | WHO (Geneva): Evidence and research cluster |
| Aug 07 – Aug 10: | Mersey deanery Academic Clinical Fellow - GP |
| Aug 06 – Aug 07: | South Manchester teaching hospitals (GP VTS) |
| Aug 05 – Aug 06: | Sheffield Teaching hospitals Foundation year 2 |
| Aug 04 – Aug 05: | Nottingham teaching hospitals Foundation year 1 |

Prizes:

SAPC Yvonne Carter award for outstanding new primary care new researcher (2013)
RCGP/Vasco Da Gamma - UK junior researcher of the year (2011)
RCGP Specialty Training award (2009)
Mersey RCGP General Practice team of the year (2009)
RCGP Great Expectations bursary (2009)
BMedSci Research award - Department of Infectious diseases (2002)
General Practice Audit prize, University of Nottingham (2001)
Certificate of merits in General Practice, University of Nottingham (2001)

Publications: https://research.edgehill.ac.uk/en/persons/greg-irving

Professional memberships:

Royal College of General Practitioners
Society for Academic Primary Care

#### Checklist for Internal Peer Review of Research Protocol Prior to Submission for STHK Sponsorship

| | |
|---|---|
| **Title of Study** | HbA1c TargeT AchIevemeNt in diabeteS study (ATTAINS study) |
| **Protocol version and date** | Version 2.0.2, dated 23rd October 2020 |

**Key Points to be considered**

- **What is already known about the topic? What value will this study add?**
- **Do the researchers have sufficient resources to complete the study effectively?**

| CONTENTS | CRITERIA | YES | NO |
|---|---|---|---|
| **Research Team** | Is the name of the principle researcher, qualifications and experience, together with those of other contributors given? | ✓ | |
| **Title** | Is the Title clear and succinct? | ✓ | |
| **Summary or Abstract** | This should be of 250-300 words giving a synopsis of the study including the design, methods of data collection and expected outcomes. | ✓ | |
| **Background** | This should make clear the health issue to be addressed, prevalence of the problem, the main clinical and conceptual issues and the "fit" of the research within the framework of existing knowledge and practice. Is a brief literature review with references included? | ✓ | |
| **Aims/Rationale** | Are the broad aims, primary and secondary (if applicable) objectives clearly stated? | ✓ | |
| **Design** | Is the methodology, type of study, numbers of subjects, inclusion/exclusion criteria, data collection etc clear? | ✓ | |
| **Statistician or Health Economist** | Is there evidence that appropriate advice has been sought at the design stage of the study to ensure its validity? | ✓ | |
| **Ethical Considerations** | Have ethical issues been considered and plans made to obtain ethical approval.? | ✓ | |
| **Intellectual Property** | Has the potential for IP exploitation been considered and advice sought? | ✓ | |
| **Expected Benefits** | Are these identified? E.g.: - More effective/efficient health care, improved knowledge, new research. | ✓ | |
| **Resources Required** | Have costs been considered?<br>RD&I costs – of the research itself and research governance<br>Service Support costs – additional tests, staff time etc<br>Excess Treatment costs | ✓ | |
| **Funding** | Is the source and amount of funding available to support the study indicated? | ✓ | |

**Name of Person undertaking the review:** DR STEVEN JOHN [illegible]

**Professional Title/Area of Expertise:** CONSULTANT PHYSICIAN / DIABETOLOGIST

**Please add any further comments on an additional page if required.**

*I have reviewed the protocol for the above study and have the following comments:*

IMPACT OF CHANGING HbA1c TARGETS ON ~~PTS~~ PSYCHOLOGICAL HEALTH.

STATISTICIAN SUPPORT WITH PHILIP [illegible]

; ANY HEALTH IMPACT OF HbA1c TARGET CHANGE LEADING TO ALTERATION OF HbA1c ATTAINED (UNLIKELY) PLUS SHORT TERM STUDY, HbA1c CHANGE (IF HAPPENS) ONLY FOR MONTHS

GOOD STUDY.

APPROVED ✓

| Date: | Signature: |
|---|---|

**Checklist for External Peer Review of Research Protocol**
**Prior to Submission for STHK Sponsorship**

| | |
|---|---|
| **Title of Study** | HbA1c TargeT AchIevemeNt in diabeteS study (ATTAINS study) |
| **Protocol version and date** | Version 2.0.2, dated 23rd October 2020 |

**Key Points to be considered**

- **What is already known about the topic? What value will this study add?**
- **Do the researchers have sufficient resources to complete the study effectively?**

| CONTENTS | CRITERIA | YES | NO |
|---|---|---|---|
| **Research Team** | Is the name of the principle researcher, qualifications and experience, together with those of other contributors given? | ✔ | |
| **Title** | Is the Title clear and succinct? | ✔ | |
| **Summary or Abstract** | This should be of 250-300 words giving a synopsis of the study including the design, methods of data collection and expected outcomes. | ✔ | |
| **Background** | This should make clear the health issue to be addressed, prevalence of the problem, the main clinical and conceptual issues and the "fit" of the research within the framework of existing knowledge and practice. Is a brief literature review with references included? | ✔ | |
| **Aims/Rationale** | Are the broad aims, primary and secondary (if applicable) objectives clearly stated? | ✔ | |
| **Design** | Is the methodology, type of study, numbers of subjects, inclusion/exclusion criteria, data collection etc clear? | ✔ | |
| **Statistician or Health Economist** | Is there evidence that appropriate advice has been sought at the design stage of the study to ensure its validity? | ✔ | |
| **Ethical Considerations** | Have ethical issues been considered and plans made to obtain ethical approval.? | ✔ | |
| **Intellectual Property** | Has the potential for IP exploitation been considered and advice sought? | ✔ | |
| **Expected Benefits** | Are these identified? E.g.: - More effective/efficient health care, improved knowledge, new research. | ✔ | |
| **Resources Required** | Have costs been considered?<br>RD&I costs – of the research itself and research governance<br>Service Support costs – additional tests, staff time etc<br>Excess Treatment costs | ✔ | |
| **Funding** | Is the source and amount of funding available to support the study indicated? | ✔ | |

**Name of Person undertaking the review:** Professor John Wilding

**Professional Title/Area of Expertise:** Professor of Medicine and Honorary Consultant in Diabe

**Please add any further comments on an additional page if required.**

*I have reviewed the protocol for the above study and have the following comments:*

This is an interesting study and protocol is generally clear and well written. I do have a few minor comments and one major comment:

CV for CI and co-investigators are missing.

Please clarify if the study is recruiting people with type 1 diabetes, type 2 diabetes or all types of diabetes. The treatment is very different and I think if all types of diabetes are being considered then it will be important to look at them separately with some stratification by diabetes type.

It is not quite clear in the protocol why targets above current HbA1c control are being used. Most patients are referred to hospital services for poor glucose control and HbA1c is often very high at point of referral. I would have preferred to have seen a modest target (5mmol/mol drop in HbA1c) vs a more stringent one (> 10mmol/mol); this would to me seem a more realistic approach. I suggest providing a stronger rationale for these higher targets or change them as suggested above.
It is not quite clear to me how the usual clinicians will be providing advice that is consistent with the targets.

The HbA1c target graph in the PIS does not go down to the level of control considered adequate by many guidelines (<53mmol/mol) - this needs to be changed to include the non-diabetic range (ie down to 48 mmol/mol and lower)

I would consider going outside a single Trust for objective C - it is likely that all the team in the St Helen's Trust will be familiar with the study, so their input may not be representative.

| Date: 03/DEC/20 | Signature: [signature] |
|---|---|

### HbA1c TargeT AchIevemeNt in diabeteS study (ATTAINS study) – Study A

Researcher: Dr Sam Westall Chief Investigator: Dr Niall Furlong

#### Introduction

You have been invited to take part in a study because you have diabetes and you are coming to St Helens Hospital diabetes clinic. Before deciding if you want to take part, it is important that you know why. Please read this sheet and talk with friends and family if you want. Ask us if there is anything that is not clear. Take time to decide if you want to take part. **Thank you for reading this.**

#### Important things you need to know

- Your usual diabetes care and the care you receive from the NHS will not be affected by taking part.
- If you need an appointment with your GP, you should continue to book this in the normal way.
- At Edge Hill, we are committed to protecting your personal information. To find out ways that we use your data, please see www.edgehill.ac.uk/about/legal/privacy

#### What do are we doing about COVID-19 (coronavirus)?

We are continuing to do research as we think it is important. Edge Hill University and the NHS are taking the necessary precautions to ensure the safety of research participants, staff and students.
**If you have symptoms of coronavirus or are self-isolating, you must not come to hospital**. Call 111 or visit 111.nhs.uk/covid-19/ for further advice. Please wear the masks provided and sanitise hands on arrival. The coronavirus pandemic is rapidly evolving, please visit gov.uk/coronavirus for up-to-date guidance.

#### What is the study about?

This is part of a group of studies testing the achievement of blood sugar targets (HbA1c) in diabetes. We are doing this to test if a bigger study is justified in the future. We also want to see what the effect of setting higher ('Group A') or lower ('Group B) average blood sugar ('A1c') targets has on your well-being and blood sugar control. We will test this by randomising (like tossing a coin) participants into each group. There is a chance that you A1c may change over the study period. We do not expect this to have an effect on your health. Your participation could help make diabetes care better for the future.

#### What does this study involve?

We want to see you three times over six months:

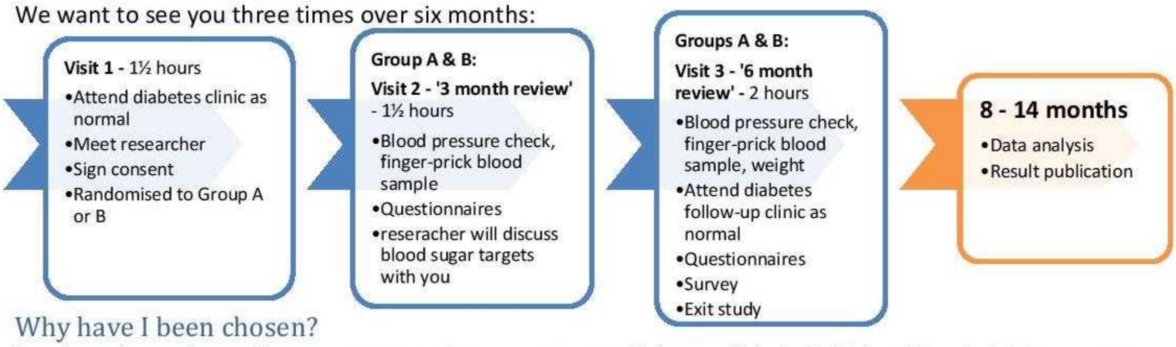

#### Why have I been chosen?

You have been chosen because you are due to come to a diabetes clinic in St Helens Hospital. To get more information on being chosen for research, visit www.nhs.uk/your-nhs-data-matters or call 0300 303 5678.

#### Do I have to take part?

It is up to you. If you want to take part, you will be given this information sheet to keep. We will ask each participant to sign a consent form. You can withdraw your consent and stop taking part at any time.

#### I am happy to take part, what do I do next?

The researcher will contact you by phone before your clinic to see if you are interested. We will meet you when you come to clinic to check that you are happy and eligible to take part. We will organise a follow-up visit at a time that suits you.

#### What if I agree to take part and change my mind?

You can withdraw from the study at any time, without giving a reason. Your diabetes care will not be affected. If you withdraw, we will no longer collect any information about you or from you. We will keep information about you that we have already obtained as this information may have already been used in some analyses. If this is the case, we will use the minimum personally identifiable information.

#### What are the possible advantages and disadvantages of taking part?

The research is not meant to provide you with a direct benefit. However, by taking part, you will add to our knowledge which could help patients in future. We will ask to see you three times over 6 months. This may take a few hours of your time.

#### Expenses and/or payments: No expenses or payments will be available.

#### What if something goes wrong/who can I complain to?

If something goes wrong you can email the researchers. Alternatively, you can send us a letter (address below). If this does not achieve a suitable result, you should contact the Research Governance Officer. If you do this, please give the name of the study and the IRAS ID (at the top the page).

#### Will my participation in the study be confidential?

All data about you will be kept securely on NHS premises. Any information you give will be anonymised before publication. Your data will be given a code which will be used instead of your name. Any identifiable information you may give will be anonymised when collected and nothing identifiable will be used in publications. We will let your GP know that you are taking part. In some cases, if something important related to your health comes up, we might have to let your GP know. To continue supporting future research, anonymised data will be made available for other research groups to use. Where appropriate and necessary, regulatory inspectors or NHS research offices will be allowed access to your personal data to verify the study results.

#### What will happen to the results of this study?

The results of the study may be printed in medical journals, reports and presentations.

#### Who is organising and funding the research? Who has reviewed this study?

This research is funded by the Diabetes Department, St Helens Hospital. Research is supervised by Edge Hill University. This study has been approved by University Research Ethics Committee and the Health Research Authority.

Address:
ATTAINS Study, Diabetes Centre, St Helens Hospital, Marshalls Cross Road, St Helens, WA9 3DA

**Thank you** for volunteering to take part in this study

For further information, contact: Dr Sam Westall

NHS Trust

**Title of project:**

### HbA1c TargeT AchIevemeNt in diabeteS study
#### (ATTAINS study) – Study B

Researcher: Dr Sam Westall

Chief Investigator: Dr Niall Furlong

##### Introduction

You have been invited to have an interview. Before you choose to take part, it is important that you know why. Please read this sheet and talk with friends and family if you want. Ask us if there is anything that is not clear. Take time to decide if you want to take part. .

**Thank you for reading this.**

##### Important things you need to know

- Your usual diabetes care and the care you receive from the NHS will not be affected by taking part.
- If you need an appointment with your GP, you should continue to book this in the normal way.
- At Edge Hill, we are committed to protecting your personal information. To find out ways that we use your data, please see www.edgehill.ac.uk/about/legal/privacy

##### What do are we doing about COVID-19 (coronavirus)?

We are continuing to do research as we think it is important. Edge Hill University and the NHS are taking the necessary precautions to ensure the safety of research participants, staff and students.

**If you have symptoms of coronavirus or are self-isolating, you must not come to hospital**. Call 111 or visit 111.nhs.uk/covid-19/ for further advice. Please wear the masks provided and sanitise hands on arrival. The coronavirus pandemic is rapidly evolving, please visit gov.uk/coronavirus for up-to-date guidance.

##### What is the study about?

This is part of a group of studies testing the achievement of blood sugar targets (HbA1c) in diabetes. We are doing this to test if a bigger study is justified in the future. We want to get your views on the use of treatment goals in diabetes. Your participation could help make diabetes care better for the future.

##### What does this study involve?

You are being asked to take part in a recorded interview. The interview will be like a conversation. The interviewer will ask you about your views on blood sugar targets in diabetes. The interview will take about 30 minutes. We will ask you if you would be happy for us to call for an interview you during your visits to the diabetes centre. If you are happy to have an interview, we will ask you to sign a consent form. We will organise a convenient time to call you.

##### Why have I been chosen?

You have been chosen because you are due to come to a diabetes clinic in St Helens Hospital. To get more information on being chosen for research, visit www.nhs.uk/your-nhs-data-matters or call 0300 303 5678.

##### Do I have to take part?

It is up to you. If you want to take part, you will be given this information sheet to keep. We will ask each participant to sign a consent form. You can withdraw your consent and stop taking part at any time.

##### I am happy to take part, what do I do next?

The researcher will have asked you if you would like to take part during your visit to the diabetes centre. The researcher will contact you by phone at the time you agreed upon for the interview.

##### What if I agree to take part and change my mind?

You can withdraw from the study at any time, without giving a reason. Your diabetes care will not be affected. If you withdraw, we will no longer collect any information about you or from you. We will keep information about you that we have already obtained as this information may have already been used in some analyses. If this is the case, we will use the minimum personally identifiable information.

##### What are the possible disadvantages of taking part?

There are no disadvantages to taking apart. All we will ask for is a little bit of your time for an interview.

##### What are the possible benefits of taking part?

The research is not meant to provide you with a direct benefit. We genuinely believe that ongoing research in diabetes is important. By taking part, you will add to our knowledge which could help patients in future.

##### Expenses and/or payments.

No expenses or payments will be available.

##### What if something goes wrong/who can I complain to?

If something goes wrong, you can email the interviewer. Alternatively, you can send us a letter (address below). If this does not achieve a suitable result, you should contact the Research Governance Officer. If you do this, please give the name of the study and the IRAS ID (at the top the page).

##### Will my participation in the study be confidential?

All data about you will be kept securely on NHS premises. Any information you give will be anonymised before publication. Your data will be given a code which will be used instead of your name. Any identifiable information you may give will be anonymised when collected and nothing identifiable will be used in publications. We will let your GP know that you are taking part. In some cases, if something important related to your health comes up, we might have to let your GP know. To continue supporting future research, anonymised data will be made available for other research groups to use. Where appropriate and necessary, regulatory inspectors or NHS research offices will be allowed access to your personal data to verify the study results.

##### What will happen to the results of this study?

The results of the study may be printed in medical journals, reports and presentations.

##### Who is organising and funding the research? Who has reviewed this study?

This research is funded by the Diabetes Department, St Helens Hospital. Research is supervised by Edge Hill University. This study has been approved by University Research Ethics Committee and the Health Research Authority.

Address:
ATTAINS Study, Diabetes Centre, St Helens Hospital, Marshalls Cross Road, St Helens, WA9 3DA

**Thank you** for volunteering to take part in this study

For further information, contact: Dr Sam Westall

Participant Information Sheet – Study C
Document version 1.0.1 – 23/10/20 IRAS ID: 291254
Title of project:

### HbA1c TargeT AchIevemeNt in diabeteS study
#### (ATTAINS study) – Study C

Researcher: Dr Sam Westall Chief Investigator: Dr Niall Furlong


##### Introduction

You have been invited to have an interview. Before you choose to take part, it is important that you know why. Please read this sheet and talk with friends, relatives and colleagues if you want. Ask us if there is anything that is not clear. Take time to decide if you want to take part. **Thank you for reading this.**

##### Important things you need to know

At Edge Hill, we are committed to protecting your personal information. To find out ways that we use your data, please see www.edgehill.ac.uk/about/legal/privacy

##### What do are we doing about COVID-19 (coronavirus)?

We are continuing to do research as we think it is important. Edge Hill University and the NHS are taking the necessary precautions to ensure the safety of research participants, staff and students.
**If you have symptoms of coronavirus or are self-isolating, you must not come to hospital**. Call 111 or visit 111.nhs.uk/covid-19/ for further advice. Please wear the masks provided and sanitise hands on arrival. The coronavirus pandemic is rapidly evolving, please visit gov.uk/coronavirus for up-to-date guidance.

##### What is the study about?

Individualised treatment targets are commonly used in UK secondary diabetes services as part of a personalised care plan for people with diabetes. Generic and diabetes-specific mental health issues are a known driver of poor blood glucose target achievement and increased morbidity and mortality. Little is known of the reciprocal effect that blood glucose target-setting has on the psychology and emotional health of people with diabetes. In a series of related studies, we are testing the feasibility of evaluating psychometric outcomes when using explicit blood glucose targets in people with diabetes. As part of these studies, we would like to interview healthcare professionals to understand their experience, views and opinions on the topic of glycaemic target-setting and individualised treatment targets.

##### What does this study involve?

You are being asked to take part in a recorded interview. The interview will take about 30 minutes. The interview will be via telephone or video call. We will organise a convenient date and time with you.

##### Why have I been chosen?

You have been invited to take part because you are a healthcare professional treating people with diabetes.

##### Do I have to take part?

It is up to you. If you want to take part, you will be given this information sheet to keep. We will ask each participant to sign a consent form. You can withdraw your consent and stop taking part at any time.

Participant Information Sheet – Study C
Document version 1.0.1 – 23/10/20 IRAS ID: 291254

##### I am happy to take part, what do I do next?

Please complete the consent form (electronically) which was attached to the email invite. Forward this to the researcher by email. The researcher will organise a convenient time for the interview.

##### What if I agree to take part and change my mind?

You can withdraw from the study at any time, without giving a reason. If you withdraw, we will no longer collect any information about you or from you. We will keep information about you that we have already obtained as this information may have already been used in some analyses and may still be used in the final study analyses. If this is the case, we will use the minimum personally-identifiable information.

##### What are the possible disadvantages of taking part?

There are no disadvantages to taking apart. All we will ask for is a little bit of your time for an interview.

##### What are the possible benefits of taking part?

The research is not meant to provide you with a direct benefit. We genuinely believe that ongoing research in diabetes is important. By taking part, you will add to our knowledge which could help patients in future.

##### Expenses and/or payments.

No expenses or payments will be available.

##### What if something goes wrong/who can I complain to?

If something goes wrong, you can email the interviewer. If this does not achieve a suitable result, you should contact the Research Governance Officer. If you do this, please give the name of the study and the IRAS ID.

##### Will my participation in the study be confidential?

All data about you will be kept securely on NHS premises. Any information you give will be anonymised before publication. Your data will be given a code which will be used instead of your name. Any identifiable information you may give will be anonymised when collected and nothing identifiable will be used in publications. To continue supporting future research, anonymised data will be made available for other research groups to use. Where appropriate and necessary, regulatory inspectors or NHS research offices will be allowed access to your personal data to verify the study results.

##### What will happen to the results of this study?

The results of the study may be printed in medical journals, reports and presentations. A short summary of findings will be made available to you upon request and you will be invited to a presentation of the findings.

##### Who is organising and funding the research?

This research is funded by the Diabetes Department, St Helens Hospital. Research is supervised by Edge Hill University.

##### Who has reviewed this study?

This study has been approved by University Research Ethics Committee and the Health Research Authority.

**Thank you** for agreeing to take part in this study v1.0.1 - 2020-10-23

Title of project:

### HbA1c TargeT AchIevemeNt in diabeteS study
#### (ATTAINS study) – Study D

Researcher: Dr Sam Westall

Chief Investigator: Dr Niall Furlong

#### Introduction

You have been invited to have an interview. Before you choose to take part, it is important that you know why. Please read this sheet and talk with friends and family if you want. Ask us if there is anything that is not clear. Take time to decide if you want to take part. **Thank you for reading this.**

#### Important things you need to know

- Your usual diabetes care and the care you receive from the NHS will not be affected by taking part.
- If you need an appointment with your GP, you should continue to book this in the normal way.
- At Edge Hill, we are committed to protecting your personal information. To find out ways that we use your data, please see www.edgehill.ac.uk/about/legal/privacy

#### What do are we doing about COVID-19 (coronavirus)?

We are continuing to do research as we think it is important. Edge Hill University and the NHS are taking the necessary precautions to ensure the safety of research participants, staff and students.
**If you have symptoms of coronavirus or are self-isolating, you must not come to hospital**. Call 111 or visit 111.nhs.uk/covid-19/ for further advice. Please wear the masks provided and sanitise hands on arrival. The coronavirus pandemic is rapidly evolving, please visit gov.uk/coronavirus for up-to-date guidance.

#### What is the study about?

In this study, we want to find out what may prevent people with diabetes from taking part in research. We would like to do this by having a chat and asking you some informal questions on the phone. Your participation could help improve diabetes research in the future.

#### What does this study involve?

You are being asked to take part in a recorded interview. The interview will be like a conversation. The interviewer will ask you about your views on some of the things that might have prevented you from taking part in earlier research. We would also like to ask you about your views on mental health in diabetes and about blood sugar targets in diabetes. The interview will take about 15 to 30 minutes. If you are happy to have an interview, please sign and return the enclosed consent form using the prepaid stamped, addressed envelope. We will organise a convenient time to call you.

#### Why have I been chosen?

You have been chosen because you are registered to come to a diabetes clinic in St Helens Hospital. To find out more information on being chosen for research please visit www.nhs.uk/your-nhs-data-matters or call 0300 303 5678.

#### Do I have to take part?

It is up to you. If you want to take part, you will be given this information sheet to keep. We will ask each participant to sign a consent form. You can withdraw your consent and stop taking part at any time.

#### I am happy to take part, what do I do next?

Please sign the consent form and put your initials in the boxes. Send it back to us using the enclosed pre-paid stamped, addressed envelope. The researcher will contact you to book a convenient time for the telephone interview.

#### What if I agree to take part and change my mind?

You can withdraw from the interview study at any time, without giving a reason. Your diabetes care will not be affected. If you withdraw, we will no longer collect any information about you or from you. We will keep information about you that we have already obtained as this information may have already been used in some analyses. If this is the case, we will use the minimum personally identifiable information.

#### What are the possible disadvantages of taking part?

There are no disadvantages to taking apart. All we will ask for is a little bit of your time for an interview.

#### What are the possible benefits of taking part?

The research is not meant to provide you with a direct benefit. We genuinely believe that ongoing research in diabetes is important. By taking part, you will add to our knowledge which may help patients in future.

#### Expenses and/or payments.

No expenses or payments will be available.

#### What if something goes wrong/who can I complain to?

If something goes wrong, you can email the interviewer. Alternatively, you can send us a letter (address below). If this does not achieve a suitable result, you should contact the Research Governance Officer. If you do this, please give the name of the study and the IRAS ID (at the top the page).

#### Will my participation in the study be confidential?

All data about you will be kept securely on NHS premises. Any information you give will be anonymised before publication. Your data will be given a code which will be used instead of your name. Any identifiable information you may give will be anonymised when collected and nothing identifiable will be used in publications. We will let your GP know that you are taking part. In some cases, if something important related to your health comes up, we might have to let your GP know. To continue supporting future research, anonymised data will be made available for other research groups to use. Where appropriate and necessary, regulatory inspectors or NHS research offices will be allowed access to your personal data to verify the study results.

#### What will happen to the results of this study?

The results of the study may be printed in medical journals, reports and presentations.

#### Who is organising and funding the research? Who has reviewed this study?

This research is funded by the Diabetes Department, St Helens Hospital. Research is supervised by Edge Hill University. This study has been approved by University Research Ethics Committee and the Health Research Authority.

Address:
ATTAINS Study, Diabetes Centre, St Helens Hospital, Marshalls Cross Road, St Helens, WA9 3DA

**Thank you** for volunteering to take part in this study

For further information, contact: Dr Sam Westall

NHS Trust

**CONSENT FORM – Study A**

**(Randomised Feasibility Study)**

Study ID:

Document version 1.0.0 – 21/9/20

Title of Project:

### HbA1c TargeT AchIevemeNt in diabeteS study

### (ATTAINS study) – Study A

IRAS ID: **291254**

*(Form to be on headed paper)*

Name of Researcher: Dr Sam Westall

Chief Investigator: Dr Niall Furlong

Please initial each box

1. I confirm that I have read Study A participant information sheet dated 26/02/21 (v 1.0.3) for the above study. I have had the opportunity to consider the information, ask questions and have had these answered satisfactorily. [1]
2. I understand that my participation is voluntary and that I am free to withdraw at any time without giving any reason, without my medical care or legal rights being affected. [2]
3. I understand that relevant sections of my medical notes and data collected during the study, may be looked at by individuals from St Helens and Knowsley Teaching Hospitals NHS Trust or from regulatory authorities where it is relevant to my taking part in this research. I give permission for these individuals to have access to my records. [3]
4. I understand that the information collected about me may be used to support other research in the future and may be shared anonymously with other researchers. [4]
5. I agree to my General Practitioner being informed of my participation in the study and I agree to my General Practitioner being informed if something important related to my health comes up in the study, including any necessary exchange of information about me between my GP and the research team. [5]
6. I understand that the information held and maintained by St Helens and Knowsley Teaching Hospitals NHS Trust may be used to help contact me or provide information about my health status. [6]
7. I agree to take part in the above study. [7]

| Name of Participant | Date | Signature |
|---|---|---|
| Name of Person taking consent | Date | Signature |

When completed: 1 for participant; 1 for researcher site file; 1 to be kept in medical notes.
Document ref: ATTAINS Study - Study A - Consent form - v 1.0.0 - 2020-09-21

**CONSENT FORM – Study B**

**Audio recording/Interview** Study ID:

Document version 1.0.0 – 21/9/20
Title of Project:

### HbA1c TargeT AchIevemeNt in diabeteS study (ATTAINS study) – Study B

IRAS ID: **291254** *(Form to be on headed paper)*

Name of Researcher: Dr Sam Westall Chief Investigator: Dr Niall Furlong

Please initial each box

1. I confirm that I have read Study B participant information sheet dated 23/10/20 (v 1.0.1) for the above study. I have had the opportunity to consider the information, ask questions and have had these answered satisfactorily. 1
2. I understand my interview will be recorded, transcribed and anonymised and that extracts from my interview may be quoted anonymously in print and online when the findings of the study are published. 2
3. I understand that the recordings will be stored securely until completion of the research, at which point they will be erased. 3
4. I understand that relevant sections of my medical notes and data collected during the study, may be looked at by individuals from St Helens and Knowsley Teaching Hospitals NHS Trust or from regulatory authorities where it is relevant to my taking part in this research. I give permission for these individuals to have access to my records. 4
5. I understand that my participation is voluntary and that I am free to withdraw at any time without giving any reason, without my medical care or legal rights being affected. In addition, should I choose not to answer any particular question, or questions, I am free to decline. 5
6. I understand that the information collected about me may be used to support other research in the future and may be shared anonymously with other researchers. 6
7. I agree to my General Practitioner being informed of my participation in the study and I agree to my General Practitioner being informed if something important related to my health comes up in the study, including any necessary exchange of information about me between my GP and the research team. 7
8. I understand that the information collected about me will be used to support other research in the future and may be shared anonymously with other researchers. 8
9. I agree to take part in the above study. 9

| | | |
|---|---|---|
| Name of Participant | Date | Signature |
| Name of Person taking consent | Date | Signature |

When completed: 1 for participant; 1 for researcher site file; 1 to be kept in medical notes.
Document ref: ATTAINS Study - Study B - Consent form - v 1.0.0 - 2020-09-21

**CONSENT FORM – Study C**

**Audio recording/Interview**

Study ID:

Document version 1.0.0 – 28/9/20

Title of Project:

### HbA1c TargeT AchIevemeNt in diabeteS study (ATTAINS study) – Study C

IRAS ID: **291254**

*(Form to be on headed paper)*

Name of Researcher: Dr Sam Westall

Chief Investigator: Dr Niall Furlong

Please initial each box

1. I confirm that I have read Study C participant information sheet dated 23/10/20 (v 1.0.1) for the above study. I have had the opportunity to consider the information, ask questions and have had these answered satisfactorily. [1]
2. I understand my interview will be recorded, transcribed and anonymised and that extracts from my interview may be quoted anonymously in print and online when the findings of the study are published. [2]
3. I understand that the recordings will be stored securely until completion of the research, at which point they will be erased. [3]
4. I understand that relevant sections of my medical notes and data collected during the study, may be looked at by individuals from St Helens and Knowsley Teaching Hospitals NHS Trust or from regulatory authorities where it is relevant to my taking part in this research. I give permission for these individuals to have access to my records. [4]
5. I understand that my participation is voluntary and that I am free to withdraw at any time without giving any reason, without my medical care or legal rights being affected. In addition, should I choose not to answer any particular question, or questions, I am free to decline. [5]
6. I understand that the information collected about me may be used to support other research in the future and may be shared anonymously with other researchers. [6]
7. I agree to my General Practitioner being informed of my participation in the study and I agree to my General Practitioner being informed if something important related to my health comes up in the study, including any necessary exchange of information about me between my GP and the research team. [7]
8. I understand that the information collected about me will be used to support other research in the future and may be shared anonymously with other researchers. [8]
9. I agree to take part in the above study. [9]

| | | |
|---|---|---|
| Name of Participant | Date | Signature |
| Name of Person taking consent | Date | Signature |

When completed: 1 for participant; 1 for researcher site file.
Document ref: ATTAINS Study - Study C - Consent form - v1.0.0 - 2020-09-28

**CONSENT FORM – Study D**

**Audio recording/Interview**

Study ID:

Document version 1.0.0 – 21/12/20

Title of Project:

### HbA1c TargeT AchIevemeNt in diabeteS study (ATTAINS study) – Study D

IRAS ID: **291254**

*(Form to be on headed paper)*

Name of Researcher: Dr Sam Westall

Chief Investigator: Dr Niall Furlong

Please initial each box

1. I confirm that I have read Study D information sheet dated 21/12/20 (v 1.0.0) for the above study. I have had the opportunity to consider the information, ask questions and have had these answered satisfactorily.
2. I understand my interview will be recorded, transcribed and anonymised and that extracts from my interview may be quoted anonymously in print and online when the findings of the study are published.
3. I understand that the recordings will be stored securely until completion of the research, at which point they will be erased.
4. I understand that relevant sections of my medical notes and data collected during the study, may be looked at by individuals from St Helens and Knowsley Teaching Hospitals NHS Trust or from regulatory authorities where it is relevant to my taking part in this research. I give permission for these individuals to have access to my records.
5. I understand that my participation is voluntary and that I am free to withdraw at any time without giving any reason, without my medical care or legal rights being affected. In addition, should I choose not to answer any particular question, or questions, I am free to decline.
6. I understand that the information collected about me may be used to support other research in the future and may be shared anonymously with other researchers.
7. I agree to my General Practitioner being informed of my participation in the study and I agree to my General Practitioner being informed if something important related to my health comes up in the study, including any necessary exchange of information about me between my GP and the research team.
8. I understand that the information collected about me will be used to support other research in the future and may be shared anonymously with other researchers.
9. I agree to take part in the above study.

| | | |
|---|---|---|
| Name of Participant | Date | Signature |
| Name of Person taking consent | Date | Signature |

When completed: 1 for participant; 1 for researcher site file; 1 to be kept in medical notes.
Document ref: ATTAINS Study - Study D - Consent form - v 1.0.0 - 2020-12-21

ATTAINS Study. **IRAS ID: 291254** Study ID:

### Diabetes and HbA1c

In people with diabetes, getting the right level of blood sugar is important for good physical health. Doctors will often give you a HbA1c goal to help you achieve this. As part of the research you are in, we are studying how HbA1c goals affect your well-being.

#### What is HbA1c?

It is a test that tells you your average blood sugar over the past few months. Sugar is needed for energy. In diabetes, your body can't use sugar as well. Sometimes there is too much sugar left in your blood. HbA1c measures how much sugar is stuck in your red blood cells.

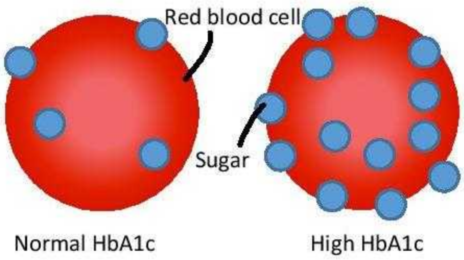

#### Why does it matter?

Everyone's diabetes is different. Generally, doctors will want to lower your HbA1c if it is too high in order to lower your risk of heart disease and eye, kidney and nerve problems. Sometimes, doctors or patients want to relax their HbA1c goal based on personal experiences, or when it is not beneficial.

#### How can you achieve your HbA1c goal?

Lots of things will affect your HbA1c. This includes your mood, stress, sickness, changes in lifestyle and medications you take. There are many things you can do to change your HbA1c but these will depend on whether you have Type 1 or Type 2 diabetes and your overall health.

In your clinic appointments at St Helens Hospital Diabetes Centre, your doctor will discuss the best way to support you to manage your diabetes.

#### What about the study?

The researcher will discuss setting a HbA1c goal for you during the study. This is so we can see what effect HbA1c goals have on your well-being.

#### Your personal HbA1c study goal

Your HbA1c today: ☒ Your personal HbA1c goal: ◯

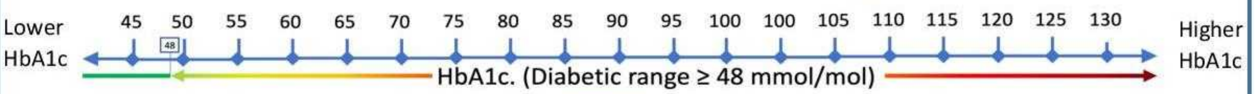

You are: Doing better than your goal: ☐

Doing worse than your goal: ☐

Your next review is: ___/___/______

#### HbA1c Lay summary script

As per the research protocol:
For study A, research visit 2, the patient should have the following completed:

- HbA1c, BMI, blood pressure.
- psychometric questionnaires.

The patient will then attend a face-to-face meeting with the researcher for the intervention (explicit HbA1c target setting).

The following summary of HbA1c will be read and discussed with the patient:

*"In people with diabetes, getting the right level of blood sugar is important for good physical health. Your diabetes doctor will often give you a HbA1c goal to help you achieve this. As part of the research you are in, we are studying how HbA1c goals affect your mental health.*

***So what is HbA1c?*** *Well, It is a test that tells you your average blood sugar over the past few months. Sugar is needed for energy. In diabetes, your body can't use sugar as well. Sometimes there is too much sugar left in your blood. HbA1c measures how much sugar is stuck in your red blood cells.*

***Why does HbA1c matter?*** *Everyone's diabetes is different. Generally, doctors will want to lower your HbA1c if it is too high in order to lower your risk of heart disease and eye, kidney and nerve problems. Sometimes, doctors or patients want to relax their HbA1c goal based on personal experiences, or when it is not beneficial.*

***How can you achieve your HbA1c goal?*** *Lots of things will affect your HbA1c. This includes your mood, stress, sickness, changes in lifestyle and medications you take. There are many things you can do to change your HbA1c but these will depend on whether you have Type 1 or Type 2 diabetes and your overall health.*
*In your clinic appointments at St Helens Hospital Diabetes Centre, your doctor will discuss the best way to support you to manage your diabetes.*

*In this study, I am going to give you a HbA1c goal and show you where your HbA1c currently is. This leaflet will also show you this."*

Researcher will then provide patient with the leaflet (ATTAINS Study - Study A - HbA1c Leaflet - v1.0.1 - 2020-12-07.docx) and address any questions the participant may have. The researcher will outline the participant's HbA1c goal versus their current HbA1c and inform the participant whether they are over or under-achieving (depending on whether the participant is in group A or B).

Once completed, thank the patient for their time. Inform the patient of their next appointment date in three months (if available). Visit 2 is completed.

### HbA1c TargeT AchIevemeNt in diabeteS study

### (ATTAINS study)

Department of Diabetes and Endocrinology
St Helens Hospital
Marshalls Cross Road
ST HELENS
Merseyside
WA9 3DA

Website: www.sthk.nhs.uk

Date:

IRAS ID: **291254**
Ref:
Hospital No:
NHS No:

Dear Dr ________,

Re: ______________ - __/__/__
____________________________________

Your patient has consented to be enrolled in research at St Helens and Knowsley Teaching Hospitals NHS Trust on DD/MM/YYYY.

In the ATTAINS Study, we are evaluating psychometric outcomes when using explicit HbA1c targets in people with diabetes over a 6-month period. The aims of the study are to test the feasibility of this type of project and to give preliminary outcome data. Some patients are also selected for interview during the study or, if they choose not to participate, to evaluate barriers to participation in research. This will not change the care that your patient will receive.

If you wish to know any further information about the research, please do not hesitate to get in touch via phone or email.

Yours Sincerely,

Dr Sam Westall
Clinical Research Fellow, St Helens and Knowsley Teaching Hospitals NHS Trust
PhD Research Student, Edge Hill University
E:
T: 01744 646668

Document ref: ATTAINS Study - GP info letter - v 1.0.0 - 2020-09-23

### HbA1c TargeT AchIevemeNt in diabeteS study

### (ATTAINS study)

Department of Diabetes and Endocrinology
St Helens Hospital
Marshalls Cross Road
ST HELENS
Merseyside
WA9 3DA

Website: www.sthk.nhs.uk

Date:

IRAS ID: **291254**
Ref:
Hospital No:
NHS No:

Dear Dr ________,

Re: ______________ - __/__/__
_____________________________________

Your patient is enrolled in the ATTAINS study at St Helens and Knowsley Teaching Hospitals NHS Trust. We are evaluating psychometric outcomes when using explicit HbA1c targets in people with diabetes. The aims of the study are to test the feasibility of this type of project and to give preliminary outcome data.

During research, we occasionally uncover health-related findings. When we reviewed the above patient, we noted _____________. This finding was identified during research, rather than during specific medical investigation. You may wish to consider further analysis or tests to assess the significance of this. Please ignore this letter if you are already aware. We have written to the patient separately to inform them of the finding.

If you wish to know any further information about the reasoning behind this letter, or about the research, please do not hesitate to get in touch via phone or email.

Yours Sincerely,

Dr Sam Westall
Clinical Research Fellow, St Helens and Knowsley Teaching Hospitals NHS Trust
PhD Research Student, Edge Hill University
E:
T: 01744 646668

Document ref: ATTAINS Study - GP update letter - v 1.0.0 - 2020-09-21

### HbA1c TargeT AchIevemeNt in diabeteS study

### (ATTAINS study)

Department of Diabetes and Endocrinology
St Helens Hospital
Marshalls Cross Road
ST HELENS
Merseyside
WA9 3DA
Website: www.sthk.nhs.uk

Date:

IRAS ID: **291254**
Ref: __________
Hospital No: ___________
NHS No: _______________
Patient ID: ______________ - __/__/__
_____________________________________

Dear Mr/Mrs/Miss _________,

**The ATTAINS Study**

We are a team of doctors and researchers from the Edge Hill University and from St Helens Hospital. We are interested in the health and needs of people with diabetes. We would like to invite you to take part in some research.

We want to study how giving you specific blood sugar goals effects your wellbeing. During the study, we want to look at your wellbeing and your blood sugar control. We may also ask you for an interview. The main aim of the study is to see if it is justifiable to do a bigger study like this in the future.

If you take part in this study, we will arrange to meet you when you come to hospital for your diabetes clinic. We have enclosed an information sheet for further details on the study.

We will call you soon to ask if you are interested and answer any questions you may have.

Thank you.

Yours Sincerely,

Dr Sam Westall
Clinical Research Fellow at St Helens and Knowsley Teaching Hospitals NHS Trust
PhD Research Student at Edge Hill University
E:
T: 01744 646668

Document ref: ATTAINS Study - Invitation letter (studies A and B) – v1.0.1 – 2020-10-30
*Enclosed: ATTAINS Study - Study A - Participant Information Sheet - v 1.0.3 - 2021-02-26*

### HbA1c TargeT AchIevemeNt in diabeteS study

### (ATTAINS study)

Department of Diabetes and Endocrinology
St Helens Hospital
Marshalls Cross Road
ST HELENS
Merseyside
WA9 3DA
Website: www.sthk.nhs.uk

Date:

IRAS ID: **291254**
Ref: __________
Hospital No: ___________
NHS No: _______________
Patient ID: ______________ - __/__/__
_____________________________________

Dear Mr/Mrs/Miss _________,

**The ATTAINS Study: barriers to participation in research interview**

We are a team of doctors and researchers from the Edge Hill University and from St Helens Hospital. We recently asked you if you would like to take part in research. We are interested in looking at things that prevent people from taking part in research so that we can make it better in the future.

We want to have a short interview with you over the telephone to ask your opinions about research and things that might prevent you from participating.

We have enclosed an information sheet for further details on the study. If you are happy to take part, please complete the consent form and send it back to us using the enclosed stamped, addressed envelope.

We will call you soon to ask if you are interested, answer any questions you may have and arrange a convenient time to have the telephone interview.

Thank you.

Yours Sincerely,

Dr Sam Westall
Clinical Research Fellow at St Helens and Knowsley Teaching Hospitals NHS Trust
PhD Research Student at Edge Hill University
E:
T: 01744 646668

Document ref: ATTAINS Study - Invitation letter (study D) – v1.0.0 – 2021-01-20
*Enclosed: (1) ATTAINS Study - Study D - Participant Information Sheet - v 1.0.0 - 2020-12-21;*
*(2) ATTAINS Study – Study D – Consent form – v 1.0.0 – 2020-12-21*
*(3) Stamped, addressed return envelope*

### HbA1c TargeT AchIevemeNt in diabeteS study

### (ATTAINS study)

Department of Diabetes and Endocrinology
St Helens Hospital
Marshalls Cross Road
ST HELENS
Merseyside
WA9 3DA
Website: www.sthk.nhs.uk
Date:

IRAS ID: **291254**
Ref: __________
Hospital No: ___________
NHS No: _______________

Patient ID: ______________ - __/__/__
______________________________________

Dear Mr/Mrs/Miss _________,

You are currently enrolled in the ATTAINS study at St Helens Hospital.

As part of the research, you have kindly attended a research visit. During your most recent visit, we noted something that may benefit from a review with your GP. *(explain in more detail if required here)*.

It may be that your GP already knows about this, in which case there is nothing that needs doing. We have written to your GP separately to check this. Your GP may get in touch with you to discuss the matter further.

If you have any questions about this matter, or the research, please get in touch with me via phone or email.

Yours Sincerely,

Dr Sam Westall
Clinical Research Fellow at St Helens and Knowsley Teaching Hospitals NHS Trust
PhD Research Student at Edge Hill University

E:
T: 01744 646668

Document ref: ATTAINS Study - Patient update letter- v1.0.0 - 2020-09-21

### HbA1c TargeT AchIevemeNt in diabeteS study

### (ATTAINS study)

**Subject line:** Invite to an interview as part of local research in diabetes (IRAS ID: **291254**)

**Email content:**

Dear Doctor/Dietician/DSN,

We would like to invite you to an interview which will form part of the a piece of research. The ATTAINS study is a feasibility study. The study will look at various aspects of using individualised glycaemic targets in people with diabetes. As healthcare professionals, your opinions on this process are of interest to the study.

This research is being undertaken as part of a PhD at Edge Hill University.

Briefly, this interview will involve healthcare professionals in secondary care who are directly involved in the care of people with diabetes. We would like to explore your experience and opinions on the use of glycaemic targets and on the use of individualised treatment targets in people with diabetes.

Please read the attached participant information sheet. If you would like to take part or have any further questions, please reply to this email and we will arrange a convenient day and time for the interview.

Yours Sincerely,

Dr Sam Westall
Clinical Research Fellow, St Helens and Knowsley Teaching Hospitals NHS Trust
PhD Research Student, Edge Hill University

E:
T: 01744 646668

Research Team:

| | |
|---|---|
| Dr Simon Watmough | Director of Studies |
| Dr Greg Irving | Supervisor |
| Dr Prakash Narayanan | Supervisor |
| Dr Niall Furlong | Chief Investigator |

Attached: ATTAINS Study - Study C - Healthcare interview PIS - v1.0.1 - 2020-10-23.pdf

Document ref: ATTAINS Study - Study C - Healthcare interview email invite - v1.0.0 - 2020-09-28

### HbA1c TargeT AchIevemeNt in diabeteS study

### (ATTAINS study)

**Subject line:** Invite to an interview as part of local research in diabetes (IRAS ID: **291254**)

**Email content:**

Dear Doctor/Dietician/DSN,

We recently emailed you to ask if you would like to be involved in a piece of research. We would like to have a brief interview with you. This interview will form part of a study evaluating the use of individualised glycaemic targets in people with diabetes.

The interview should only take a short amount of time.

Please reply to this email to let us know if you would or would not like to take part. If you would like to take part, we will arrange a convenient day and time for the interview with you.

We have attached an information sheet to answer any questions you may have.

Yours Sincerely,

Dr Sam Westall
Clinical Research Fellow, St Helens and Knowsley Teaching Hospitals NHS Trust
PhD Research Student, Edge Hill University

E:
T: 01744 646668

Research Team:

| | |
|---|---|
| Dr Simon Watmough | Director of Studies |
| Dr Greg Irving | Supervisor |
| Dr Prakash Narayanan | Supervisor |
| Dr Niall Furlong | Chief Investigator |

Attached: ATTAINS Study - Study C - Healthcare interview PIS - v1.0.1 - 2020-10-23.pdf

**Acceptability survey – Study A.** IRAS ID: **291254**

Document version 1.0.0 – 06-10-2020 Study ID:

Title of Project:

### HbA1c TargeT AchIevemeNt in diabeteS study

### (ATTAINS study) – Study A

IRAS ID: *(Form to be on headed paper)*

Name of Researcher: Dr Sam Westall

Chief Investigator: Dr Niall Furlong

You have been involved in a study looking at the treatment goals in diabetes. Now that the study has finished, we would like to ask your views on it. Please read the following statements and circle your answer.

To what extent do you agree or disagree with these statements about the research:

| **1) The recruitment process was easy.** | | | | |
|---|---|---|---|---|
| Strongly Agree | Agree | Neutral | Disagree | Strongly Disagree |
| Please explain: | | | | |

| **2) Random selection into different groups in the study was acceptable.** | | | | |
|---|---|---|---|---|
| Strongly Agree | Agree | Neutral | Disagree | Strongly Disagree |
| Please explain: | | | | |

**Acceptability survey – Study A.** IRAS ID: **291254**

Document version 1.0.0 – 06-10-2020 Study ID:

| **3) I was given enough information about the study.** | | | | |
|---|---|---|---|---|
| Strongly Agree | Agree | Neutral | Disagree | Strongly Disagree |
| Please explain: | | | | |

| **4) It was acceptable to be given a blood sugar target.** | | | | |
|---|---|---|---|---|
| Strongly Agree | Agree | Neutral | Disagree | Strongly Disagree |
| Please explain: | | | | |

| **5) After the study, I have a better understanding of blood sugar treatment goals in diabetes.** | | | | |
|---|---|---|---|---|
| Strongly Agree | Agree | Neutral | Disagree | Strongly Disagree |
| Please explain: | | | | |

**Acceptability survey – Study A.** IRAS ID: **291254**

Document version 1.0.0 – 06-10-2020 Study ID:

| **6) The time commitments of the study were acceptable.** | | | | |
|---|---|---|---|---|
| Strongly Agree | Agree | Neutral | Disagree | Strongly Disagree |
| Please explain: | | | | |

| **7) The questionnaires were easy to understand.** | | | | |
|---|---|---|---|---|
| Strongly Agree | Agree | Neutral | Disagree | Strongly Disagree |
| Please explain: | | | | |

| **8) If I had any problems, it was easy to contact the researchers.** | | | | |
|---|---|---|---|---|
| Strongly Agree | Agree | Neutral | Disagree | Strongly Disagree |
| Please explain: | | | | |

Page 3 of 4

Document ref: ATTAINS Study - Study A - Acceptability survey - v1.0.0 - 2020-10-06

**Acceptability survey – Study A.** IRAS ID: **291254**

Document version 1.0.0 – 06-10-2020 Study ID:

| **As a result of your visit to the doctor today do you feel you are** | **Much better** | **Better** | **Same or less** | **Not applicable** |
|---|---|---|---|---|
| able to cope with life? | ☐ | ☐ | ☐ | ☐ |
| able to understand your illness? | ☐ | ☐ | ☐ | ☐ |
| able to cope with your illness? | ☐ | ☐ | ☐ | ☐ |
| able to keep yourself healthy? | ☐ | ☐ | ☐ | ☐ |
| | **Much more** | **More** | **Same or less** | **Not applicable** |
| confident about your health? | ☐ | ☐ | ☐ | ☐ |
| able to help yourself? | ☐ | ☐ | ☐ | ☐ |

| **Further comments:** |
|---|

**Thank you for taking the time to complete this survey.** The research team really appreciates your involvement in this study. If you have any questions, please ask the researcher.

**B to P survey – Study A.** IRAS ID: **291254**

Document version 1.0.0 – 13-01-2021 Study ID:

Title of Project:

### HbA1c TargeT AchIevemeNt in diabeteS study (ATTAINS study) – Barriers to Participation

IRAS ID: **291254**

Name of Researcher: Dr Sam Westall

*(Form to be on headed paper)*

Chief Investigator: Dr Niall Furlong

We have been asking people to take part in diabetes research. People often have many good reasons to decline taking part. We would like to ask you a few questions to see what we could do in the future to help more people take part in research.

To what extent do you agree or disagree with these statements about taking part in research:

| **1) My work commitments stopped me from taking part in research.** | | | | |
|---|---|---|---|---|
| Strongly Agree | Agree | Neutral | Disagree | Strongly Disagree |
| Comments: | | | | |

| **2) I am a person with a disability, which makes it difficult to take part in research.** | | | | |
|---|---|---|---|---|
| Strongly Agree | Agree | Neutral | Disagree | Strongly Disagree |
| Comments: | | | | |

| **3) I did not understand what the research is about.** | | | | |
|---|---|---|---|---|
| Strongly Agree | Agree | Neutral | Disagree | Strongly Disagree |
| Comments: | | | | |

| **4) - A language barrier prevented me from taking part in the research.<br>- Bariera językowa uniemożliwiła mi udział w badaniach.<br>- Zúčastniť sa na výskume mi bránila jazyková bariéra.<br>- 语言障碍使我无法参与研究<br>- Isang hadlang sa wika ang pumigil sa akin na makilahok sa pagsasaliksik.<br>- ஒரு மொழித் தடை என்னை ஆராய்ச்சியில் பங்கேற்கவிடாமல் தடுத்தது.** | | | | |
|---|---|---|---|---|
| Strongly Agree | Agree | Neutral | Disagree | Strongly Disagree |
| Comments: | | | | |

| **5) I do not have enough time to take part in this research.** | | | | |
|---|---|---|---|---|
| Strongly Agree | Agree | Neutral | Disagree | Strongly Disagree |
| Comments: | | | | |

**B to P survey – Study A.** IRAS ID: **291254**

Document version 1.0.0 – 13-01-2021 Study ID:

| **6) I am a person with other physical or mental health problems that prevent me from taking part in research.** | | | | |
|---|---|---|---|---|
| Strongly Agree | Agree | Neutral | Disagree | Strongly Disagree |
| Comments: | | | | |

| **Further comments:** |
|---|

**<u>Thank you for taking the time to complete this survey.</u>** The research team really appreciates your feedback. If you have any questions, please ask the researcher.

### HbA1c TargeT AchIevemeNt in diabeteS study

### (ATTAINS study)

This page explains how health researchers use information from patients. If you are asked to take part in research, you can ask what will happen in the study.

**What is patient data?**

When you go to your GP or hospital, the doctors and others looking after you will record information about your health. This will include your health problems, and the tests and treatment you have had. They might want to know about family history, if you smoke or what work you do. All this information that is recorded about you is called patient data or patient information.

When information about your health care joins together with information that can show who you are (like your name or NHS number) it is called identifiable patient information. It's important to all of us that this identifiable patient information is kept confidential to the patient and the people who need to know relevant bits of that information to look after the patient. There are special rules to keep confidential patient information safe and secure.

**What sort of patient data does health and care research use?**

There are lots of different types of health and care research.

If you take part in a clinical trial, researchers will be testing a medicine or other treatment. Or you may take part in a research study where you have some health tests or answer some questions. When you have agreed to take part in the study, the research team may look at your medical history and ask you questions to see if you are suitable for the study. During the study you may have blood tests or other health checks, and you may complete questionnaires. The research team will record this data in special forms and combine it with the information from everyone else in the study. This recorded information is research data.

In other types of research, you won't need to do anything different, but the research team will be looking at some of your health records. This sort of research may use some data from your GP, hospital or central NHS records. Some research will combine these records with information from other places, like schools or social care. The information that the researcher collects from the health records is research data.

IRAS ID: **291254**

**Why does health and care research use information from patients?**

In clinical trials, the researchers are collecting data that will tell them whether one treatment is better or worse than other. The information they collect will show how safe a treatment is, or whether it is making a difference to your health. Different people can respond differently to a treatment. By collecting information from lots of people, researchers can use statistics to work out what effect a treatment is having.

Other types of research will collect data from lots of health records to look for patterns. It might be looking to see if any problems happen more in patients taking a medicine. Or to see if people who have screening tests are more likely to stay healthier.

Some research will use blood tests or samples along with information about the patient's health. Researchers may be looking at changes in cells or chemicals due to a disease.

All research should only use the patient data that it really needs to do the research. You can ask what parts of your health records will be looked at.

**How does research use patient data?**

If you take part in some types of research, like clinical trials, some of the research team will need to know your name and contact details so they can contact you about your research appointments, or to send you questionnaires. Researchers must always make sure that as few people as possible can see this sort of information that can show who you are.

In lots of research, most of the research team will not need to know your name. In these cases, someone will remove your name from the research data and replace it with a code number. This is called coded data, or the technical term is pseudonymised data. For example, your blood test might be labelled with your code number instead of your name. It can be matched up with the rest of the data relating to you by the code number.

In other research, only the doctor copying the data from your health records will know your name. They will replace your name with a code number. They will also make sure that any other information that could show who you are is removed. For example, instead of using your date of birth they will give the research team your age. When there is no information that could show who you are, this is called anonymous data.

**Where will my data go?**

Sometimes your own doctor or care team will be involved in doing a research study. Often, they will be part of a bigger research team. This may involve other hospitals, or universities or companies developing new treatments. Sometimes parts of the research team will be in other countries. You can ask about where your data will go. You can also check whether the data they get will include information that could show who you are. Research teams in other countries must stick to the rules that the UK uses.

All the computers storing patient data must meet special security arrangements.

IRAS ID: **291254**

**What are my choices about my patient data?**

- You can stop being part of a research study at any time, without giving a reason, but the research team will keep the research data about you that they already have. You can find out what would happen with your data before you agree to take part in a study.
- In some studies, once you have finished treatment the research team will continue to collect some information from your doctor or from central NHS records over a few months or years so the research team can track your health. If you do not want this to happen, you can say you want to stop any more information being collected.
- Researchers need to manage your records in specific ways for the research to be reliable. This means that they won't be able to let you see or change the data they hold about you. Research could go wrong if data is removed or changed.

**What happens to my research data after the study?**

Researchers must make sure they write the reports about the study in a way that no-one can work out that you took part in the study.

Once they have finished the study, the research team will keep the research data for several years, in case they need to check it. You can ask about who will keep it, whether it includes your name, and how long they will keep it.

Usually your hospital or GP where you are taking part in the study will keep a copy of the research data along with your name. The organisation running the research will usually only keep a coded copy of your research data, without your name included. This is kept so the results can be checked.

If you agree to take part in a research study, you may get the choice to give your research data from this study for future research. Sometimes this future research may use research data that has had your name and NHS number removed. Or it may use research data that could show who you are. You will be told what options there are. You will get details if your research data will be joined up with other information about you or your health, such as from your GP or social services.

Once your details like your name or NHS number have been removed, other researchers won't be able to contact you to ask you about future research.

Any information that could show who you are will be held safely with strict limits on who can access it.

You may also have the choice for the hospital or researchers to keep your contact details and some of your health information, so they can invite you to take part in future clinical trials or other studies. Your data will not be used to sell you anything. It will not be given to other organisations or companies except for research.

IRAS ID: **291254**

**Will the use of my data meet GDPR rules?**

GDPR stands for the General Data Protection Regulation. In the UK we follow the GDPR rules and have a law called the Data Protection Act. All research using patient data must follow UK laws and rules.

Universities, NHS organisations and companies may use patient data to do research to make health and care better.

When companies do research to develop new treatments, they need to be able to prove that they need to use patient data for the research, and that they need to do the research to develop new treatments. In legal terms this means that they have a 'legitimate interest' in using patient data.

Universities and the NHS are funded from taxes and they are expected to do research as part of their job. They still need to be able to prove that they need to use patient data for the research. In legal terms this means that they use patient data as part of 'a task in the public interest'.

If they could do the research without using patient data, they would not be allowed to get your data.

Researchers must show that their research takes account of the views of patients and ordinary members of the public. They must also show how they protect the privacy of the people who take part. An NHS research ethics committee checks this before the research starts.

**What if I don't want my patient data used for research?**

You will have a choice about taking part in a clinical trial testing a treatment. If you choose not to take part, that is fine.

In most cases you will also have a choice about your patient data being used for other types of research. There are two cases where this might not happen:

1. When the research is using anonymous information. Because it's anonymous, the research team don't know whose data it is and can't ask you.
2. When it would not be possible for the research team to ask everyone. This would usually be because of the number of people who would have to be contacted. Sometimes it will be because the research could be biased if some people chose not to agree. In this case a special NHS group will check that the reasons are valid. You can opt-out of your data being used for this sort of research. You can ask your GP about opting-out, or you can find out more at https://www.hra.nhs.uk/information-about-patients/

IRAS ID: **291254**

**Who can I contact if I have a complaint?**

If you want to complain about how researchers have handled your information, you should contact the research team. If you are not happy after that, you can contact the Data Protection Officer. The research team can give you details of the right Data Protection Officer.

Research team: Dr Sam Westall,

Chief Investigator: Dr Niall Furlong,

Research governance officer/data protection officer:

If you are not happy with their response or believe they are processing your data in a way that is not right or lawful, you can complain to the Information Commissioner's Office (ICO) (www.ico.org.uk or 0303 123 1113).

Patient interview topic guide and protocol (Study B)

**Venue**: Via telephone

**Please ensure a quiet and distraction-free environment for the interview. Study B interview dates and times should be arranged during patient visits to the diabetes centre during Study A final visit. Interviews should be carried after participants have completed study A.**

**Proposed duration:** 15 – 30 minutes.

**Structure:** Call participant and inform them that the call is recorded for research purposes and that their data will remain confidential. Briefly summarise the purpose of the study. Ensure participant has read and understood Study B participant information sheet. Answer any questions they may have. Consent already obtained during Study A.

**The interview will begin, (start audio recording):**
(Topics in blue, underlined. Themes identified in **bold.** Probing questions indented)

Greet participant, introductions.

"Please tell me, how long have you had diabetes?" (*easy question/ice-breaker*)
- "How were you diagnosed/what happened?"

#### Perceptions on the use of 'A1c' targets

"**What do you know about the goals/targets for your 'A1c'**?"
- "What do these mean personally for you?"
- "Why do you think 'A1c' is important?"

"**How did you feel about being given an 'A1c' target in the study**?"
- "Did it make you feel driven to do better?"
- "Did it make you disheartened and demotivated?"
- "If you were doing well, did it make you relax your efforts towards your diabetes?"
- "Did it make you feel elated?"
- "if you weren't doing well, did it make you more motivated or less motivated?"

**"Had you been given an 'A1c' target before the study?"**
- "How did it make you feel?"
- "Why did it make you feel like this?"

"**What is your opinion on using 'A1c' targets in people with diabetes?"**

#### Individualisation of 'A1c' targets

"There is a lot of research that tells us that **people with diabetes should be given individualised goals for 'A1c'**. What are your views about this?"
- "Can you tell me more about that?"
- "In your opinion, what are the positive things about having an 'A1c' target?"
- "In your opinion, what are the negative things about having and 'A1c' target?"

**Transcribe the interview as soon as possible after completion.**

Document ref: ATTAINS Study – Study B – patient interview guide – v 1.0.0 – 12-10-2020

Healthcare professionals interview topic guide and protocol
(Study C)

**Venue**: Via telephone/video call

**Ensure a distraction-free environment is located for telephone/video calls. Ensure the interview has appropriate access to telephone/video call facilities within the trust. Study C interview dates and times should be arranged via email with the healthcare professional.**

**Proposed duration:** 15 – 30 minutes.

**Structure:** Call participant and inform them that the call is recorded for research purposes and that their data will remain confidential. Briefly summarise the purpose of the study. Ensure participant has read and understood Study C participant information sheet. Answer any questions they may have. Ensure consent form has been completed.

**The interview will begin, (start audio recording):**
(Topics in blue, underlined. Themes identified in **bold.** Probing questions indented.)

Greet participant, introductions.

"Can you tell me a little bit about your professional background as a diabetes healthcare professional?" (easy question/ice-breaker)

#### Perceptions on the use of HbA1c targets

**"**HbA1c targets are universally used to aid management decisions in people with diabetes. We know it is beneficial to help patients achieve these targets. **What do you think patients' understanding of HbA1c is?"**
**"What sort of discussions would you normally have with a patient on the topic of HbA1c?"**
**"What do you think patients' views on HbA1c targets are?"**
> "What do you think could be done to help improve patient perceptions of HbA1c?"

**"What are your views on the use of HbA1c targets in people with diabetes?"**
**"What are the main advantages and disadvantages to using HbA1c targets in the management of people with diabetes?"**
**"If you knew that HbA1c targets had an impact on the mental health of people with diabetes, in what way would it change your discussions with patients?"**

#### Individualisation of HbA1c targets

A lot of research has suggested that HbA1c targets should be individualised to the patient.
**"How has this informed your practice?"**
**"What approach do you take to individualising HbA1c targets?"**
> "Have you found that this is beneficial?"
> "If so, in what way?"
> "If not, why not?"

**"In what way would you say HbA1c target-setting in people with diabetes could be improved?"**

**Transcribe the interview as soon as possible after completion.**

Healthcare professionals interview topic guide and protocol
(Study C)

Diabetes and mental health

Mental health comorbidities are known to have an impact upon the self-management abilities of people with diabetes. **"What do you think is the best way to improve the mental health of people with diabetes?"**
**"Do you think setting HbA1c targets in people with diabetes has an impact upon their mental health?"**
**"How can healthcare professionals be supported to deliver better care for people with diabetes?"**

**Probing:**
Active listening to guide further questioning
Appropriate use of silence
Politely interrupt and re-focus the interview if going off-topic
Clarify/elaborate/expand

**Closing:**
"Is there anything else you would like to add?"

Once complete, answer any questions the interviewee may have, stop the recording and thank the interviewee for their participation.

**Transcribe the interview as soon as possible after completion.**

Patient interview topic guide and protocol (Study D).
IRAS ID: **291254**

**Venue**: Via telephone

**Please ensure a quiet and distraction-free environment for the interview. Study D interview dates and times should be arranged by contacting the patient in advance.**

**Proposed duration:** 15 – 30 minutes.

**Structure:** Call participant and inform them that the call is recorded for research purposes and that their data will remain confidential. Briefly summarise the purpose of the study. Ensure participant has read and understood Study D participant information sheet. Answer any questions they may have. Consent form to be sent back in stamped, addressed envelope.

**The interview will begin, (start audio recording):**
(Topics in blue, underlined. Themes identified in **bold.** Probing questions indented)

Greet participant, introductions.

#### Icebreaker

"Please tell me, how long have you had diabetes?" (*easy question/ice-breaker*)

> "How were you diagnosed/what happened?"

#### Study information

"Did you feel you were given enough **information about the study**?"

> "Do you feel that you understood the information you were given?"
> "Is there anything you would have wanted to know about the study, which we perhaps did not tell you or explain to you in the letter or on the telephone?"
> "Did you know who the doctors were from whom you first received the letters about the study?"
> "Would it have made a difference to your decision if you knew them?"

#### Barriers to taking part

"May I ask the **reasons you declined** to take part in the study?"

> "What do you think would have **made a difference** to your decision to take part in the study?"
> "Is there anything that may have made you **more interested or willing** to take part in the study?"

#### Randomisation

"The study involved patients being **allocated by chance** to two groups."

> "Is there anything about that set up which influenced your decision not to take part?"

**Transcribe the interview as soon as possible after completion.**

Patient interview topic guide and protocol (Study D).
IRAS ID: **291254**

#### Travel

"You would have needed to come to St Helens Hospital for the research."

"Did this affect your decision to take part?"

#### Misc.

"Have you taken part in research studies before?"

"What are your feelings about your experience in previous research studies?"

#### Treatment targets

"As doctors, we often provide patients with goals to achieve to help improve their diabetes care."

"What is your **opinion about the use of treatment goals** in diabetes for your average blood sugar (A1c)?"

"Did this make any difference to your decision to take part?"

"**What do you know about the goals/targets for your 'A1c'**?"

"What do these mean personally for you?"

"Why do you think 'A1c' is important?"

**"What is your opinion on using 'A1c' targets in people with diabetes?"**

"There is a lot of research that tells us that **people with diabetes should be given individualised goals for 'A1c'**. What are your views about this?"

"Can you tell me more about that?"

"In your opinion, what are the positive things about having an 'A1c' target?"

"In your opinion, what are the negative things about having and 'A1c' target?"

#### Diabetes and mental health

"**Do you feel that diabetes has had an effect on your sense of well-being**?"

"In what way?"

"How have you dealt with this?"

"**Do you think that diabetes has affected your mental health**?"

"how/in what way?"

"why do you think that diabetes has had this effect?"

**"What do you think would make you feel better about living with diabetes?"**

**Probing:**

Active listening to guide further questioning
Appropriate use of silence
Politely interrupt and re-focus the interview if going off-topic
Clarify/elaborate/expand

**Closing:**

"Is there anything else you would like to add?"

Once complete, answer any questions the interviewee may have, stop the recording and thank the interviewee for their participation.

**Transcribe the interview as soon as possible after completion.**

### HbA1c TargeT AchIevemeNt in diabeteS study (ATTAINS study)

*A Data Management Plan created using DMPonline*

**Creator:** Sam Westall

**Affiliation:** Edge Hill University

**Funder:** St Helens and Knowsley Teaching Hospitals NHS Trust

**Template:** DCC Template

**ORCID iD:** https://orcid.org/0000-0003-1428-3681

**Project abstract:**

Background: Individualised treatment targets are commonly used in secondary care diabetes services in the UK in order to personalise the care of people with diabetes (PWD). Still, achievement of blood glucose targets in PWD is often poor. Generic and diabetes-specific mental health issues are a known driver of poor blood glucose target achievement and increased morbidity and mortality. Little is known, however, of the reciprocal effect that blood glucose target-setting has on the psychology and emotional health of PWD. Aim: In a series of related studies, the feasibility of evaluating psychometric outcomes when setting explicit blood glucose targets in PWD will be tested. Methods: Using a mixed-methods approach, a single-centre, randomised feasibility study will be conducted, aiming to recruit 50 eligible PWD due to attend a secondary care diabetes clinic in the Northwest UK. Participants will be randomised into one of two groups. Group A will have explicit average blood glucose (HbA1c) targets set 5mmol/mol above their present HbA1c reading. Group B will have explicit HbA1c targets set 5mmol/mol below their present HbA1c reading. Standard clinical care will continue. Measures of feasibility (eligibility rate, recruitment rate, response rate, retention rate, acceptability of study processes for participants and investigators) and preliminary outcome (psychometric questionnaire scores and HbA1c) will be evaluated at 0, 3 and 6 months. Semi-structured interviews with a purposive sample of study participants and healthcare professionals will capture experience and perspective on the study processes, evaluate barriers to participation in research and bring further meaning to the preliminary study findings. Interviews will be analysed thematically using the framework approach. Original contribution to knowledge: Whilst the current literature points to the clear physical health benefits of having individualised targets for diabetes, there is minimal understanding of the psychological effect of their use. The proposed research aims to test the feasibility of conducting a full-scale randomised control trial in evaluating psychometric outcomes when using explicit glycaemic targets in PWD.

**Last modified:** 20-01-2021

### HbA1c TargeT AchIevemeNt in diabeteS study (ATTAINS study)

#### Data Collection

##### What data will you collect or create?

Data will be collected in several formats using mixed methodologies as described in the research protocol. Data types and formats are illustrated below.

**Study A**:

*Data type and format*

Biochemical (finger-prick blood test: HbA1c in mmol/mol), physiological (blood pressure in mmHg, Body Mass Index in kg/m2) and baseline demographic (age, gender, ethnicity, diabetes type, diabetes duration, GP practice, postcode, educational status, prescriptions) data from a pragmatic sample of 100 randomly selected participants due to attend diabetes clinic at St Helens Hospital in the Northwest UK will be obtained at recruitment (baseline), 3 months and 6 months (endpoint).

Patient-reported outcome data (validated, self-completed, paper questionnaires) will be recorded from the same population at 3 months and 6 months. Biochemical, physiological and demographic data will be recorded in a password-protected Excel database (.xlsx) and analysed using SPSS (.sav). Questionnaire scores will be recorded in a password-protected excel database and analysed using SPSS.

*Data volume*

Electronic data: Estimated less than 100MB.

Physicial data: One filing cabinet drawer (physical copies of consent forms and completed questionnaires).

*Data sources*

Baseline data will be sourced from the STHK electronic patient record system.

**Study B**:

*Data type and format*

Audio-recorded semi-structured interview data will be saved on a password-protected dictaphone device in '.wav' or '.mp3' format.

Data from the interview will be transcribed and analysed for themes using the framework method of content analysis with support from NVIVO qualitative data analysis software (.nvb).

*Data volume*

Electronic data: Estimated 500MB.

Physical data: One filing cabinet drawer (physical copies of consent forms).

**Study C**:

*Data type and format*

Audio-recorded semi-structured interview data will be saved on a password-protected dictaphone device in '.wav' or '.mp3' format.

Data from the interview will be transcribed and analysed for themes using the framework method of content analysis with support from NVIVO qualitative data analysis software (.nvb).

*Data volume*

Electronic data: Estimated 500MB.

Physical data: One filing cabinet drawer (physical copies of consent forms).

**Study D**:

*Data type and format*

Audio-recorded semi-structured interview data will be saved on a password-protected dictaphone device in '.wav' or '.mp3' format.

Data from the interview will be transcribed and analysed for themes using the framework method of content analysis with support from NVIVO qualitative data analysis software (.nvb).

*Data volume*

Electronic data: Estimated 500MB.

Physical data: One filing cabinet drawer (physical copies of consent forms).

**All Studies:** Data storage usage for software applications and collected-data is anticipated to be less than 1-terabyte. The research will be written up in Microsoft Word (.docx) with referencing assistance from Mendeley (.ris). These software options have been chosen due to current wide usage in academia and Edge Hill University technical support options.

##### How will the data be collected or created?

The proposed studies will be undertaken using a mixed-methods approach.

*Methodologies*

The methodologies uses will be:

- a randomised feasibility study
- semi-structured interviews

*Files/data versioning*

Documents will be held in a hierarchical file system. Documents will be named in the following fashion:

'study title' - 'document name' - 'document version number (1.0.0)' - 'document creation/update date (YYYY-MM-DD)'.extension

Document version and date will be displayed in the file name and on each page of the document. Ratified master copies of the documents will be held as read-only copies in a master file on the computer of the principal investigator. Document version numbers are displayed as 'Major.Minor.Revision'. The first version of a document would be '1.0.0' with revisions displayed as '1.0.1', minor adaptations as '1.1.0' and major changes as '2.0.0'. Once updated documents have been ratified, old copies of the documents will be archived and the new copies will replace them in the master file. Document revisions will be recorded on a cover page within each document.

*Data collection*

A password-protected Microsoft Excel dataset template will be created for data collection purposes. The dataset master copy will be saved on an NHS-encrypted computer. Daily cloud backups are made to minimise data loss. Data will be obtained from the following sources:

1. Blood test POC HbA1c data will be captured from electronic patient records systems.
2. Baseline demographic data will be obtained firstly from electronic patient records systems and if unavailable, directly from the patient.
3. Physiological measurement (blood pressure, height, weight) will be obtained at participant visits.
4. Paper questionnaires will be completed by hand by study participants and subsequently inputted into the dataset by the study investigator.
5. Audio data on a password-protected electronic dictaphone, subsequently transcribed into a password-protected Microsoft Word document on an NHS-encrypted computer for analysis.

Quality control of all equipment will be as per NHS standards.

*Data security*

The secure holding of all data will be on password-protected NHS computers with NHS-level data encryption. Transfer of study data between computing devices will be either with an NHS-encrypted USB drive or a secure N3 (NHS network encrypted) email. Non-electronic data will be held in a secure file in a locked office on NHS sites when not in use.

#### Documentation and Metadata

##### What documentation and metadata will accompany the data?

Data collected will be inputted into a database for use in future research should it be required. The following data will be collected in the database:

- Provenance metadata: when and where the data was collected and by whom.
- Rights and access metadata: information on rights and access usage rules.
- Structural metadata: data will be formatted in an easily-accessible manner.

A 'readme.txt' file will accompany datasets to include: titles, author names, keywords, a data summary (detailing how and why the data was collected), research funders and data time points. Information will be provided on the methodology used to collect the data, analytical and procedural information, units of measurement, assumptions made, the format and file type of the data and the software used to collect and process the data.

It is planned for the data to be held in the Edge Hill University research data repository upon completion of the thesis. Data will be anonymised. Data access should be limited to those involved in health, social care and psychology research. Requests for access to the data should be submitted to the chief investigator to be reviewed. Access to the metadata may be re-used in any medium without prior permission for not-for-profit purposes in line with Edge Hill University policy.

#### Ethics and Legal Compliance

##### How will you manage any ethical issues?

Signed, informed, written consent will be obtained for all participants in the research at the following stages:

- entry into the randomised feasibility study
- prior to an audio-recorded interview

Consent will be requested to allow for data to be preserved, shared and reused in future research. Access to the data for future research will be via the University research data repository and granted at the discretion of the chief investigator, dependent on the nature of the request. The metadata will be accessible without prior permission for not-for-profit purposes in line with Edge Hill University policy.

All data obtained will be anonymised for person- and place-identifiable information during transcription/database input by removal of direct identifiers. The secure holding of all data during the study period will be on password-protected NHS computers with NHS-level data encryption. Transfer of study data between computing devices will be either with an NHS-encrypted USB drive or a secure N3 (NHS network encrypted) email. Non-electronic data (e.g. consent forms, completed questionnaire) will be held in a secure file in a separate locked office on NHS sites when not in use. Audio-recording data (.mp3/.wav) will be stored securely on a password-protected recorder device in a separate locked office on NHS sites when not in use. Audio recordings will be held separately to signed consent form documents. All paper documentation will be held in a separate, locked file in a separate, locked room. Any detailed, identifiable free-text responses from surveys or semi-structured interviews will have their meaning generalised during transcription to prevent disclosure. Direct identifiers in all data obtained will be replaced with a code. Identifier code data will be located in a password-protected Microsoft Excel document on NHS-encrypted computers.

University and Health Research Authority ethical approval will be obtained prior to recruitment/commencement of data collection.

##### How will you manage copyright and Intellectual Property Rights (IPR) issues?

Following completion of the project, the data will be owned by Edge Hill University.

Data will be made available following the publication of the research results on completion of the PhD project.

#### Storage and Backup

##### How will the data be stored and backed up during the research?

Edge Hill University IT systems allocate students 1 terabyte (TB) of storage space for secure cloud storage of documents (via Microsoft OneDrive). This storage space will be used to back up study resource documents (participant information sheets, etc) and for back up of the thesis document. Key research data files containing any sensitive information (e.g. data collected) will be stored on an NHS-issued, encrypted, password-protected NHS laptop. Data will be stored via the laptop on NHS trust cloud storage associated with the PhD student's clinical research fellow post in the NHS. Therefore, if data were to be lost or become corrupt, for whatever reason, regular (daily) backups are accessible from the NHS trust network. The PhD student, Dr Sam Westall, will be responsible for maintaining data storage, backups and security.

Physical data (e.g. completed questionnaires, audio recording devices, consent forms) will be catalogued, filed securely and placed in a separate, locked room on NHS property within one day of receipt. Physical data will be accessed when required on hospital site. Physical data will remain on-site until no longer required, at which point it will be destroyed.

##### How will you manage access and security?

The main risks to consider are unauthorised access to computers, audio recording devices and paper documents via theft, loss of device or by other means.

Risks will be minimised by:

- ensuring all electronic devices are encrypted, password protected and locked when not in use.
- keeping all physical data in a secure, locked file after being collected and depositing into a locked room on NHS site when not in use, in a timely manner.
- ensuring investigators are kept up to date with information governance and data security NHS training

#### Selection and Preservation

##### Which data are of long-term value and should be retained, shared, and/or preserved?

All data collected could be of potential value for future research. As such, the collected, anonymised computer data will be submitted to the Edge Hill University research data repository for use.

Physical copies of data will be kept for the duration specified by St Helens and Knowsley Teaching Hospitals NHS Trust (STHK) Research Guidelines, at which point further retention of the data can be reviewed. Consent forms will be kept for as long as the research data are retained in the data repository. Following the completion of the research, they will be digitised and stored securely (encrypted) with the university. At this point, original physical copies will be destroyed securely (https://www.ukdataservice.ac.uk/manage-data/legal-ethical/consent-data-sharing/consent-forms).

##### What is the long-term preservation plan for the dataset?

Data will be held for as long as felt useful by Edge Hill University. Access to data and metadata from the university is free of charge. Submission to the repository is free of charge.

#### Data Sharing

##### How will you share the data?

Data will be shared in a number of ways:

1. The research will be published in medical journals and on posters at conferences in order to disseminate the results. Publications are planned between September 2021 and September 2022.
2. Results relevant to the participants will be made available to them, should they wish.
3. One of the questionnaire licence holders has requested results to be sent to them.
4. Data from the research will be presented to the participating GP practices and to the hospital.
5. Research student poster presentations are planned at the university.
6. Abstract submission with subsequent poster and/or oral presentation at diabetes conferences.
7. The data will be made available in the university research data repository (access to which is freely available via the university website) following completion of the thesis.

##### Are any restrictions on data sharing required?

Exclusive use of the data will be required until all publications related to the research have been submitted, after which the data will be submitted to the research data repository. In order to widen the research publication dissemination, 'green' open access status for published articles will be sought.

#### Responsibilities and Resources

##### Who will be responsible for data management?

Responsibility for the implementation, review and revision of the data management plan lies with the PhD student, Dr Sam Westall. Responsibility for each data management activity lies with Dr Sam Westall unless stated otherwise. This includes, but is not limited to: data capture, metadata production, data quality, data storage and backup, data archiving and data sharing.

##### What resources will you require to deliver your plan?

The provision of all the required hardware and software services and support are provided via the STHK IT services and Edge Hill University. Further technical expertise, such as advanced statistical calculations can be supported by expert staff within the NHS and Edge Hill University. A breakdown of the costs involved in the research is contained within the 'Study costs.xlsx' spreadsheet.

EQ-5D-5L

**Health Questionnaire**

**English version for the UK**

*UK (English) © 2009 EuroQol Group EQ-5D™ is a trade mark of the EuroQol Group*

Under each heading, please tick the ONE box that best describes your health TODAY.

**MOBILITY**

I have no problems in walking about ❑

I have slight problems in walking about ❑

I have moderate problems in walking about ❑

I have severe problems in walking about ❑

I am unable to walk about ❑

**SELF-CARE**

I have no problems washing or dressing myself ❑

I have slight problems washing or dressing myself ❑

I have moderate problems washing or dressing myself ❑

I have severe problems washing or dressing myself ❑

I am unable to wash or dress myself ❑

**USUAL ACTIVITIES** *(e.g. work, study, housework, family or leisure activities)*

I have no problems doing my usual activities ❑

I have slight problems doing my usual activities ❑

I have moderate problems doing my usual activities ❑

I have severe problems doing my usual activities ❑

I am unable to do my usual activities ❑

**PAIN / DISCOMFORT**

I have no pain or discomfort ❑

I have slight pain or discomfort ❑

I have moderate pain or discomfort ❑

I have severe pain or discomfort ❑

I have extreme pain or discomfort ❑

**ANXIETY / DEPRESSION**

I am not anxious or depressed ❑

I am slightly anxious or depressed ❑

I am moderately anxious or depressed ❑

I am severely anxious or depressed ❑

I am extremely anxious or depressed ❑

*UK (English) © 2009 EuroQol Group EQ-5D™ is a trade mark of the EuroQol Group*

- We would like to know how good or bad your health is TODAY.
- This scale is numbered from 0 to 100.
- 100 means the best health you can imagine.
  0 means the worst health you can imagine.
- Mark an X on the scale to indicate how your health is TODAY.
- Now, please write the number you marked on the scale in the box below.

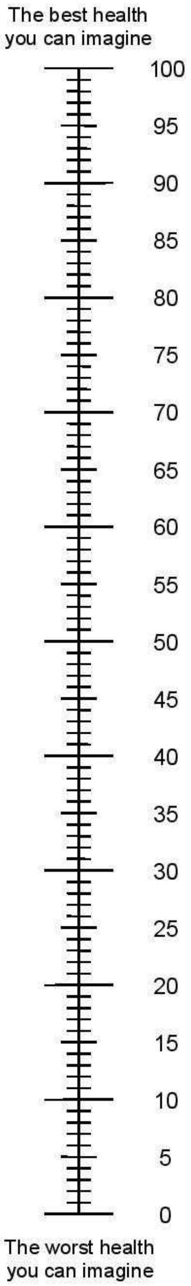

YOUR HEALTH TODAY = ☐

*UK (English) © 2009 EuroQol Group EQ-5D™ is a trade mark of the EuroQol Group*

Date: ___________________

**Date:** _______________

### University of Michigan Diabetes Research and Training Center

#### DIABETES ATTITUDE QUESTIONNAIRE

| PLEASE ANSWER THE FOLLOWING QUESTIONS |
| --- |

Date: ___________________

**Attitudes Toward Diabetes – DES**

| In general, I believe that I: | Strongly Agree | Agree | Neutral | Disagree | Strongly Disagree |
|---|---|---|---|---|---|
| 1. ...know what part(s) of taking care of my diabetes that I am **satisfied** with. | ( ) | ( ) | ( ) | ( ) | ( ) |
| 2. ...know what part(s) of taking care of my diabetes that I am **dissatisfied** with. | ( ) | ( ) | ( ) | ( ) | ( ) |
| 3. ...know what part(s) of taking care of my diabetes that I am ready to change. | ( ) | ( ) | ( ) | ( ) | ( ) |
| 4. ...know what part(s) of taking care of my diabetes that I am not ready to change. | ( ) | ( ) | ( ) | ( ) | ( ) |
| 5. ...can choose realistic diabetes goals. | ( ) | ( ) | ( ) | ( ) | ( ) |
| 6. ...know which of my diabetes goals are **most** important to me. | ( ) | ( ) | ( ) | ( ) | ( ) |
| 7. ...know the things about **myself** that either help or prevent me from reaching my diabetes goals. | ( ) | ( ) | ( ) | ( ) | ( ) |
| 8. ...can come up with good ideas to help me reach my goals. | ( ) | ( ) | ( ) | ( ) | ( ) |
| 9. ...am able to turn my diabetes goals into a workable plan. | ( ) | ( ) | ( ) | ( ) | ( ) |

University of Michigan
Diabetes Research and Training Center

Date: ____________________

| | Strongly Agree | Agree | Neutral | Disagree | Strongly Disagree |
|---|---|---|---|---|---|
| In general, I believe that I: | | | | | |
| 10. ...can reach my diabetes goals once I make up my mind. | ( ) | ( ) | ( ) | ( ) | ( ) |
| 11. ...know which **barriers** make reaching my diabetes goals more difficult. | ( ) | ( ) | ( ) | ( ) | ( ) |
| 12. ...can **think** of different ways to overcome barriers to my diabetes goals | ( ) | ( ) | ( ) | ( ) | ( ) |
| 13. ...can try out different ways of overcoming barriers to my diabetes goals. | ( ) | ( ) | ( ) | ( ) | ( ) |
| 14. ...am able to decide which way of overcoming barriers to my diabetes goals works best for me. | ( ) | ( ) | ( ) | ( ) | ( ) |
| 15. ...can tell how I'm feeling about **having** diabetes. | ( ) | ( ) | ( ) | ( ) | ( ) |
| 16. ...can tell how I'm feeling about **caring** for my diabetes | ( ) | ( ) | ( ) | ( ) | ( ) |
| 17. ...know the ways that having diabetes causes stress in my life. | ( ) | ( ) | ( ) | ( ) | ( ) |
| 18. ...know the **positive** ways I cope with diabetes-related stress. | ( ) | ( ) | ( ) | ( ) | ( ) |
| 19. ...know the **negative** ways I cope with diabetes-related stress. | ( ) | ( ) | ( ) | ( ) | ( ) |

University of Michigan
Diabetes Research and Training Center

Date: ____________________

| | Strongly Agree | Agree | Neutral | Disagree | Strongly Disagree |
|---|---|---|---|---|---|
| In general, I believe that I: | | | | | |
| 20. ...can cope well with diabetes-related stress. | ( ) | ( ) | ( ) | ( ) | ( ) |
| 21. ...know where I can get support for having and caring for my diabetes. | ( ) | ( ) | ( ) | ( ) | ( ) |
| 22. ...can ask for support for having and caring for my diabetes when I need it. | ( ) | ( ) | ( ) | ( ) | ( ) |
| 23. ...can support myself in dealing with my diabetes. | ( ) | ( ) | ( ) | ( ) | ( ) |
| 24. ...know what helps me stay motivated to care for my diabetes. | ( ) | ( ) | ( ) | ( ) | ( ) |
| 25. ..can motivate myself to care for my diabetes. | ( ) | ( ) | ( ) | ( ) | ( ) |
| 26. ...know enough about diabetes to make self-care choices that are right for me. | ( ) | ( ) | ( ) | ( ) | ( ) |
| 27. ...know enough about my-self as a person to make diabetes care choices that are right for me. | ( ) | ( ) | ( ) | ( ) | ( ) |
| 28. ...am able to figure out if it is worth my while to change how I take care of my diabetes. | ( ) | ( ) | ( ) | ( ) | ( ) |

**Thank you very much for completing this questionnaire.**

University of Michigan
Diabetes Research and Training Center

#### The Summary of Diabetes Self-Care Activities

The questions below ask you about your diabetes self-care activities during the past 7 days. If you were sick during the past 7 days, please think back to the last 7 days that you were not sick. Circle your answers.

##### Diet

How many of the last SEVEN DAYS have you followed a healthful eating plan?

0 1 2 3 4 5 6 7

On average, over the past month, how many DAYS PER WEEK have you followed your eating plan?

0 1 2 3 4 5 6 7

On how many of the last SEVEN DAYS did you eat five or more servings of fruits and vegetables?

0 1 2 3 4 5 6 7

On how many of the last SEVEN DAYS did you eat high fat foods such as red meat or full-fat dairy products?

0 1 2 3 4 5 6 7

##### Exercise

On how many of the last SEVEN DAYS did you participate in at least 30 minutes of physical activity? (Total minutes of continuous activity, including walking).

0 1 2 3 4 5 6 7

On how many of the last SEVEN DAYS did you participate in a specific exercise session (such as swimming, walking, biking) other than what you do around the house or as part of your work?

0 1 2 3 4 5 6 7

##### Blood Sugar Testing

On how many of the last SEVEN DAYS did you test your blood sugar?

0 1 2 3 4 5 6 7

On how many of the last SEVEN DAYS did you test your blood sugar the number of times recommended by your health care provider?

0 1 2 3 4 5 6 7

##### Foot Care

On how many of the last SEVEN DAYS did you check your feet?

0 1 2 3 4 5 6 7

On how many of the last SEVEN DAYS did you inspect the inside of your shoes?

0 1 2 3 4 5 6 7

##### Smoking

Have you smoked a cigarette—even one puff—during the past SEVEN DAYS?

YES NO

If yes, how many cigarettes did you smoke on an average day? Number of cigarettes:

___

Scoring for SDSCA

Step 1:

For items 1–10, use the number of days per week on a scale of 0–7. Note that this response scale will not allow for direct comparison with the percentages provided in Table 1.

Step 2: Scoring Scales

General Diet = Mean number of days for items 1 and 2.

Specific Diet = Mean number of days for items 3, and 4, reversing item 4 (0=7, 1=6, 2=5, 3=4, 4=3, 5=2, 6=1, 7=0).

Given the *low inter-item correlations for this scale*, using the individual items is recommended.

Exercise = Mean number of days for items 5 and 6.

Blood-Glucose Testing = Mean number of days for items 7 and 8.

Foot-Care = Mean number of days for items 9 and 10.

Smoking Status = Item 11 (0 = nonsmoker, 1 = smoker), and number of cigarettes smoked per day.

**Scoring for Additional Items**

Recommended regimen = Items 1A - 4A, and items 12A - 14A, no scoring required.

Diet = Use total number of days for item 5A.

Medications = Use item 6A - OR - 7A AND 8A, use total number of days for item 6A, use mean number of days if both 7A and 8A are applicable.

Foot-Care = Mean number of days for items 9A - 11A, after reversing 10A and including items 9 and 10 from the brief version.

#### Well-Being Questionnaire (W-BQ12)

**Please circle one number on each scale, from 3 (all the time) to 0 (not at all), to indicate how often you feel each statement has applied to you in the past few weeks.**

| | all the time | | | not at all |
|---|---|---|---|---|
| 1. I have crying spells or feel like it | 3 | 2 | 1 | 0 |
| 2. I feel downhearted and blue | 3 | 2 | 1 | 0 |
| 3. I feel afraid for no reason at all | 3 | 2 | 1 | 0 |
| 4. I get upset easily or feel panicky | 3 | 2 | 1 | 0 |
| 5. I feel energetic, active or vigorous | 3 | 2 | 1 | 0 |
| 6. I feel dull or sluggish | 3 | 2 | 1 | 0 |
| 7. I feel tired, worn out, used up or exhausted | 3 | 2 | 1 | 0 |
| 8. I have been waking up feeling fresh and rested | 3 | 2 | 1 | 0 |
| 9. I have been happy, satisfied or pleased with my personal life | 3 | 2 | 1 | 0 |
| 10. I have lived the kind of life I wanted to | 3 | 2 | 1 | 0 |
| 11. I have felt eager to tackle my daily tasks or make new decisions | 3 | 2 | 1 | 0 |
| 12. I have felt I could easily handle or cope with any serious problem or major change in my life | 3 | 2 | 1 | 0 |

**Please make sure that you have considered each of the 12 statements and have circled one number in response to each statement.**

DO NOT USE: contact

NOT FOR USE: for study administration only, by Dr S WESTALL, ref CB967
W-BQ12 © Prof Clare Bradley 6/96A. Standard UK English (instructions rev. 31.1.02)
Health Psychology Research Unit, Royal Holloway, University of London, Egham, Surrey, TW20 0EX, UK.

### Problem Areas in Diabetes Questionnaire (PAID)

**INSTRUCTIONS: Which of the following diabetes issues are currently a problem for you?**
Circle the number that gives the best answer for you. Please provide an answer for each question. Please bring the completed form with you to your next consultation where it will form the basis for a dialogue about how you are coping with your diabetes.

Patient name: Completion date: Interview date:

| | Not a problem | Minor problem | Moderate problem | Somewhat serious problem | Serious problem |
|---|---|---|---|---|---|
| 1. Not having clear and concrete goals for your diabetes care? | 0 | 1 | 2 | 3 | 4 |
| 2. Feeling discouraged with your diabetes treatment plan? | 0 | 1 | 2 | 3 | 4 |
| 3. Feeling scared when you think about living with diabetes? | 0 | 1 | 2 | 3 | 4 |
| 4. Uncomfortable social situations related to your diabetes care (e.g., people telling you what to eat)? | 0 | 1 | 2 | 3 | 4 |
| 5. Feelings of deprivation regarding food and meals? | 0 | 1 | 2 | 3 | 4 |
| 6. Feeling depressed when you think about living with diabetes? | 0 | 1 | 2 | 3 | 4 |
| 7. Not knowing if your mood or feelings are related to your diabetes? | 0 | 1 | 2 | 3 | 4 |
| 8. Feeling overwhelmed by your diabetes? | 0 | 1 | 2 | 3 | 4 |
| 9. Worrying about low blood sugar reactions? | 0 | 1 | 2 | 3 | 4 |
| 10. Feeling angry when you think about living with diabetes? | 0 | 1 | 2 | 3 | 4 |
| 11. Feeling constantly concerned about food and eating? | 0 | 1 | 2 | 3 | 4 |
| 12. Worrying about the future and the possibility of serious complications? | 0 | 1 | 2 | 3 | 4 |
| 13. Feelings of guilt or anxiety when you get off track with your diabetes management? | 0 | 1 | 2 | 3 | 4 |
| 14. Not "accepting" your diabetes? | 0 | 1 | 2 | 3 | 4 |
| 15. Feeling unsatisfied with your diabetes physician? | 0 | 1 | 2 | 3 | 4 |
| 16. Feeling that diabetes is taking up too much of your mental and physical energy every day? | 0 | 1 | 2 | 3 | 4 |
| 17. Feeling alone with your diabetes? | 0 | 1 | 2 | 3 | 4 |
| 18. Feeling that your friends and family are not supportive of your diabetes management efforts? | 0 | 1 | 2 | 3 | 4 |
| 19. Coping with complications of diabetes? | 0 | 1 | 2 | 3 | 4 |
| 20. Feeling "burned out" by the constant effort needed to manage diabetes? | 0 | 1 | 2 | 3 | 4 |

PAID - © 1999 Joslin Diabetes Center

#### 11.2 Appendix 2 – Schedule of Procedures

| **Procedures** | **Visits** | | | | |
|---|---|---|---|---|---|
| | **Pre-visit 1** | **Baseline/ screening: visit 1** | **Visit 2** | **Visit 3** | **Telephone interview** |
| | - | 0 months | 3 months | 6 months | 0 – 12 months |
| Screening | x | | | | |
| Study A invitation | x | | | | |
| Study B invitation | | x | | | |
| Study C invitation | | | | | x |
| Study D invitation | | x | | | |
| Informed consent study A | | x | | | |
| Informed consent study B | | | | | x |
| Informed consent study C | | | | | x |
| Informed consent study D | | | | | x |
| POC HbA1c | | x | x | x | |
| Demographics | | x | | | |
| Baseline psychometric questionnaires | | | x | | |
| Intervention | | | x | | |
| Endpoint psychometric questionnaires | | | | x | |
| Acceptability survey | | | | x | |
| Barriers to participation survey | | x | | | |
| Patient interviews (study B) | | | | | x |
| Patient interviews (study D) | | | | | x |
| Healthcare professional interviews | | | | | x |

##### 11.3 Appendix 3 – Amendment History

| Amendment No. | Protocol version no. | Date issued | Author(s) of changes | Details of changes made |
|---|---|---|---|---|
| - | 1.0.0 | 03/12/2019 | | Initial protocol document. |
| 1 | 2.0.0 | 01/10/2020 | Sam Westall | Significant amendments to research protocol: methods altered to reduce patient contact and change study focus from primary care to secondary care (cf. COVID). |
| 2 | 2.0.1 | 12/10/2020 | Sam Westall | Revision of protocols: supervisory team feedback. |
| 3 | 2.0.2 | 23/10/2020 | Sam Westall | Updated CI to Dr N Furlong. Updated patient facing documents: SU group feedback. |
| 4 | 2.0.3 | 07/12/2020 | Sam Westall | Updated protocol in response to peer reviewer feedback – clarifying rationale, recruitment population, stratification, PIS sheets, CI and co-investigator CV's added. |
| 5 | 2.0.4 | 21/12/2020 | Sam Westall | Amendments to protocol following Edge Hill University project registration feedback:<br>- In the event of poor recruitment into the main study (A) and in response to supervisor team and service user group feedback, study D added to evaluate barriers to participation in the main study.<br>- Survey assessment of barriers to participation added for those declining entry into study A.<br>- Go/no go criteria outlined. |
| 6 | 2.0.5 | 26/02/2021 | Sam Westall | - Updated document wording in response to sponsor feedback in "PROTOCOL CONTRIBUTORS" section, page VIII.<br>- Following correspondence with the Confidentiality Advisory Committee (CAG), confirming application to the CAG for this project would not be required, section 8 updated. |

#### 11.4 Appendix 4 – Study estimated costs summary

**Estimated Costs Document** 14/01/2021

| Item | Cost per item | Number required | Total cost | Notes |
|---|---|---|---|---|
| Pack of 25 C5 envelopes | £3.99 | 7 | £27.93 | from WH SMITH |
| Book of 100 2nd class stamps | £61.00 | 2 | £122.00 | From Post Office shop |
| SDSCA licence | £77.32 | 1 | £77.32 | (cost is $100, cost in GBP correct at time of writing) |
| DCA Vantage HbA1c reagent test kit (10 test | £52.00 | 7.2 | £374.40 | |
| Printing | see separate table >> | | £64.21 | |
| PAID licence | Free | 1 | £0.00 | |
| WBQ-12 licence | Free | 1 | £0.00 | |
| EQ-5D-5L licence | Free | 1 | £0.00 | |
| DES-LF licence | Free | 1 | £0.00 | |
| Pack of 100 A4 Laminating pouches | £6.88 | 1 | £6.88 | from Euroffice |
| sealed-envelope.com randomisation servic | Free | 1 | £0.00 | |
| | | **Total** | £672.74 | |

Figures based on:
72 participants in study A
15 participant interviews in study B
15 participant interview in study C (emailed PIS, consent)

| **PRINTING COSTS BREAKDOWN** | Colour? | Sides | Number | Cost |
|---|---|---|---|---|
| **Questionnaires** | | | | |
| SDSCA | yes | 2 | 144 | £8.64 |
| PAID | yes | 1 | 144 | £4.32 |
| EQ-5D-5L | yes | 4 | 144 | £17.28 |
| WBQ-12 | no | 1 | 144 | £0.04 |
| DES-LF | no | 4 | 144 | £0.17 |
| Study A Acceptability survey v 1.0.0 | yes | 4 | 50 | £6.00 |
| Study A Barriers to participation survey v 1.0.0 | yes | 3 | 22 | £1.98 |
| **Consent forms**: three copies per person (one for participant, one for researcher, one for patient notes) | | | | |
| Study A v 1.0.0 | yes | 1 | 216 | £6.48 |
| Study B v 1.0.0 | yes | 1 | 45 | £1.35 |
| Study C v1.0.0 | yes | 1 | 45 | £1.35 |
| Study D v1.0.0 | yes | 1 | 45 | £1.35 |
| **Participant information sheets** | | | | |
| Study A PIS v 1.0.2 | yes | 2 | 72 | £4.32 |
| Study B PIS v 1.0.1 | yes | 2 | 15 | £0.90 |
| Study D PIS v 1.0.0 | yes | 2 | 15 | £0.90 |
| **Leaflets** | | | | |
| HbA1c leaflet v 1.0.0 | yes | 2 | 72 | £4.32 |
| **Letters** | | | | |
| Study A patient invitation letter v 1.0.1 | yes | 1 | 72 | £2.16 |
| Study A patient update letter v 1.0.0 | yes | 1 | 8 | £0.24 |
| Study A GP info letter v 1.0.0 | yes | 1 | 72 | £2.16 |
| Study A GP update letter v 1.0.0 | yes | 1 | 8 | £0.24 |
| | | | **Total** | £64.21 |
| based on cost: 3p per colour sheet, 0.03p per mono sheet | | | | |

#### 11.5 Appendix 5 – Timeline

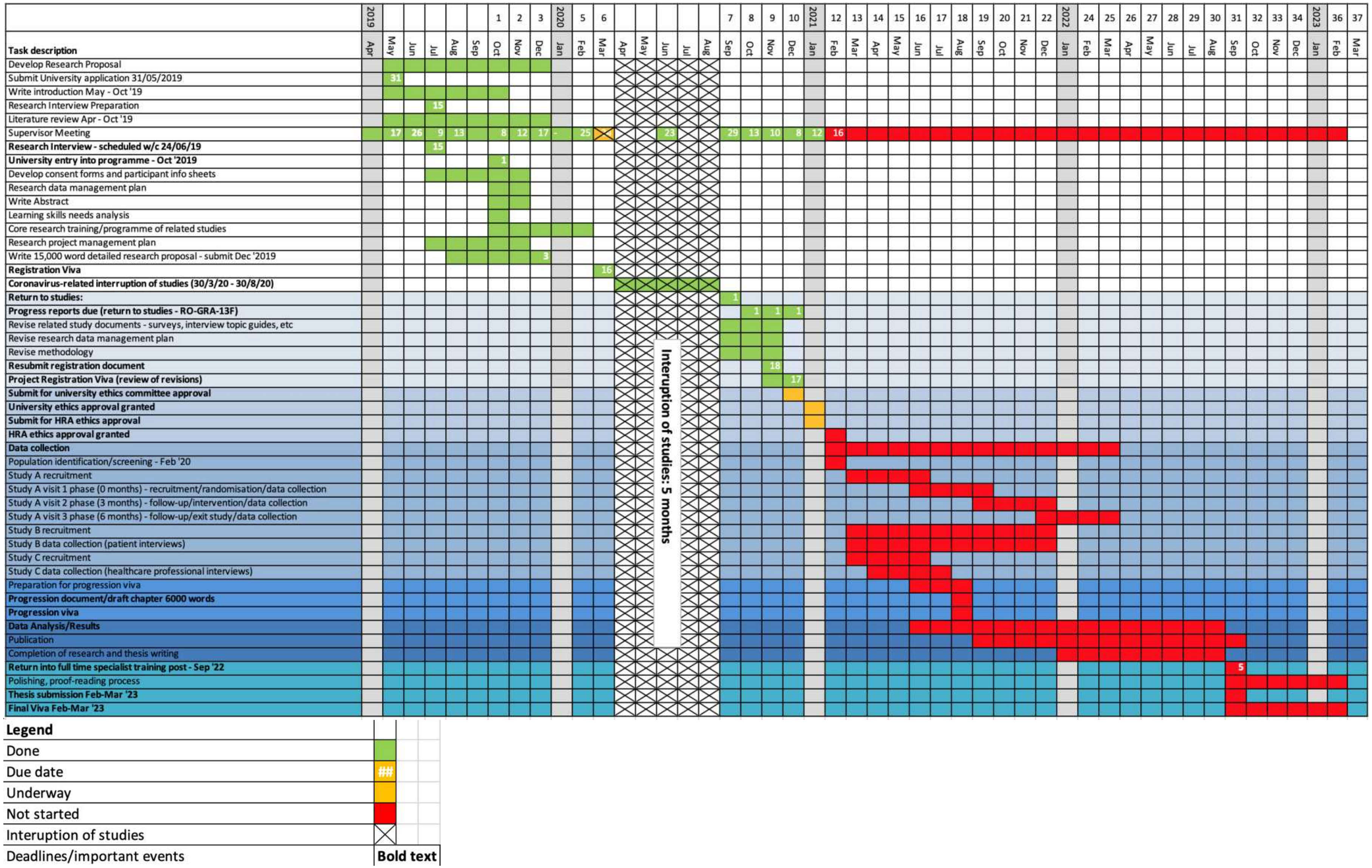
