## Supplementary material for "Psychometric and biomedical outcomes of glycated haemoglobin target-setting in adults with type 1 and type 2 diabetes: A mixed-methods parallel-group randomised feasibility study": S4 patient interview topic guide

**Transcribe the interview as soon as possible after completion.**

Patient interview topic guide and protocol (Study B).
IRAS ID: **291254**

### Diabetes and mental health

**"Do you feel that diabetes has had an effect on your sense of well-being**?"

"In what way?"

"How have you dealt with this?"

"**Do you think that diabetes has affected your mental health**?"

"how/in what way?"

"why do you think that diabetes has had this effect?"

**"What do you think would make you feel better about living with diabetes?"**

### Study Processes

**"How did you feel about being involved in diabetes research?"**

**"People in this study, including you, were randomly put into different groups with different interventions. How do you feel about this?"**

"If you don't mind, why?"

"If you do mind, why?"

**"How did you feel about the time commitments for the study?"**

**"Did you find the leaflet about 'A1c' useful?"**

"if so, in what way?"

"if not, why not?"

**"We testing to see if we were able to keep enough people in the study. You stayed/did not stay in the study until the end. Why was this?"**

"Is there anything that we should have done differently?"

**"Did you find the information sheets and letters in the study easy to understand?"**

"can you give further information on that?"

"how could we make the documents better?"

**"There were a few questionnaires to complete during the study. How did you find these?"**

**"Would you take part in research again?"**

**Probing:**
Active listening to guide further questioning
Appropriate use of silence
Politely interrupt and re-focus the interview if going off-topic
Clarify/elaborate/expand

**Closing:**
"Is there anything else you would like to add?"

Once complete, answer any questions the interviewee may have, stop the recording and thank the interviewee for their participation.

**<u>Transcribe the interview as soon as possible after completion.</u>**
