## Supplementary material for "Psychometric and biomedical outcomes of glycated haemoglobin target-setting in adults with type 1 and type 2 diabetes: A mixed-methods parallel-group randomised feasibility study": S2 HRA letter of approval

Ymchwil Iechyd a Gofal Cymru
Health and Care Research Wales

NHS
Health Research Authority


Dr Niall Furlong
Consultant Physician in Diabetes and Endocrinology
St Helens and Knowsley Teaching Hospitals NHS Trust
Diabetes Centre
St Helens Hospital
Marshalls Cross Road, St Helens
WA9 3DAN/A


11 May 2021

Dear Dr Furlong

### HRA and Health and Care Research Wales (HCRW) Approval Letter

| | |
|---|---|
| **Study title:** | **HbA1c TargeT AchIevemeNt in diabeteS study (ATTAINS study)** |
| **IRAS project ID:** | **291254** |
| **Protocol number:** | **2.0.5** |
| **REC reference:** | **21/SW/0043** |
| **Sponsor** | **St Helens and Knowsley Teaching Hospitals NHS Trust** |

I am pleased to confirm that **HRA and Health and Care Research Wales (HCRW) Approval** has been given for the above referenced study, on the basis described in the application form, protocol, supporting documentation and any clarifications received. You should not expect to receive anything further relating to this application.

Please now work with participating NHS organisations to confirm capacity and capability, in line with the instructions provided in the "Information to support study set up" section towards the end of this letter.

**How should I work with participating NHS/HSC organisations in Northern Ireland and Scotland?**
HRA and HCRW Approval does not apply to NHS/HSC organisations within Northern Ireland and Scotland.

If you indicated in your IRAS form that you do have participating organisations in either of these devolved administrations, the final document set and the study wide governance report (including this letter) have been sent to the coordinating centre of each participating nation. The relevant national coordinating function/s will contact you as appropriate.

Please see IRAS Help for information on working with NHS/HSC organisations in Northern Ireland and Scotland.

**How should I work with participating non-NHS organisations?**
HRA and HCRW Approval does not apply to non-NHS organisations. You should work with your non-NHS organisations to obtain local agreement in accordance with their procedures.

**What are my notification responsibilities during the study?**

The standard conditions document "*After Ethical Review – guidance for sponsors and investigators*", issued with your REC favourable opinion, gives detailed guidance on reporting expectations for studies, including:

- Registration of research
- Notifying amendments
- Notifying the end of the study

The HRA website also provides guidance on these topics, and is updated in the light of changes in reporting expectations or procedures.

**Who should I contact for further information?**
Please do not hesitate to contact me for assistance with this application. My contact details are below.

Your IRAS project ID is **291254**. Please quote this on all correspondence.

Yours sincerely,
Sharon Northey

Approvals Manager

*Copy to:* *Mrs Jeanette Anders, St Helens and Knowsley Teaching Hospitals NHS Trust*

**List of Documents**

The final document set assessed and approved by HRA and HCRW Approval is listed below.

| *Document* | *Version* | *Date* |
|---|---|---|
| GP/consultant information sheets or letters [ATTAINS Study - GP info letter - v1.0.0 - 2020-09-23] | 1.0.0 | 23 September 2020 |
| GP/consultant information sheets or letters [ATTAINS Study - GP update letter - v1.0.0 - 2020-09-21] | 1.0.0 | 21 September 2020 |
| GP/consultant information sheets or letters [ATTAINS Study - Patient update letter- v1.0.0 - 2020-09-21] | 1.0.0 | 21 September 2020 |
| Interview schedules or topic guides for participants [ATTAINS Study – Study B – patient interview guide – v 1.0.0 – 12-10-2020] | 1.0.0 | 12 October 2020 |
| Interview schedules or topic guides for participants [ATTAINS Study – Study C – healthcare professional interview guide – v 1.0.1 – 23-10-2020] | 1.0.1 | 23 October 2020 |
| Interview schedules or topic guides for participants [ATTAINS Study – Study D – patient interview guide – v 1.0.0 – 21-12-2020] | 1.0.0 | 21 December 2020 |
| IRAS Application Form [IRAS_Form_16032021] | | 16 March 2021 |
| IRAS Checklist XML [Checklist_16032021] | | 16 March 2021 |
| IRAS Checklist XML [Checklist_10052021] | | 10 May 2021 |
| Letter from sponsor [Intention to sponsor letter] | STHK-2021-003 | 04 March 2021 |
| Letters of invitation to participant [ATTAINS Study - Invitation letter (studies A and B) – v1.0.2 – 2021-05-04] | 1.0.2 | 04 May 2021 |
| Letters of invitation to participant [ATTAINS Study - Invitation letter (study D) – v1.0.1 – 2021-05-04] | 1.0.1 | 04 May 2021 |
| Letters of invitation to participant [ATTAINS Study - Study C - Healthcare interview email invite - v1.0.1 - 2021-05-04] | 1.0.1 | 04 May 2021 |
| Letters of invitation to participant [ATTAINS Study - Study C - Healthcare interview email reminder - v1.0.1 - 2021-05-04] | 1.0.1 | 04 May 2021 |
| Non-validated questionnaire [ATTAINS Study - Study A - Acceptability survey + Patient enablement instrument - v1.0.0 - 2020-10-06] | 1.0.0 | 06 October 2020 |
| Non-validated questionnaire [Study - Study A - Barriers to Participation survey - v1.0.0 - 2021-01-13] | 1.0.0 | 13 January 2021 |
| Participant consent form [ATTAINS Study - Study A - Consent form - v1.0.1 - 2021-05-04] | 1.0.1 | 04 May 2021 |
| Participant consent form [ATTAINS Study - Study B - Consent form -v1.0.1 - 2021-05-04] | 1.0.1 | 04 May 2021 |
| Participant consent form [ATTAINS Study - Study C - Consent form - v1.0.1 - 2021-05-04] | 1.0.1 | 04 May 2021 |
| Participant consent form [ATTAINS Study - Study D - Consent form - v1.0.1 - 2021-05-04] | 1.0.1 | 04 May 2021 |
| Participant information sheet (PIS) [ATTAINS Study - GDRP patient info - v1.0.0 - 21-09-2020] | 1.0.0 | 21 September 2020 |
| Participant information sheet (PIS) [ATTAINS Study - Study A - Participant Information Sheet - v 1.0.4 - 2021-05-04] | 1.0.4 | 04 May 2021 |
| Participant information sheet (PIS) [ATTAINS Study - Study B - Participant Information Sheet - v 1.0.2 - 2021-05-04] | 1.0.2 | 04 May 2021 |
| Participant information sheet (PIS) [ATTAINS Study - Study C - Healthcare interview PIS - v1.0.2 - 2021-05-04] | 1.0.2 | 04 May 2021 |
| Participant information sheet (PIS) [ATTAINS Study - Study D - Participant Information Sheet - v 1.0.1 - 2021-05-04] | 1.0.1 | 04 May 2021 |
| Referee's report or other scientific critique report [Peer_Review_Form_-_External JW] | n/a | 03 December 2020 |

| | | |
|---|---|---|
| Referee's report or other scientific critique report [Peer_Review_Form_-_Internal SM] | n/a | 23 October 2020 |
| Research protocol or project proposal [ATTAINS Study - Protocol - v2.0.5 - 2021-02-26] | 2.0.5 | 26 February 2021 |
| Sample diary card/patient card [ATTAINS Study - Study A - HbA1c Leaflet - v1.0.1 - 2020-12-07] | 1.0.1 | 07 December 2020 |
| Summary CV for Chief Investigator (CI) [NJF Short Research CV March 21.pdf] | 1.0.0 | 03 March 2021 |
| Summary CV for student [Curriculum-Vitae (research)_Nov 2020] | n/a | 09 November 2020 |
| Summary CV for supervisor (student research) [NRES CV Dec 2020 PN] | n/a | 01 December 2020 |
| Summary, synopsis or diagram (flowchart) of protocol in non technical language [ATTAINS Study - Study processes flowchart - v 1.0.0 - 2020-01-20] | 1.0.0 | 20 January 2021 |
| Validated questionnaire [EQ-5D-5L] | | |
| Validated questionnaire [Summary of Diabetes Self-Care Activities] | | |
| Validated questionnaire [Problems areas in diabetes] | | |
| Validated questionnaire [Diabetes Empowerment Scale (Long Form)] | | |
| Validated questionnaire [Wel-being Questionnaire 12] | | |

| IRAS project ID | 291254 |
|---|---|

#### Information to support study set up

The below provides all parties with information to support the arranging and confirming of capacity and capability with participating NHS organisations in England and Wales. This is intended to be an accurate reflection of the study at the time of issue of this letter.

| Types of participating NHS organisation | Expectations related to confirmation of capacity and capability | Agreement to be used | Funding arrangements | Oversight expectations | HR Good Practice Resource Pack expectations |
|---|---|---|---|---|---|
| This is a non-commercial single site study taking place in the NHS where that single NHS organisation is also the study sponsor. Therefore there is only one site type involved in the research. | This is a single site study sponsored by the participating NHS organisation. You should work with your sponsor R&D office to make arrangements to set up the study. The sponsor R&D office will confirm to you when the study can start following issue of HRA and HCRW Approval. | This is a non-commercial single site study taking place in the NHS where that single NHS organisation is also the study sponsor. Therefore no study agreements are expected. | Study funding is part of a fellowship award. | A Principal Investigator is expected to be in place at the NHS organisation. | Local staff in participating organisations in England who have a contractual relationship with the organisation will undertake the expected activities. Therefore no honorary research contracts or letters of access are expected for this study. |

##### Other information to aid study set-up and delivery

| *This details any other information that may be helpful to sponsors and participating NHS organisations in England and Wales in study set-up.* |
|---|
| The applicant has indicated that they do not intend to apply for inclusion on the NIHR CRN Portfolio. |
